# Muscle Functional Capacity Modifies the Association Between Adiposity and Sarcopenia: Evidence from Two Population-Based Cohorts

**DOI:** 10.64898/2026.08.01.26359483

**Authors:** Shi Li, Yu-rong Chai

**Affiliations:** The First Clinical Medical College, Zhengzhou University, Zhengzhou, Henan, China; School of Basic Medical Sciences, Zhengzhou University, Zhengzhou, Henan, China

**Keywords:** sarcopenia, obesity paradox, grip strength, relative grip strength, CHARLS, NHANES, effect modification, muscle functional capacity

## Abstract

**Background:** Whether adiposity is protective against or detrimental to skeletal muscle health in older adults remains unresolved. The conflicting associations between adiposity and sarcopenia, ranging from apparently protective to harmful effects, have been described as the “obesity paradox”. We investigated whether this paradox could be explained by heterogeneity in muscle functional capacity, hypothesising that the adiposity–sarcopenia relationship is modified by relative grip strength (RGS).

**Methods:** We conducted a cross-sectional analysis of the China Health and Retirement Longitudinal Study (CHARLS; n=15,701), with independent external validation in the US National Health and Nutrition Examination Survey (NHANES; n=10,730). RGS was defined as maximal grip strength divided by body weight. To minimise selective reporting, we performed a prespecified systematic screen of 536 interaction terms derived from 10 anthropometric exposures, 33 functional modifiers, and two sarcopenia outcomes. Core findings were evaluated through cross-metric and cross-outcome replication, sensitivity analyses addressing concerns regarding diagnostic circularity and mathematical coupling, and mediation analyses exploring potential biological pathways.

**Findings:** Among 536 tested interactions, 36 met the Bonferroni-corrected significance threshold, and 32 (89%) involved grip-related modifiers. The interaction between waist circumference and RGS for possible sarcopenia was highly significant (p=6.19 × 10⁻²⁴). Stratified analyses showed that higher adiposity was associated with lower odds of sarcopenia, but the magnitude of this association differed substantially by RGS. For BMI, the inverse association was approximately 10-fold stronger among individuals with high RGS than among those with low RGS (OR 0.64, 95% CI 0.60–0.69 vs OR 0.97, 95% CI 0.96–0.98). Similar effect modification patterns were observed across four anthropometric measures and both sarcopenia outcomes, and were independently replicated in NHANES (p<1.0 × 10⁻¹⁶). The interaction was no longer evident after restricting analyses to participants with preserved grip strength (p=0.65). Mediation analyses suggested that the association was predominantly direct, with triglycerides accounting for 10.5% of the total effect.

**Interpretation:** The association between adiposity and sarcopenia is strongly modified by relative grip strength and appears to be concentrated among individuals with preserved muscle functional capacity. These findings provide a potential explanation for heterogeneity underlying the obesity paradox and suggest that integrating grip strength assessment into adiposity evaluation may improve risk stratification for sarcopenia in older adults.

## Introduction

Whether adiposity is protective against or detrimental to skeletal muscle aging in older adults remains unresolved. Obesity is an established risk factor for numerous chronic diseases; however, among older populations, higher body mass index (BMI) and greater adiposity have paradoxically been associated with lower prevalence of sarcopenia in several epidemiological studies [1]. This phenomenon, often discussed within the broader context of the “obesity paradox” in aging, challenges the assumption that adiposity exerts uniformly harmful effects on skeletal muscle health. Whether these observations represent genuine biological effects or arise from residual confounding, survival bias, and incomplete characterization of body composition remains actively debated.

Several mechanisms have been proposed to explain this counterintuitive association. Greater nutritional reserve and energy availability may provide protection against age-related catabolic stress [2], whereas increased mechanical loading from higher body mass may stimulate skeletal muscle adaptation through mechanotransduction pathways [3]. However, adiposity represents a heterogeneous biological state that encompasses both metabolically harmful components, such as visceral fat accumulation, chronic inflammation, and insulin resistance, and potentially adaptive components related to energy storage and mechanical demand [4]. Therefore, conventional anthropometric measures, including BMI and waist circumference, may not fully capture the complex relationship between adiposity and muscle health. Previous studies have predominantly examined the average association between adiposity and sarcopenia, implicitly assuming that this relationship is consistent across individuals. This assumption may obscure clinically relevant heterogeneity if the consequences of adiposity depend on the functional state of skeletal muscle. In other words, the same degree of adiposity may have different implications among individuals with preserved versus impaired muscle function. Identifying such effect modifiers may therefore provide a new framework for understanding the heterogeneity underlying the obesity paradox in sarcopenia.

Relative grip strength (RGS), defined as maximal grip strength normalized to body weight, may serve as an accessible operational indicator of muscle functional capacity relative to body size. Unlike absolute grip strength, which reflects force-generating capacity without accounting for body size, RGS captures the relationship between muscular strength and the mechanical demand imposed by body mass. Individuals with preserved RGS may have sufficient functional capacity to tolerate or respond to the mechanical and nutritional demands associated with higher body mass, whereas individuals with lower RGS may have reduced capacity to benefit from these potential advantages, allowing adverse metabolic consequences of adiposity to become more prominent. Thus, RGS may represent a previously unrecognized dimension contributing to heterogeneity in the adiposity–sarcopenia relationship.

Despite its biological plausibility, whether RGS modifies the association between adiposity and sarcopenia has not been systematically investigated. Existing studies have primarily examined grip strength as a diagnostic component of sarcopenia or as an adjustment variable in regression models rather than as an effect modifier [5]. Methodological concerns, including the potential for diagnostic circularity and mathematical coupling when grip strength is used as both a diagnostic component and a modifier, may have further limited investigation of this hypothesis. Consequently, the role of relative muscle function in shaping the adiposity–sarcopenia relationship remains insufficiently explored. To address this gap, we conducted a large-scale, two-stage population-based investigation using the China Health and Retirement Longitudinal Study (CHARLS) [6] and the US National Health and Nutrition Examination Survey (NHANES) [7]. We first performed a prespecified systematic interaction screen across 536 combinations of anthropometric exposures, functional modifiers, and sarcopenia outcomes, including possible sarcopenia and confirmed sarcopenia, to identify dominant effect modification patterns while minimizing selective reporting. We then evaluated the reproducibility of core findings through independent external validation, cross-metric and cross-outcome replication, targeted sensitivity analyses addressing potential methodological artifacts, and mediation analyses exploring potential biological pathways. We hypothesized that RGS modifies the association between adiposity and sarcopenia, with stronger inverse associations among individuals with higher RGS, and that this pattern would be consistent across multiple adiposity indicators, sarcopenia definitions, and population settings.

## Methods

### Study Design and Data Sources

This study utilized a cross-sectional design with external validation in an independent population-based dataset. The primary analysis was conducted using the China Health and Retirement Longitudinal Study (CHARLS) [6], and the core findings were subsequently evaluated in the U.S. National Health and Nutrition Examination Survey (NHANES) [7].

For CHARLS, we used data from Wave 3 (2015), as this was the only wave containing standardized grip strength measurements required for the present analysis. Because grip strength measurements were unavailable in other CHARLS waves, the current study was designed as a cross-sectional analysis rather than a longitudinal analysis. Among 25,586 community-dwelling adults aged ≥45 years initially identified from Wave 3, participants with missing data on grip strength, anthropometric measurements, sarcopenia-related variables, or covariates required for the fully adjusted models were excluded. The final CHARLS analytic sample was established after applying these predefined eligibility criteria. For NHANES, 51,089 participants were initially identified from available survey cycles. After applying the same eligibility criteria as in CHARLS and excluding participants with missing data on grip strength, anthropometric measurements, sarcopenia-related variables, or covariates required for the fully adjusted models, 10,730 participants were included in the final analytic sample for external validation.

### Study Population

Participants were excluded if they lacked data on grip strength, body weight, height, waist circumference, or any covariates included in the fully adjusted models. The final analytic samples were established after complete-case analysis. Baseline characteristics were summarized across tertiles of the grip strength-to-weight ratio using means and standard deviations for continuous variables and frequencies with percentages for categorical variables. Group comparisons were performed using ANOVA for continuous variables and chi-square tests for categorical variables (Table 1).

**Table 1.** Baseline characteristics of the CHARLS study population, stratified by grip strength-to-weight ratio tertiles. Continuous variables are presented as mean ± standard deviation and were compared across tertiles using one-way analysis of variance (ANOVA). Categorical variables are presented as frequency (percentage) and were compared using chi-square tests. The grip strength-to-weight ratio was calculated as maximal grip strength (kg) divided by body weight (kg) and categorized into tertiles: Low, Medium, and High. Abbreviations: BMI, body mass index; WHtR, waist-to-height ratio; ABSI, A Body Shape Index; WWI, Weight-adjusted Waist Index; CMI, Cardiometabolic Index; CRP, C-reactive protein; TG, triglycerides; HDL, high-density lipoprotein cholesterol; LDL, low-density lipoprotein cholesterol; eGFR, estimated glomerular filtration rate.

| Characteristic | Tertile 1 (Low) | Tertile 2 (Medium) | Tertile 3 (High) |
| --- | --- | --- | --- |
| **N** | 5234 | 5233 | 5234 |
| **Age, years** | 62.0 ± 11.0 | 59.0 ± 10.2 | 57.1 ± 9.5 |
| **Male, n (%)** | 1005 (19.2%) | 2267 (43.3%) | 4093 (78.2%) |
| **BMI, kg/m<sup>2</sup>** | 27.0 ± 39.8 | 24.3 ± 11.4 | 22.5 ± 6.0 |
| **Waist circumference, cm** | 90.4 ± 10.6 | 86.6 ± 9.8 | 82.6 ± 9.2 |
| **Grip strength, kg** | 21.5 ± 6.2 | 30.6 ± 6.3 | 39.7 ± 7.9 |
| **Grip strength/weight** | 0.350 ± 0.075 | 0.509 ± 0.037 | 0.680 ± 0.086 |
| **Possible sarcopenia, n (%)** | 1848 (52.4%) | 402 (15.2%) | 85 (4.0%) |
| **Sarcopenia, n (%)** | 684 (14.5%) | 104 (2.0%) | 9 (0.2%) |
| **Multimorbidity, n (%)** | 1866 (35.7%) | 1357 (25.9%) | 1000 (19.1%) |

### Key Variables and Definitions

The primary exposure was central obesity, assessed by waist circumference (cm). General obesity, assessed by body mass index (BMI, kg/m²), was considered a secondary exposure because of its widespread clinical application and accessibility in community-based screening.

The primary candidate effect modifier was relative grip strength (RGS), defined as maximal grip strength (kg) divided by body weight (kg). In CHARLS, grip strength was measured using a dynamometer, and the maximum value obtained from the dominant hand was used. In NHANES, grip strength was assessed using a comparable dynamometer protocol, and the combined grip strength from both hands was used according to the NHANES examination protocol. RGS was categorized into tertiles for visualization of dose-response patterns and dichotomized at the median for stratified analyses to facilitate interpretation of effect modification patterns.

The primary outcome was possible sarcopenia, defined according to the Asian Working Group for Sarcopenia (AWGS) 2019 criteria [5] as reduced muscle strength accompanied by impaired physical performance. Specifically, possible sarcopenia was defined as grip strength <28 kg in men or <18 kg in women, combined with either gait speed <1.0 m/s or a five-time chair stand test duration ≥12 seconds. The secondary outcome was confirmed sarcopenia, which additionally required reduced muscle mass, defined as appendicular skeletal muscle mass index <7.0 kg/m² in men or <5.7 kg/m² in women, according to AWGS 2019 criteria [5].

### Covariates

Covariates were selected a priori based on previous literature and biological plausibility regarding the associations between adiposity, muscle function, and sarcopenia. Three progressively adjusted logistic regression models were constructed. Model 1 adjusted for basic demographic characteristics, including age, sex, education level, and marital status. Model 2 additionally adjusted for lifestyle factors (smoking status and alcohol consumption) and multimorbidity. Multimorbidity was defined as the presence of 10 self-reported chronic conditions, including hypertension, diabetes, heart disease, stroke, kidney disease, chronic lung disease, arthritis, psychiatric disorders, cancer, and dyslipidemia. Model 3 further adjusted for BMI in analyses using waist circumference as the primary exposure, allowing the association of central adiposity to be evaluated independently of overall body size.

### Interaction Screening Framework

To avoid selective reporting and to identify whether grip-related modifiers dominated the effect modification landscape, we performed a systematic, pre-specified exhaustive interaction screening. A three-layer variable pool was constructed: 10 exposure variables (including waist circumference, BMI, waist-to-height ratio, body weight, and six derived adiposity indices such as A Body Shape Index [ABSI], Weight-adjusted Waist Index [WWI], and Cardiometabolic Index [CMI]), 33 candidate modifiers (including grip strength-to-weight ratio, absolute grip strength, grip-BMI ratio, and other physical performance metrics), and 2 outcomes (possible sarcopenia and confirmed sarcopenia). After excluding structurally invalid combinations, including interactions between mathematically dependent variables (e.g., when a candidate modifier was directly derived from the corresponding exposure variable), a total of 536 interaction terms were systematically evaluated.

Each interaction term was evaluated using logistic regression with the same covariate adjustment as Model 2. Interaction p-values were extracted and ranked across all tested combinations. Bonferroni correction was applied to control the family-wise error rate (FWER) at 0.05. Given the 536 interaction tests performed, the Bonferroni-adjusted significance threshold was calculated as 0.05/536, corresponding to p < 9.3 × 10⁻⁵. False discovery rate (FDR) correction using the Benjamini–Hochberg procedure was additionally performed as a complementary approach to identify potentially meaningful signals while accounting for multiple comparisons. Signals were classified into three tiers: Grade 1 (Bonferroni significant), Grade 2 (FDR significant), and Grade 3 (nominal p < 0.05). The complete results of all 536 interaction terms are publicly reported in Supplementary Table S1.

### Primary and Secondary Analyses

The core analysis tested the interaction between waist circumference and grip strength-to-weight ratio on possible sarcopenia. The shape of the association was visualized using restricted cubic spline (RCS) curves with three knots, stratified by tertiles of the grip/weight ratio, adjusted for Model 2 covariates. Non-linear p-values were reported.

Stratified logistic regression was then performed to estimate odds ratios (ORs) and 95% confidence intervals (CIs) for the association between each anthropometric measure and possible sarcopenia, separately in high versus low grip/weight groups (dichotomized at the median). Cross-metric replication was assessed by repeating the stratified analysis for waist circumference, BMI, waist-to-height ratio, and body weight.

### Sensitivity Analyses

Five pre-specified sensitivity analyses were conducted to evaluate the robustness of the primary findings and to assess whether alternative explanations could account for the observed effect modification.

First, to assess whether extreme triglyceride measurements influenced the observed association, participants with triglyceride values truncated at assay detection limits were excluded, and the primary interaction analyses were repeated.

Second, to evaluate whether extreme body size values affected the results, participants in the highest and lowest 1% of the BMI distribution were excluded, followed by re-analysis of the primary interaction models.

Third, to assess whether the observed interaction could be attributed to mathematical coupling between relative grip strength and anthropometric measures, absolute grip strength was substituted for the grip strength-to-weight ratio as the candidate effect modifier. Because relative grip strength incorporates body weight in its calculation, repeating the analyses with absolute grip strength provided an approach to evaluate the potential contribution of shared mathematical components.

Fourth, to examine whether the observed interaction was driven by the inclusion of grip strength within the sarcopenia diagnostic framework, analyses were restricted to participants whose grip strength exceeded the AWGS 2019 thresholds for low muscle strength (≥28 kg for men and ≥18 kg for women) [5]. Persistence of the interaction in this subgroup would suggest that the findings were not solely attributable to overlap between the modifier and outcome definition.

Fifth, to evaluate cross-outcome consistency, the primary interaction between waist circumference and relative grip strength was repeated using confirmed sarcopenia as the outcome instead of possible sarcopenia.

### Mediation Analysis

To explore whether metabolic pathways potentially contributed to the observed association between waist circumference and sarcopenia risk, mediation analyses were performed. Three candidate mediators were selected based on their established relevance to obesity-related metabolic dysfunction: C-reactive protein (inflammation), fasting glucose (insulin resistance), and triglycerides (lipid metabolism). The total effect of waist circumference on possible sarcopenia was decomposed into the average direct effect (ADE) and the average causal mediation effect (ACME). The proportion mediated was calculated as the ratio of ACME to the total effect. Stratified mediation analyses were further conducted among participants with high and low relative grip strength to assess whether potential mediation patterns differed according to muscle functional status. All mediation models were adjusted for Model 2 covariates plus BMI, and uncertainty was quantified using bootstrapped 95% confidence intervals based on 1,000 simulations.

### External Validation in NHANES

All core interaction findings were evaluated in NHANES using the same analytical framework as the primary CHARLS analysis. The relative grip strength ratio was calculated as maximal grip strength divided by body weight, and tertile cutoffs were derived from the NHANES study population. Logistic regression models were adjusted for age, sex, education level, smoking status, alcohol consumption, and multimorbidity. For analyses involving waist circumference, BMI was additionally adjusted to distinguish central adiposity from overall body size. Because the pre-processed NHANES dataset did not retain survey design variables required for nationally representative estimation, NHANES analyses were interpreted as external validation of association patterns rather than estimates of population-level prevalence or effect sizes.

### Statistical Software

All analyses were performed using R version 4.x [8]. The rms package was used for restricted cubic spline analyses, and the mediation package was used for mediation analyses. Statistical significance was defined as a two-sided p value <0.05 after Bonferroni correction for multiple comparisons where applicable. Variance inflation factors (VIFs) were calculated for covariates included in the fully adjusted models to assess potential multicollinearity.

## Results

### Study Population and Baseline Characteristics

The CHARLS analytic sample included 15,701 participants (mean age, 62.4 ± 9.8 years; 52.1% women) after excluding participants with missing data on grip strength, anthropometric measurements, or covariates. Relative grip strength (RGS) ranged from 0.08 to 1.23 kg/kg, with a median value of 0.47 kg/kg. Across RGS tertiles, participants with lower RGS were older, more likely to be women, had higher BMI and waist circumference, and showed a higher prevalence of possible sarcopenia (85.2% vs. 28.6% in the highest tertile) and multimorbidity (Table 1).

The NHANES external validation sample included 10,730 participants (mean age, 58.7 ± 11.4 years; 51.5% women) with complete information required for the fully adjusted models.

### Systematic Interaction Screen

Among the 536 interaction terms systematically evaluated, 36 reached the Bonferroni-adjusted significance threshold (p < 9.3 × 10⁻⁵) and were classified as Grade 1 signals. An additional 35 interactions remained significant after false discovery rate correction (Grade 2), and 38 additional interactions reached nominal significance (Grade 3) (Table S1).

Grip-related modifiers, including RGS, absolute grip strength, and grip strength-to-BMI ratio, accounted for 32 of the 36 Grade 1 signals (89%). These signals were consistently observed across multiple anthropometric measures, including waist circumference, BMI, waist-to-height ratio, body weight, derived adiposity indices, and both possible and confirmed sarcopenia outcomes.

### Core Finding: Waist Circumference × RGS Interaction on Possible Sarcopenia

The interaction between waist circumference and RGS for possible sarcopenia was highly significant. In the fully adjusted model (Model 2: adjusted for age, sex, education, marital status, smoking status, alcohol consumption, and multimorbidity, with additional adjustment for BMI), the interaction p-value was 6.19 × 10⁻²⁴.

This association remained significant after Bonferroni correction for the five core interaction signals (adjusted p = 3.10 × 10⁻²³) and across progressively adjusted models (Model 1: p = 2.03 × 10⁻²³; Model 3: p = 1.06 × 10⁻²³).

Restricted cubic spline analyses demonstrated distinct associations between waist circumference and possible sarcopenia across RGS tertiles (Figure 1). Among participants in the highest RGS tertile, the adjusted predicted probability of possible sarcopenia decreased from 26.2% at a waist circumference of 60 cm to 0.2% at 100 cm. In contrast, among participants in the lowest RGS tertile, the probability decreased from 97.7% to 68.3%. The nonlinear component of the interaction was significant (p for nonlinearity = 1.31 × 10⁻⁵).

**Figure 1.**
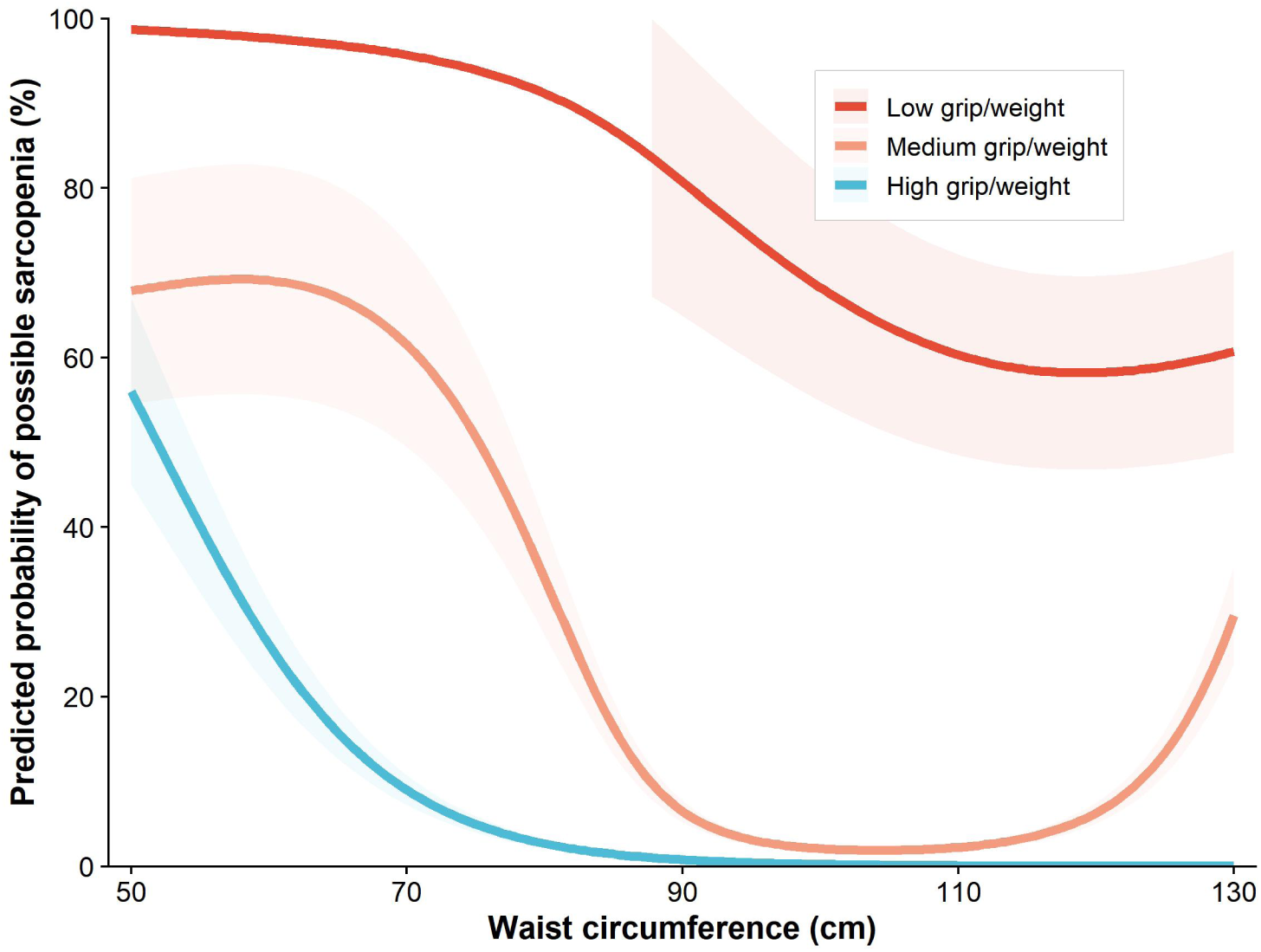
Restricted cubic spline (RCS) curves for the association between waist circumference and possible sarcopenia, stratified by grip strength-to-weight ratio tertiles. Curves represent the adjusted predicted probability of possible sarcopenia (y-axis) across waist circumference (x-axis), derived from logistic regression models with three knots. Adjustments include age, sex, education, marital status, smoking status, alcohol consumption, and multimorbidity. Shaded areas represent 95% confidence intervals. The grip strength-to-weight ratio was categorized into tertiles: Low (red), Medium (orange), and High (blue). The inverse association between waist circumference and sarcopenia risk is most pronounced in the high grip/weight tertile and substantially attenuated in the low tertile, demonstrating systematic effect modification. Non-linear p-value = 1.31 × 10⁻⁵.

In stratified analyses using the median RGS cutoff, waist circumference showed inverse associations with possible sarcopenia in both RGS groups. Each 1-cm increase in waist circumference was associated with lower odds of possible sarcopenia among participants with high RGS (adjusted OR = 0.969, 95% CI: 0.941–0.998, p = 3.51 × 10⁻²) and low RGS (adjusted OR = 0.947, 95% CI: 0.941–0.953, p = 7.00 × 10⁻⁶²) (Figure 2).

**Figure 2.**
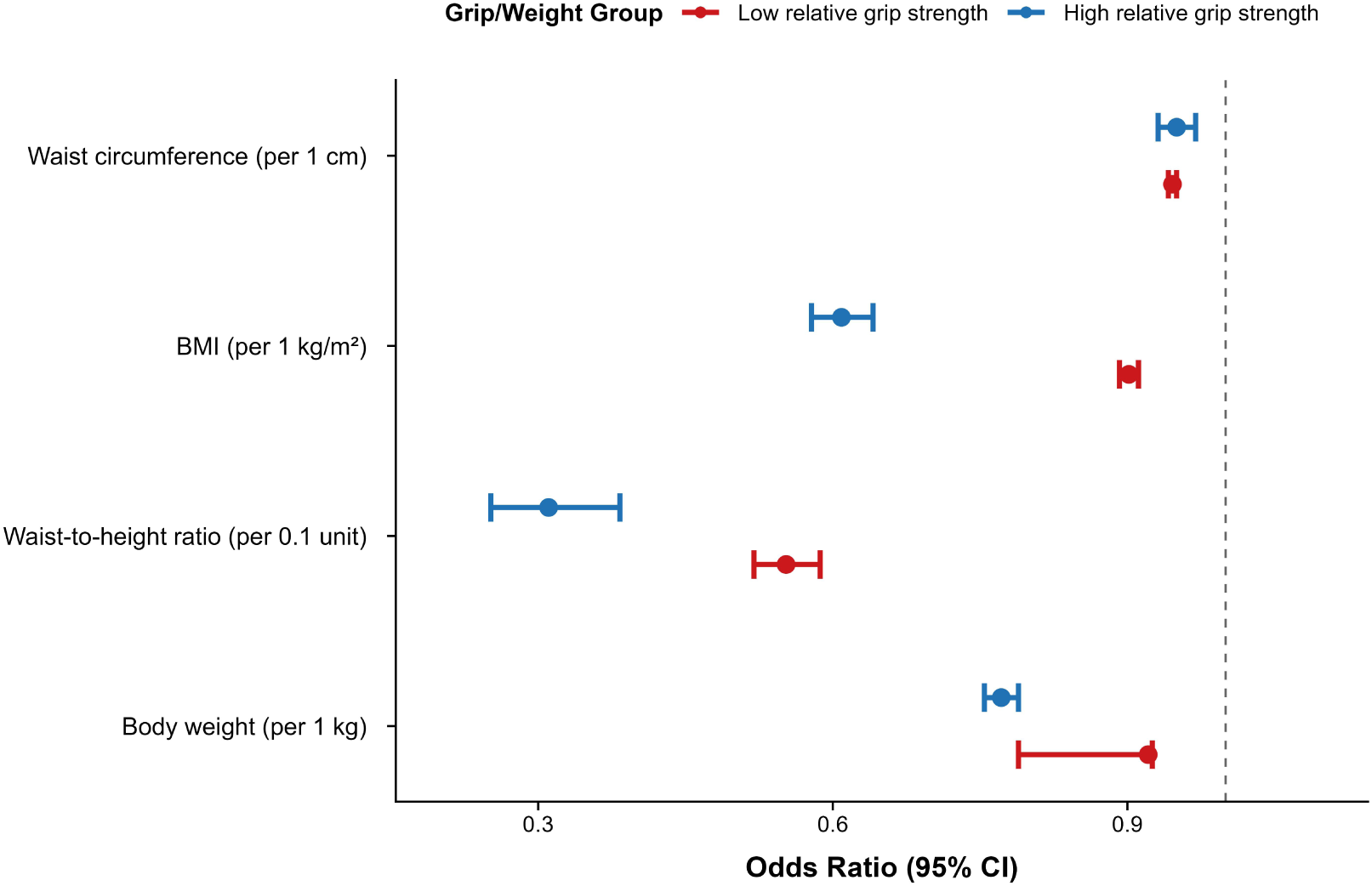
Stratified odds ratios (ORs) for the associations between four anthropometric measures and possible sarcopenia, by grip strength-to-weight ratio (high vs. low). ORs were estimated from logistic regression models adjusted for age, sex, education, marital status, smoking status, alcohol consumption, and multimorbidity. For waist circumference, BMI was additionally adjusted. The grip/weight ratio was dichotomized at the median. Blue squares represent the high grip/weight group; red circles represent the low grip/weight group. Error bars indicate 95% confidence intervals. The dashed vertical line marks OR = 1. All four anthropometric measures—waist circumference, BMI, waist-to-height ratio (WHtR), and body weight—exhibit a consistent pattern: the inverse association with sarcopenia is stronger in the high grip/weight group, demonstrating cross-metric replication of the effect modification.

### Cross-Metric Replication: BMI and Other Anthropometric Measures

The effect modification pattern was replicated when BMI replaced waist circumference as the adiposity measure. The interaction between BMI and RGS was highly significant in the fully adjusted model (p = 5.83 × 10⁻¹⁰⁴).

Stratified analyses demonstrated stronger inverse associations between BMI and possible sarcopenia among participants with high RGS (adjusted OR per 1-unit increase in BMI = 0.642, 95% CI: 0.599–0.685) compared with participants with low RGS (adjusted OR = 0.965, 95% CI: 0.963–0.978). Similar patterns were observed for waist-to-height ratio (interaction p = 2.83 × 10⁻¹⁷) and body weight (interaction p = 4.41 × 10⁻¹¹³), with consistent directions of effect modification across all four anthropometric measures (Figure 2).

### Cross-Outcome Replication: Confirmed Sarcopenia

When confirmed sarcopenia was used as the outcome, the interaction between waist circumference and RGS remained significant across adjustment models.

The interaction remained significant in Model 2 (p = 1.28 × 10⁻¹⁰) and Model 3 with additional BMI adjustment (p = 8.89 × 10⁻¹⁵). The association remained significant after Bonferroni correction for the five core interaction signals (adjusted p = 4.44 × 10⁻¹⁴).

### Sensitivity Analyses

All five core interaction signals remained significant after excluding participants with triglyceride values affected by assay truncation (n = 240 excluded) and after excluding participants in the highest and lowest 1% of BMI distribution (n = 9,699 excluded) (Table S2; Figure S1).

Additional sensitivity analyses were conducted to evaluate potential methodological explanations. When absolute grip strength replaced RGS as the effect modifier, the interaction between BMI and absolute grip strength on possible sarcopenia was not significant (p = 0.63).

When analyses were restricted to participants with grip strength above the AWGS 2019 diagnostic thresholds (≥28 kg for men and ≥18 kg for women), the interaction was no longer significant (p = 0.65) (Figure S1; Table S2).

### Mediation Analysis

Mediation analyses were performed to explore whether metabolic biomarkers contributed to the association between waist circumference and possible sarcopenia.

Among the three evaluated pathways, triglycerides accounted for 10.5% of the total association (ACME p = 0.022). In contrast, CRP (proportion mediated: −2.2%, ACME p = 0.018) and fasting glucose (proportion mediated: −0.7%, ACME p = 0.810) showed limited mediation effects (Figure 4; Table S3).

Stratified mediation analyses showed no significant mediation effects among participants with high RGS. Among participants with low RGS, triglycerides mediated a small proportion of the association, whereas CRP and glucose remained non-significant.

### External Validation in NHANES

In the NHANES external validation sample, the interaction pattern between adiposity measures and RGS was consistently reproduced.

For waist circumference, the interaction p-value fell below the numerical precision threshold of the statistical software (p < 1 × 10⁻¹⁶).

Stratified analyses showed stronger inverse associations between waist circumference and possible sarcopenia among participants with high RGS (adjusted OR = 0.783, 95% CI: 0.773–0.793) compared with participants with low RGS (adjusted OR = 0.908, 95% CI: 0.904–0.912).

Similarly, the interaction between BMI and RGS remained highly significant (p < 1 × 10⁻¹⁶), with consistent directions of effect modification between CHARLS and NHANES (Figure 3).

**Figure 3.**
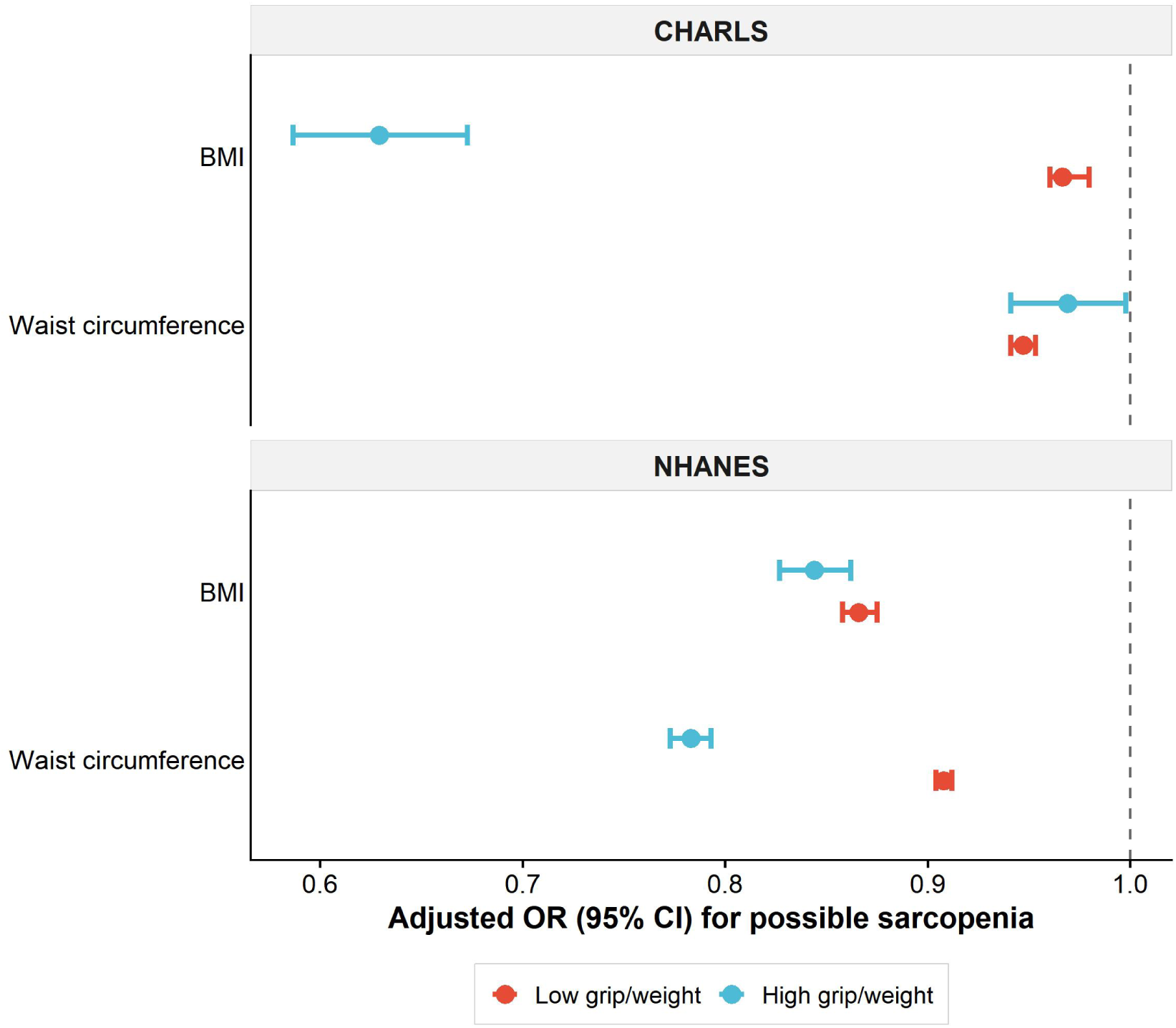
Independent replication of the effect modification in the U.S. National Health and Nutrition Examination Survey (NHANES). Stratified odds ratios for the association between waist circumference (top panel) and BMI (bottom panel) with possible sarcopenia, by grip strength-to-weight ratio (high vs. low), in CHARLS (left) and NHANES (right). Both analyses used logistic regression adjusted for age, sex, education, smoking status, alcohol consumption, and multimorbidity. For waist circumference, BMI was additionally adjusted. In NHANES, the grip/weight ratio was calculated as grip strength divided by body weight, with tertile cutoffs derived from the NHANES sample. The consistent pattern across two independent databases—stronger inverse association in the high grip/weight group—provides cross-population validation. Error bars represent 95% confidence intervals.

**Figure 4.**
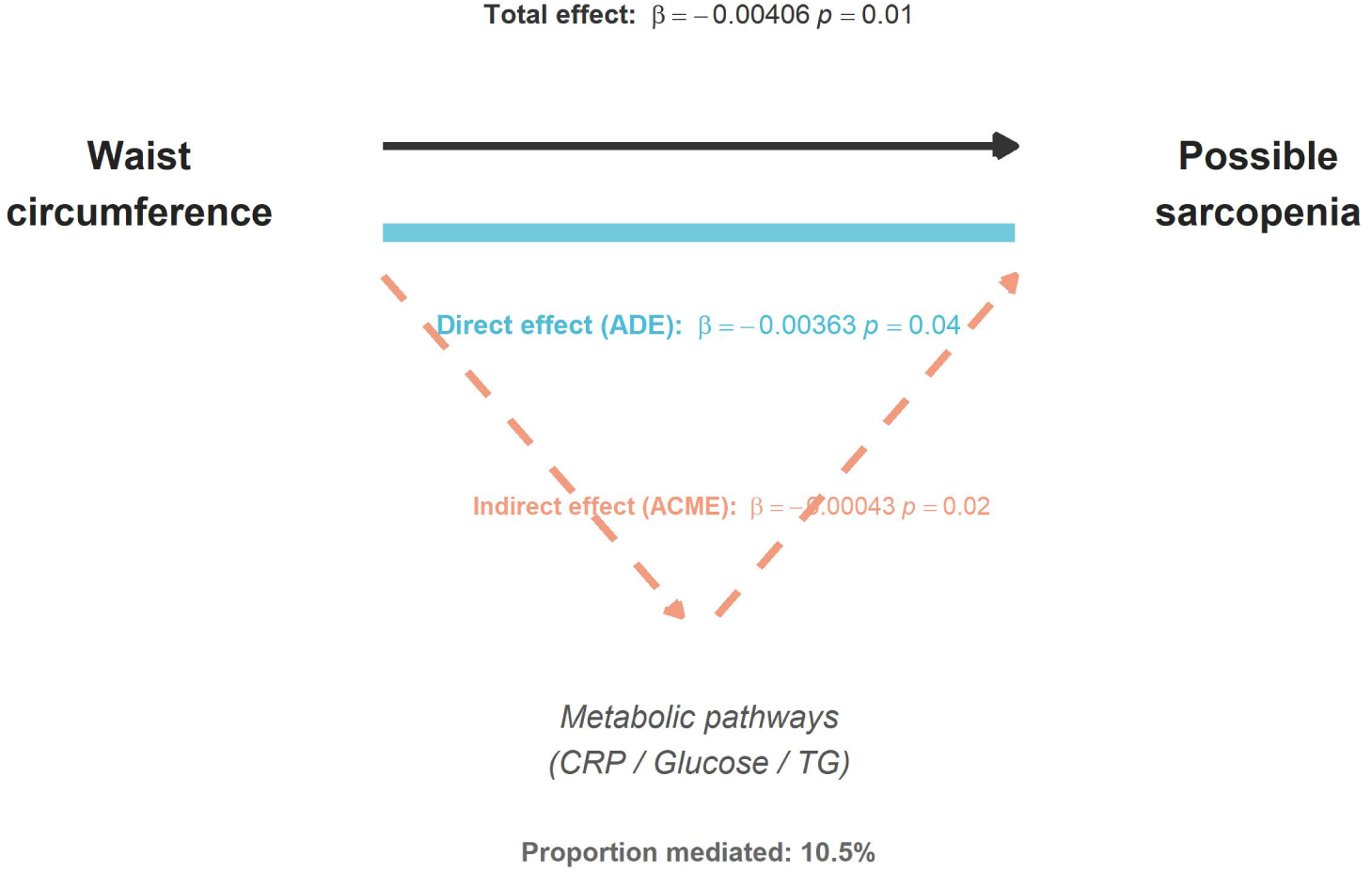
Mediation analysis of metabolic pathways linking waist circumference to possible sarcopenia. The total effect (dark grey, top arrow) represents the overall association between waist circumference and sarcopenia risk. The average direct effect (ADE, blue, middle arrow) represents the effect not transmitted through the three candidate metabolic mediators. The average causal mediation effect (ACME, orange dashed arrows) represents the effect mediated through C-reactive protein (inflammation), fasting glucose (insulin resistance), and triglycerides (lipid metabolism). The proportion mediated was 10.5%, indicating that the inverse association between waist circumference and sarcopenia is predominantly direct rather than mediated through these metabolic pathways. All models were adjusted for age, sex, education, marital status, smoking status, alcohol consumption, multimorbidity, and BMI. Bootstrapped 95% confidence intervals are based on 1,000 simulations.

### Exploratory Analysis: Environmental Risk and RGS

In an exploratory analysis, we examined whether environmental risk exposure further stratified sarcopenia prevalence according to RGS categories.

Participants with both high environmental risk and low RGS showed the highest prevalence of sarcopenia compared with other joint categories. This exploratory finding suggested a potential combined association between environmental exposure burden and reduced muscle functional status (Figure S2).

## Discussion

### Principal Findings

In this large-scale population-based study incorporating systematic interaction screening and independent external validation, we identified relative grip strength (RGS) as a key dimension underlying heterogeneity in the association between adiposity and sarcopenia. Among 536 prespecified interaction terms evaluated in CHARLS, grip-related modifiers accounted for 89% of

Bonferroni-significant signals. The interaction between waist circumference and RGS demonstrated consistent patterns across multiple anthropometric indicators, sarcopenia definitions, sensitivity analyses, and an independent NHANES population.

These findings suggest that the relationship between adiposity and sarcopenia is not uniform across older adults but varies according to underlying muscle functional status. Higher adiposity was associated with lower sarcopenia prevalence among individuals with relatively preserved RGS, whereas this inverse association was substantially attenuated among individuals with lower RGS. Therefore, RGS may represent an important dimension for understanding heterogeneity within the obesity paradox observed in sarcopenia and aging research.

### Rethinking the Obesity Paradox Through Muscle Function

The obesity paradox in older adults has remained a persistent and clinically relevant observation, whereby higher body mass or adiposity is sometimes associated with lower risks of adverse outcomes despite the established metabolic consequences of obesity [1]. Previous explanations have proposed several mechanisms, including greater nutritional reserve, protection against catabolic stress, and survival-related selection effects.

However, adiposity represents a heterogeneous biological state that includes both metabolically detrimental components, such as visceral fat accumulation and inflammation, and potentially adaptive components related to energy storage and mechanical loading. Consequently, a single anthropometric measure may incompletely capture the biological context in which adiposity influences aging-related outcomes.

Previous studies have predominantly examined the average association between adiposity and sarcopenia at the population level, implicitly assuming that this relationship is relatively consistent across individuals. Such an approach may obscure clinically relevant heterogeneity arising from differences in underlying muscle functional status.

Our findings extend this concept by suggesting that muscle functional capacity may represent an important modifier of the adiposity–sarcopenia relationship. Individuals with higher RGS may have greater functional capacity to utilize potential mechanical and nutritional advantages associated with greater body mass, whereas individuals with lower RGS may have reduced capacity to benefit from these effects, allowing the detrimental consequences of excess adiposity to become more apparent.

### A Proposed Muscle–Adiposity Balance Framework

Based on the observed interaction pattern, we propose a muscle–adiposity balance framework to interpret the heterogeneous relationship between adiposity and sarcopenia.

Adiposity may exert competing biological influences. On one hand, greater body mass increases habitual mechanical demands on skeletal muscle, potentially promoting maintenance of muscle function through mechanotransduction pathways, anabolic signaling, and adaptation to loading [3]. On the other hand, excessive adiposity may contribute to chronic inflammation [9], insulin resistance, ectopic lipid accumulation, and impaired muscle quality.

The net association between adiposity and sarcopenia may therefore depend on the balance between these opposing processes and the individual’s functional muscle status. Among individuals with relatively preserved RGS, the potential mechanical and nutritional advantages associated with greater body mass may outweigh metabolic disadvantages, resulting in a stronger inverse association between adiposity and sarcopenia. In contrast, among individuals with reduced RGS, impaired muscle function may indicate a state in which these compensatory mechanisms are insufficient.

Importantly, RGS should be interpreted in this study as an operational measure of relative muscle functional capacity rather than a direct measurement of physiological reserve. The concept of relative functional reserve provides a possible biological interpretation for why individuals with similar levels of adiposity may experience different risks of sarcopenia.

### Biological Interpretation of the Observed Interaction

Several biological mechanisms may contribute to the observed effect modification.

First, mechanical loading represents a potential pathway linking body mass to muscle maintenance. Greater body weight increases habitual mechanical demands on skeletal muscle, which may promote maintenance of strength and function through mechanotransduction pathways [3]. However, this potential benefit likely depends on sufficient muscle capacity to respond to increased loading.

Second, adipose tissue may serve as an energy reservoir during aging-related catabolic stress [2]. Older adults frequently experience periods of illness, inflammation, and reduced nutritional intake, during which greater energy availability may partially buffer against muscle loss. This mechanism may be particularly relevant among individuals with preserved functional capacity.

Third, the limited contribution of conventional metabolic mediators, with triglycerides accounting for only a small proportion of the association, suggests that metabolic pathways may explain only part of the observed interaction [10]. Instead, the findings are compatible with a broader relationship involving body composition, mechanical demand, and functional muscle status. The complex interplay between adipose tissue and skeletal muscle involves bidirectional signaling beyond simple metabolic mediation, including endocrine and mechanical crosstalk [11,12].

These interpretations remain hypothesis-generating because the present study used cross-sectional data and cannot establish temporal relationships or causal direction.

### Robustness of the Effect Modification Pattern

Several methodological considerations require attention because RGS shares mathematical components with body weight and grip strength contributes to sarcopenia definitions [5].

First, the observed interaction was not restricted to a single adiposity indicator. Similar patterns were observed across waist circumference, BMI, waist-to-height ratio, and body weight, suggesting that the finding was not dependent on one specific anthropometric measure. Notably, the interaction was initially identified using waist circumference, an anthropometric measure that does not share body weight as a component with RGS, and subsequently replicated using BMI and other adiposity indices. This consistency suggests that the observed modification pattern is not an artifact of shared mathematical components but reflects a genuine biological interaction.

Second, replacing RGS with absolute grip strength substantially attenuated the interaction, addressing concerns regarding mathematical coupling between relative measures and adiposity variables.

Third, excluding individuals meeting AWGS low-grip-strength thresholds eliminated the interaction pattern. This finding suggests that the observed modification was concentrated among individuals with compromised muscle function rather than representing a statistical artifact caused solely by incorporating grip strength into sarcopenia classification.

Together, the convergence of multiple analytical strategies—including cross-metric replication, substitution with absolute grip strength, restriction to participants with preserved grip strength, cross-outcome replication, and external validation in NHANES—makes it unlikely that the observed effect modification is attributable solely to a single methodological artifact.

### Clinical Implications

The findings may have implications for how adiposity and sarcopenia risk are interpreted in older adults.

Current clinical approaches often emphasize weight reduction among individuals with obesity, while the potential role of preserving muscle function receives increasing attention. Current guidelines for sarcopenic obesity recommend concurrent resistance training during weight loss to prevent muscle loss [4]. However, these recommendations are principle-based and do not specify at what threshold of impaired muscle function the priority should shift from weight management to muscle preservation.

Our findings suggest that adiposity assessment alone may be insufficient for evaluating sarcopenia vulnerability, and that measures reflecting relative muscle function may provide additional clinical information. The magnitude of this effect modification—an approximately 10-fold difference in the BMI–sarcopenia association between high- and low-RGS groups—suggests that RGS-based stratification may have potential relevance for identifying heterogeneous sarcopenia risk profiles in clinical settings. For older adults with reduced RGS, interventions targeting muscle preservation and functional improvement may deserve prioritization before aggressive weight-loss strategies are considered. Conversely, among individuals with preserved muscle function, higher adiposity may not uniformly indicate increased sarcopenia susceptibility. This stratified approach, based on a simple ratio of grip strength to body weight—both readily obtainable in community and primary care settings—represents a practical, low-cost tool that can be implemented without specialized equipment. Future intervention studies are needed to determine whether incorporating functional muscle measures into obesity management strategies can improve personalized prevention and treatment approaches.

### Strengths and Limitations

This study has several strengths. First, the systematic interaction screening framework reduced the possibility of selective reporting and allowed identification of dominant effect modification patterns among a broad range of anthropometric and functional variables. Second, the findings were supported by multiple levels of validation, including cross-metric replication, cross-outcome replication, extensive sensitivity analyses, and independent external validation in NHANES. Third, the mediation analyses provided additional insight into potential biological pathways underlying the observed associations.

Several limitations should also be acknowledged. First, the cross-sectional design prevents determination of temporal relationships and causal inference. While we observed a robust interaction replicated across independent databases, we cannot determine whether low RGS precedes the attenuation of adiposity-associated protection or whether sarcopenic changes lead to declining relative strength [13]. However, the complete disappearance of the interaction in individuals with preserved grip strength argues against reverse causation as the sole explanation. Second, grip strength measurements were available only in specific survey waves, limiting longitudinal evaluation of within-person changes in muscle function and their relationship with evolving adiposity-sarcopenia dynamics. Third, although multiple analyses addressed potential statistical explanations, residual confounding cannot be completely excluded [14]. Fourth, Mendelian randomization analysis could not be performed because suitable genetic instruments for sarcopenia-related outcomes remain unavailable, although instruments for grip strength [15] and BMI [16] exist from large-scale genome-wide association studies.

### Conclusion

In conclusion, this study identifies relative grip strength as an important modifier of the association between adiposity and sarcopenia risk. The findings suggest that the relationship between obesity and sarcopenia is heterogeneous and may depend substantially on underlying muscle functional status. Integrating functional muscle assessment into adiposity evaluation may improve risk stratification and support more individualized strategies for preventing sarcopenia in obese older adults.

## Data Availability

The CHARLS data that support the findings of this study are openly available at http://charls.pku.edu.cn/. The NHANES data are openly available at https://www.cdc.gov/nchs/nhanes/. All 536 interaction terms analyzed in this study are publicly reported in Supplementary Table S1.

https://www.cdc.gov/nchs/nhanes/

## Declarations

### Author contributions

Shi Li: Conceptualization, Methodology, Software, Formal analysis, Investigation, Data curation, Writing – original draft, Writing – review & editing, Visualization.

Yu-rong Chai: Writing – review & editing, Supervision, Project administration, Funding acquisition (for publication costs).

All authors have read and approved the final version of the manuscript and agree to be accountable for all aspects of the work.

### Ethics statement

The CHARLS study was approved by the Biomedical Ethics Review Committee of Peking University (IRB00001052-11015). The NHANES protocol was approved by the National Center for Health Statistics Research Ethics Review Board (Protocol #2011-17, Continuation of Protocol #2011-17). All participants provided written informed consent. The present secondary analysis used de-identified publicly available data and did not require additional ethical approval.

### Consent for publication

Not applicable.

## Acknowledgments

We thank the China Health and Retirement Longitudinal Study (CHARLS) team and the National Health and Nutrition Examination Survey (NHANES) team for making their data publicly available.

---

**Figure S1.**
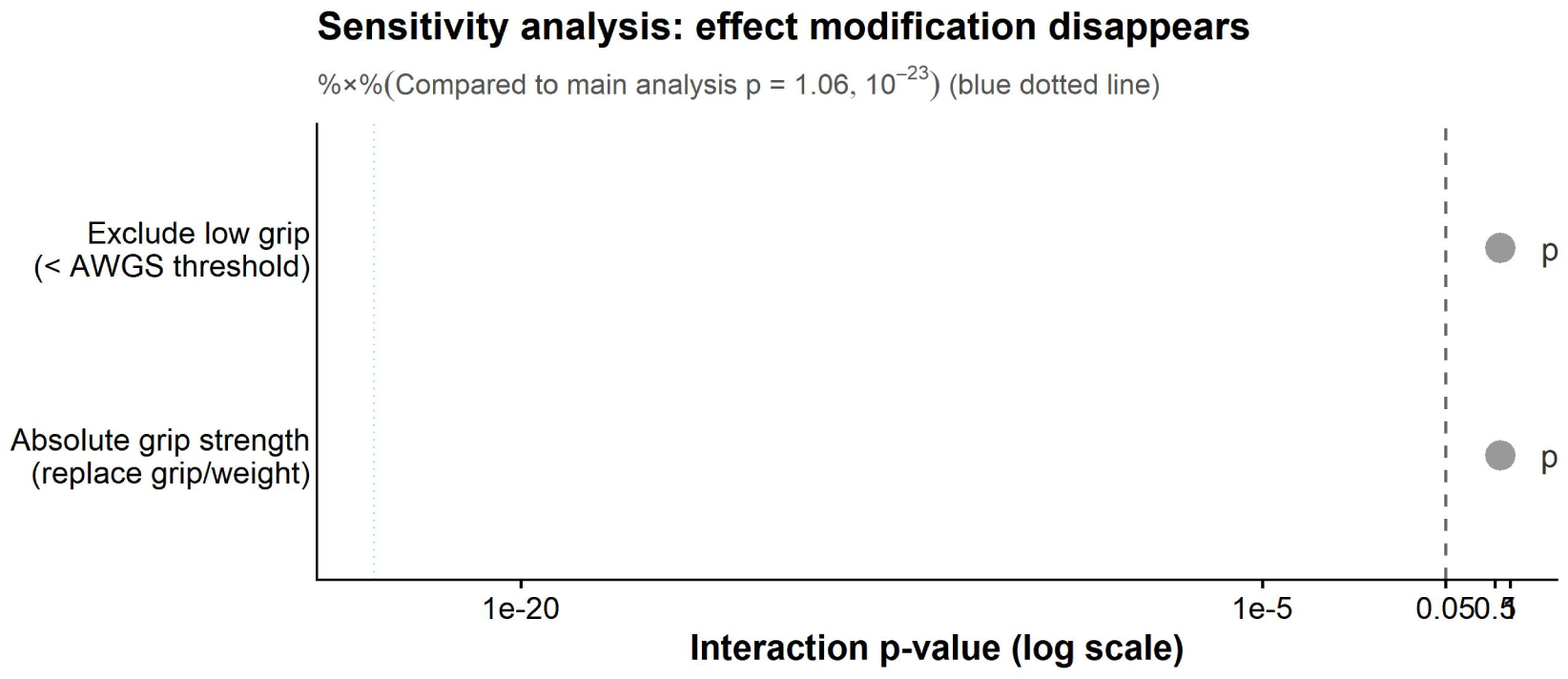
Sensitivity analyses: exclusion of participants with low grip strength and substitution of absolute grip strength for the grip/weight ratio. The x-axis displays the interaction p-value on a –log₁₀ scale. The dashed vertical line at p = 0.05 marks the conventional significance threshold. The dotted blue line marks the p-value from the main analysis (p = 1.06 × 10⁻²³). Grey points represent the p-values from two critical sensitivity analyses: (1) restricting the sample to participants with grip strength above the AWGS 2019 diagnostic thresholds (≥28 kg for men, ≥18 kg for women; p = 0.65), and (2) replacing the grip/weight ratio with absolute grip strength as the modifier (p = 0.63). Both sensitivity analyses show complete disappearance of the effect modification, supporting that the main finding reflects a genuine threshold effect of muscle function rather than circular reasoning or mathematical coupling driven by shared weight components.

**Figure S2.**
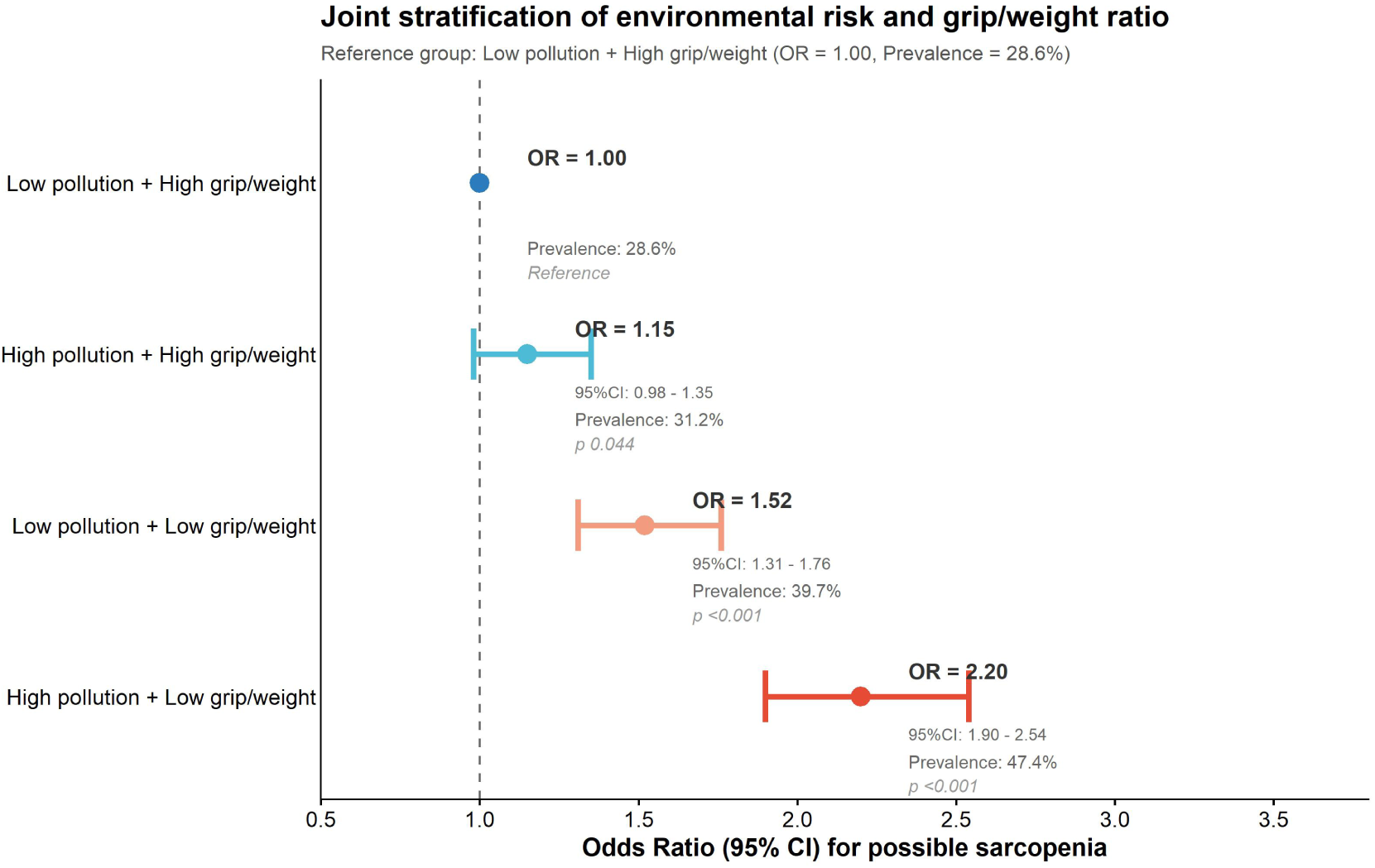
Joint stratification of environmental risk score and grip strength-to-weight ratio on possible sarcopenia risk. Odds ratios were estimated using logistic regression adjusted for age, sex, education, marital status, smoking status, alcohol consumption, multimorbidity, and BMI. The reference group is “Low environmental risk + High grip/weight ratio.” Environmental risk was dichotomized at the median of a composite environmental risk score incorporating air pollutants (PM₂.₅, PM₁₀, SO₂, NO₂, CO, O₃), climate factors, and green space exposure. The grip/weight ratio was dichotomized at the median. The highest sarcopenia risk was observed in the “High environmental risk + Low grip/weight ratio” group (OR = 2.20, 95% CI: 1.90–2.54, p < 0.001), indicating a synergistic association of adverse environmental conditions and low muscle function with sarcopenia prevalence. Error bars represent 95% confidence intervals.

**Table S1.**
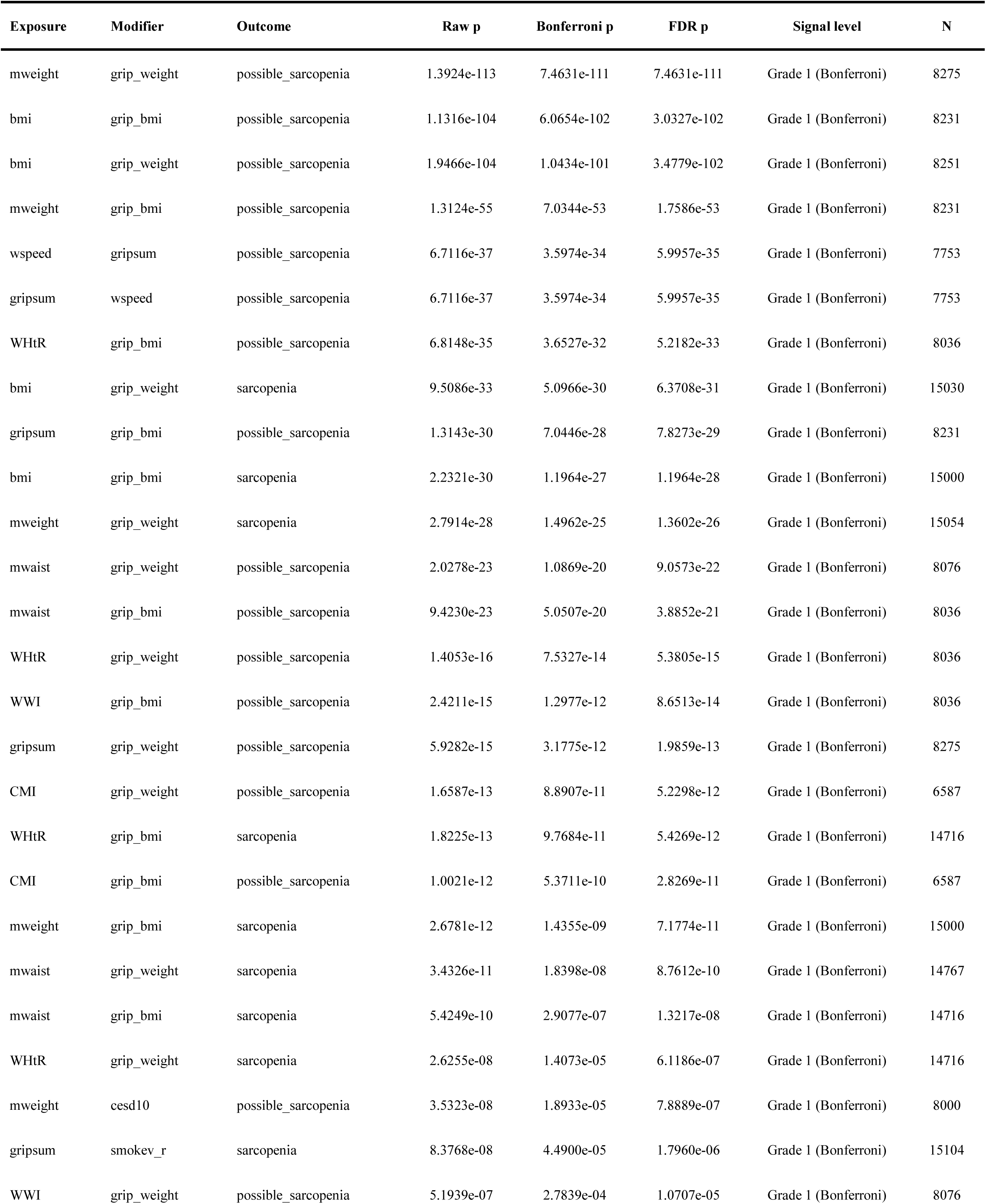
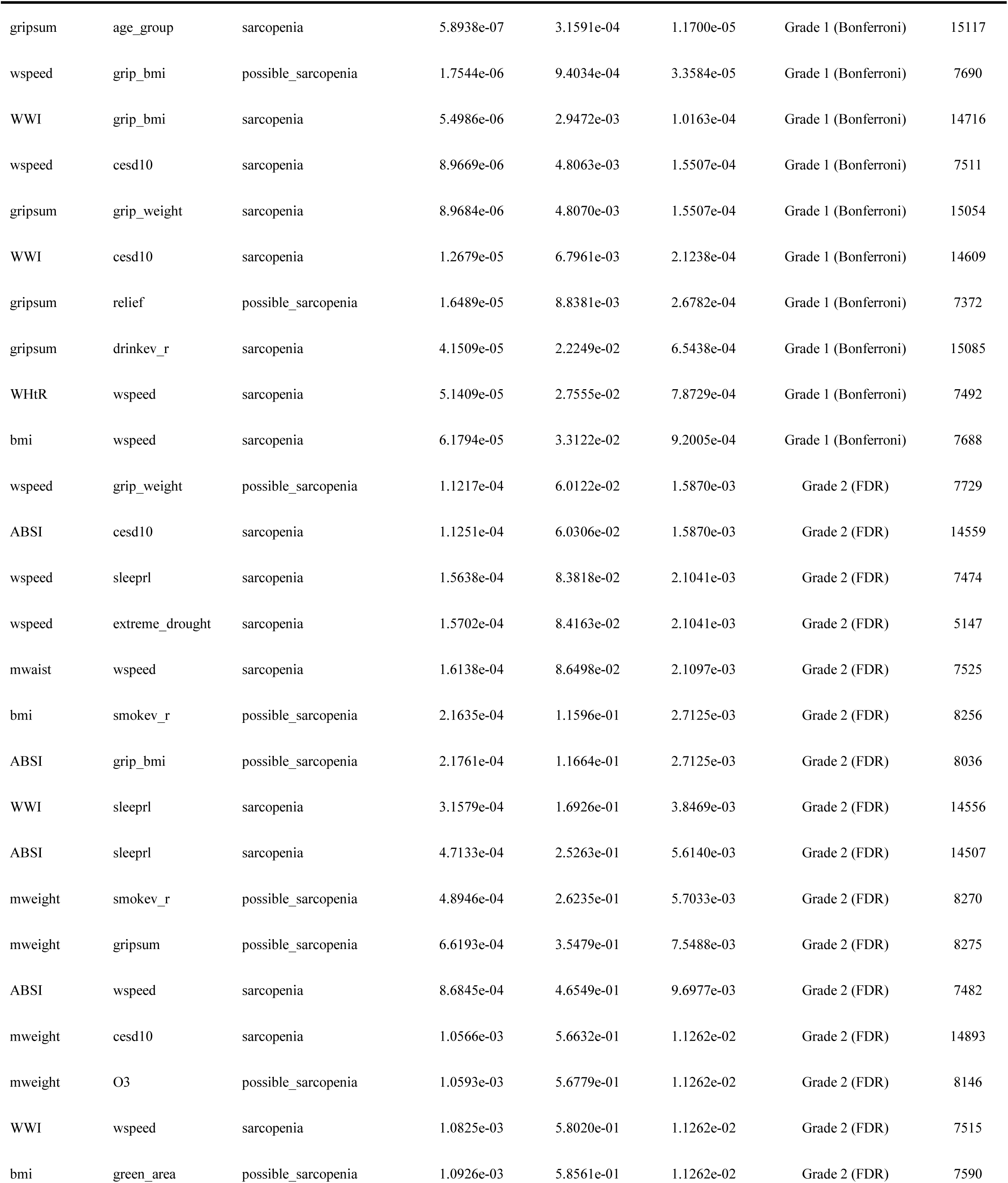
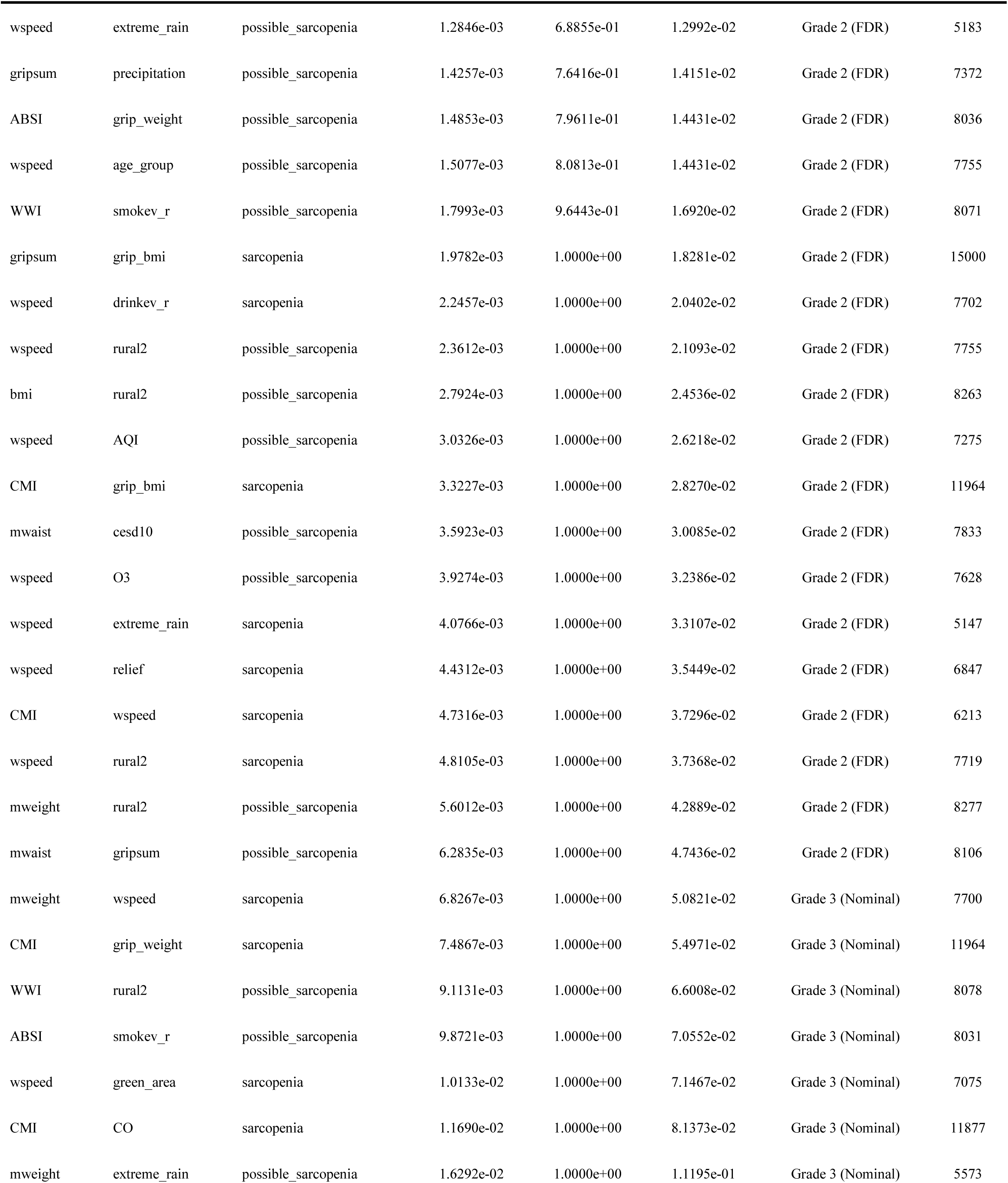
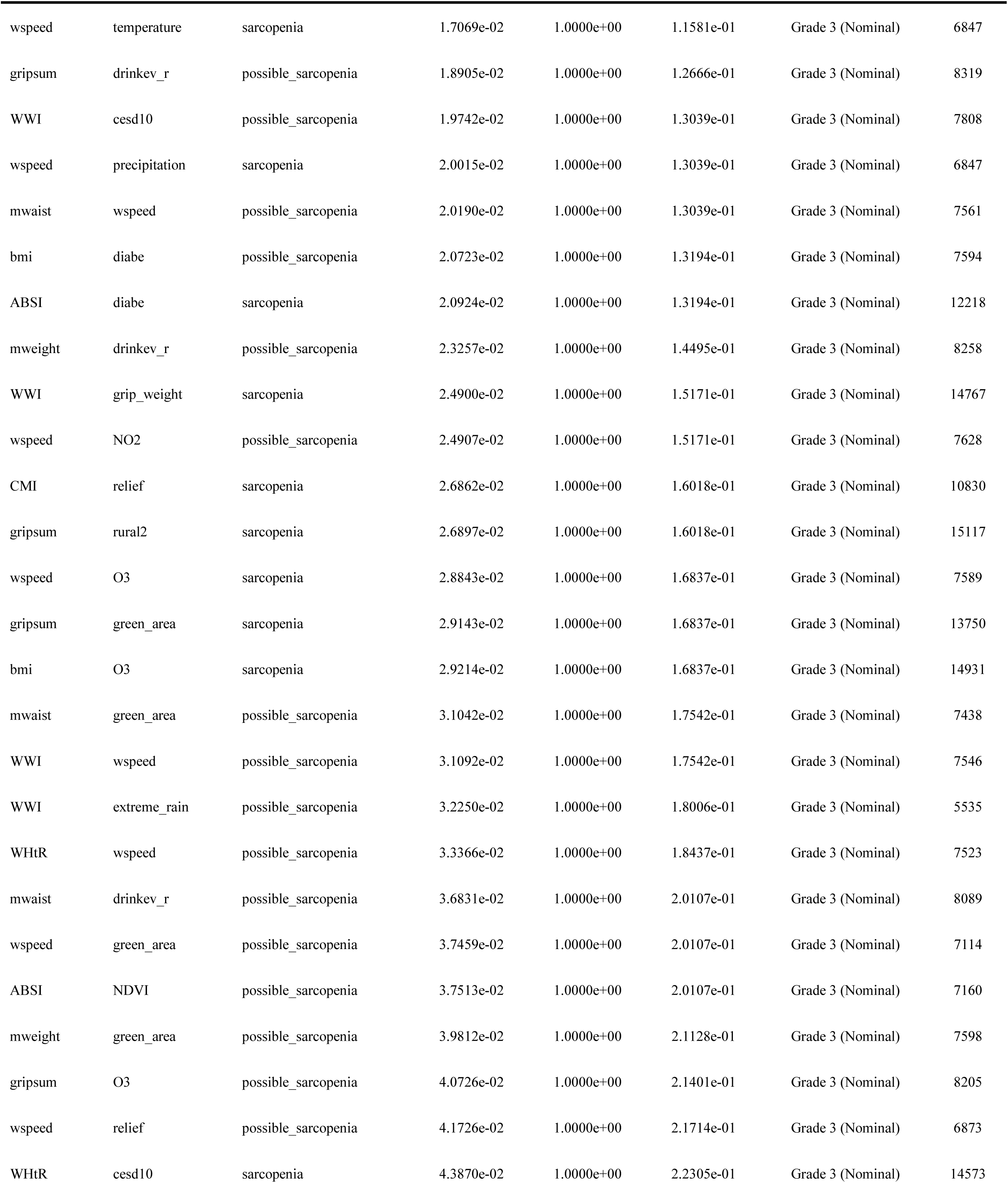
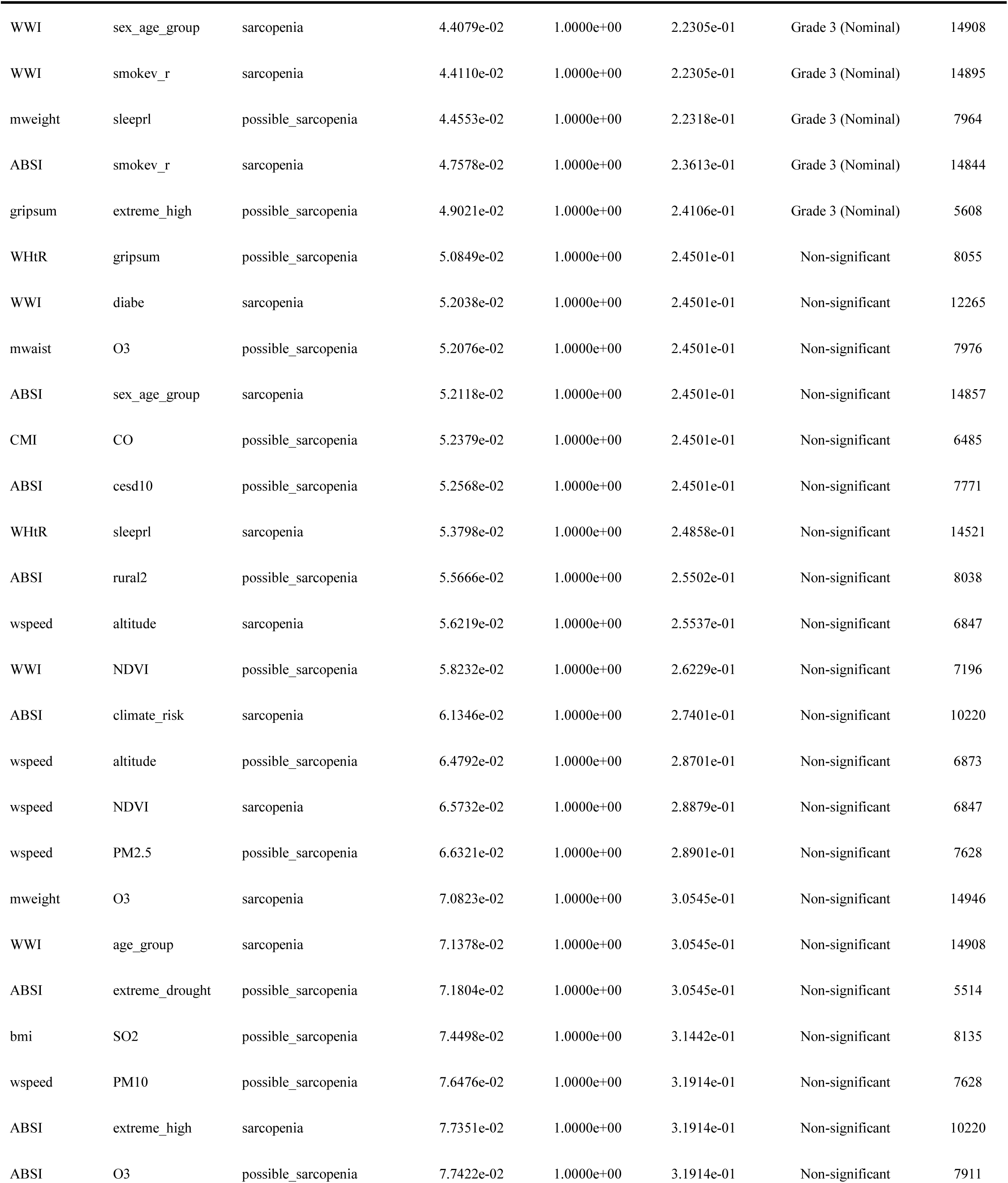
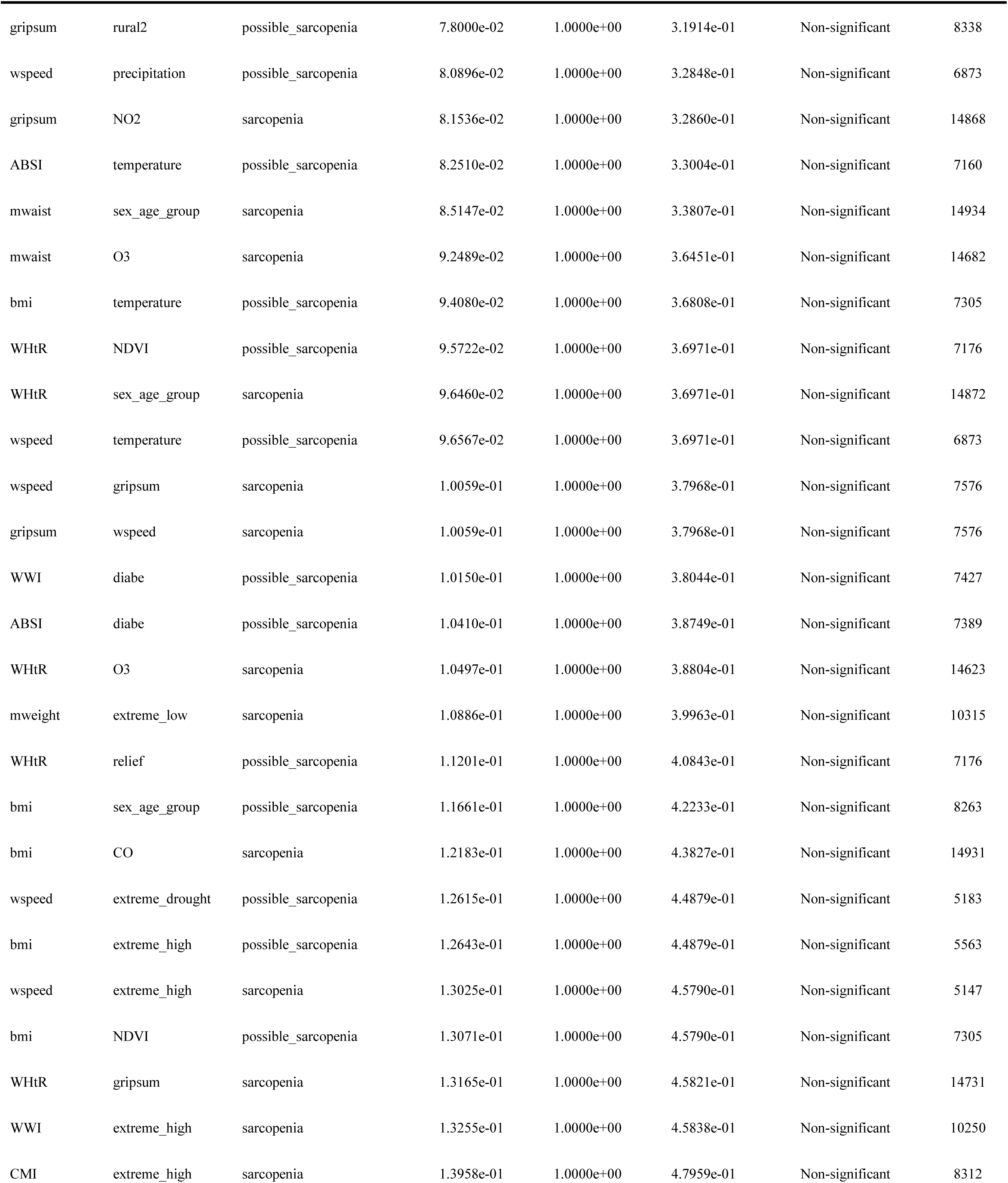
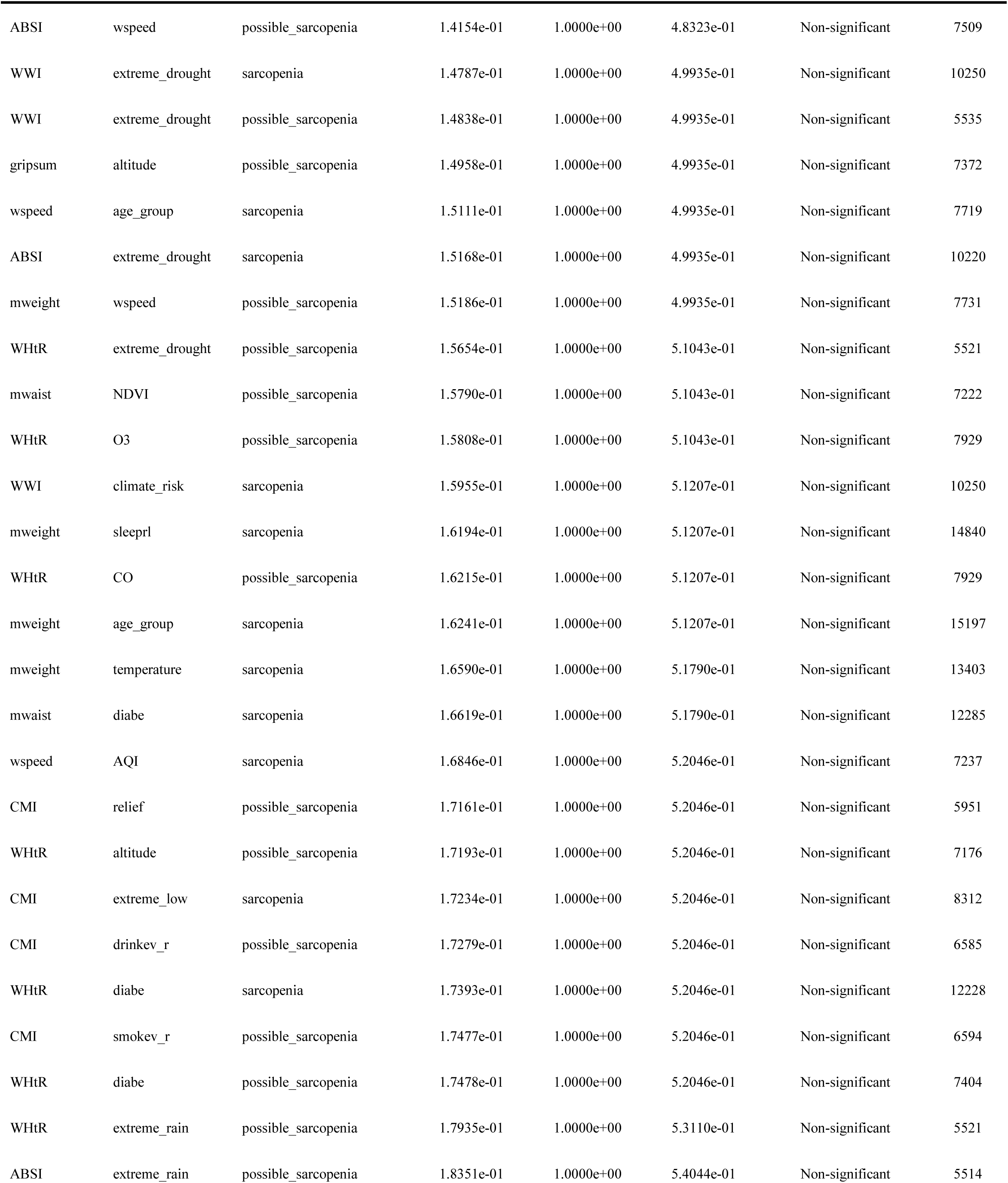
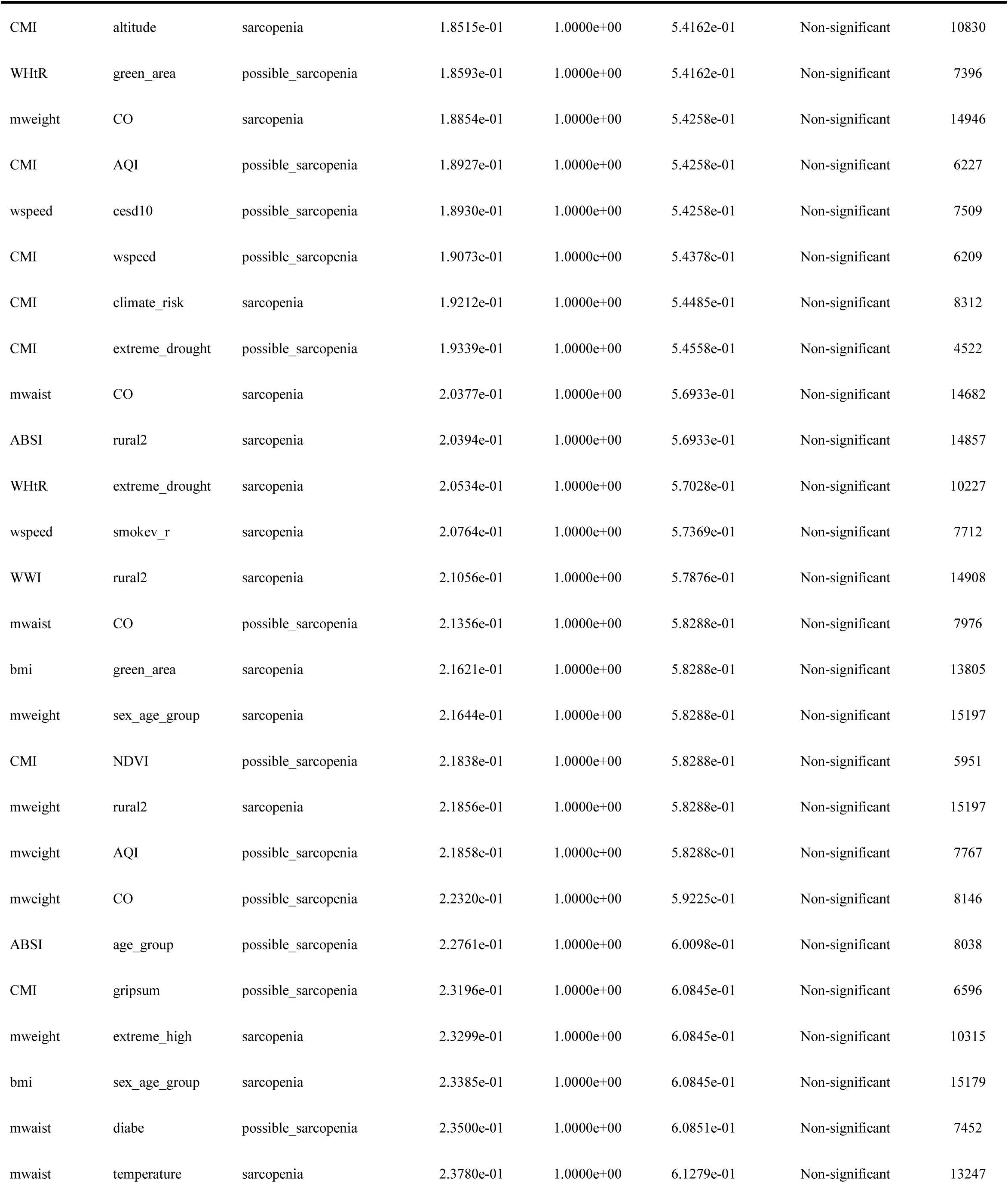
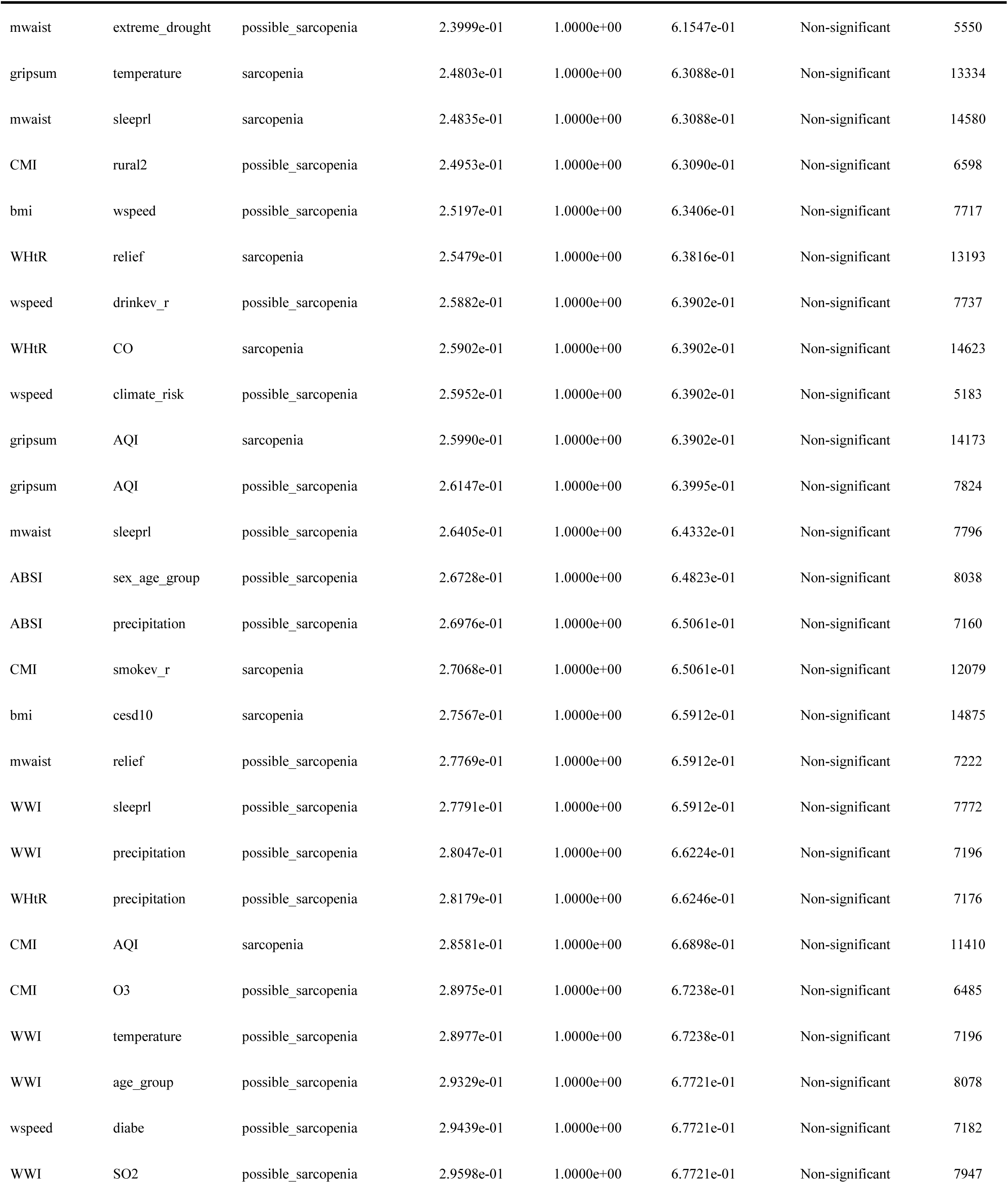
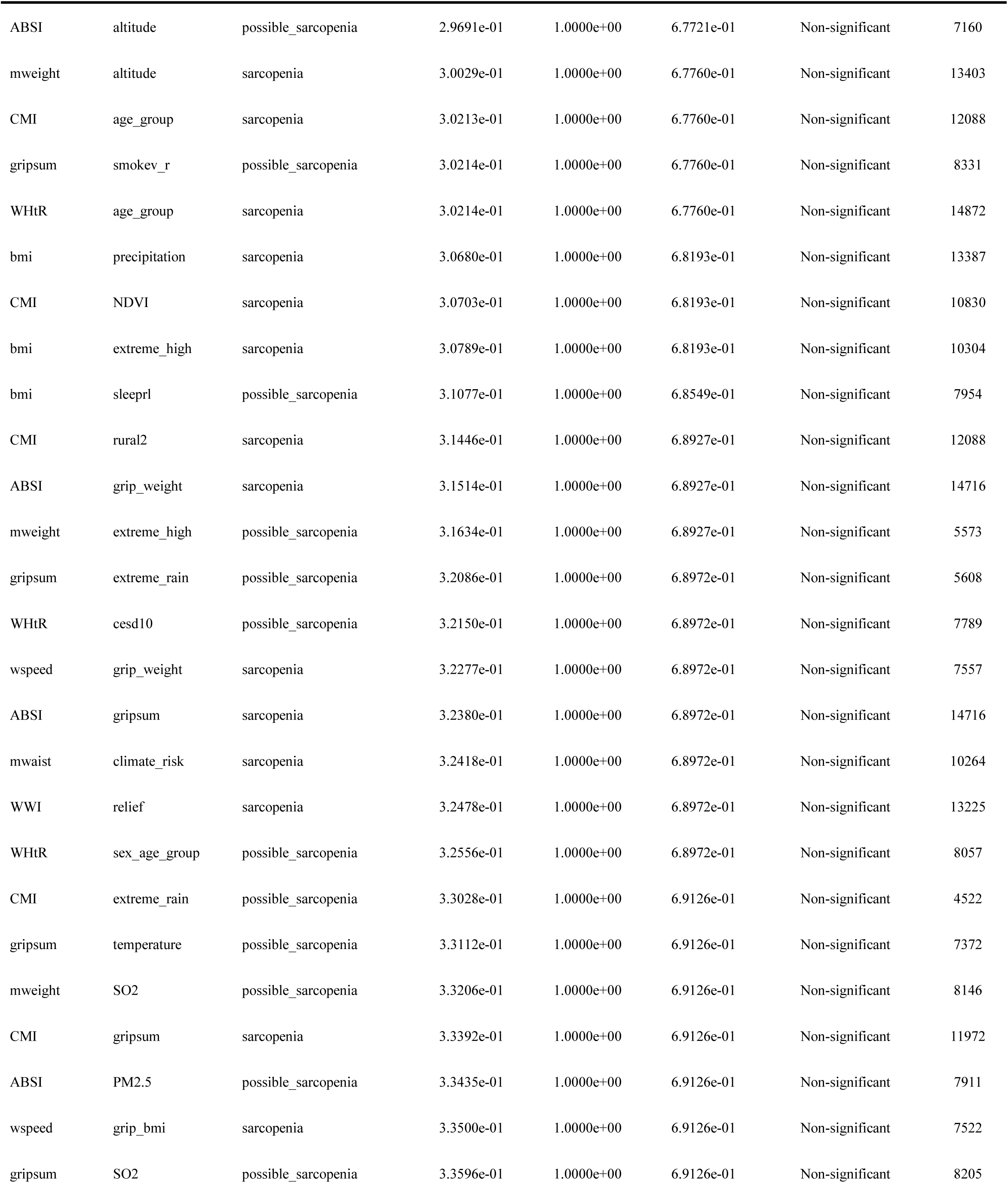
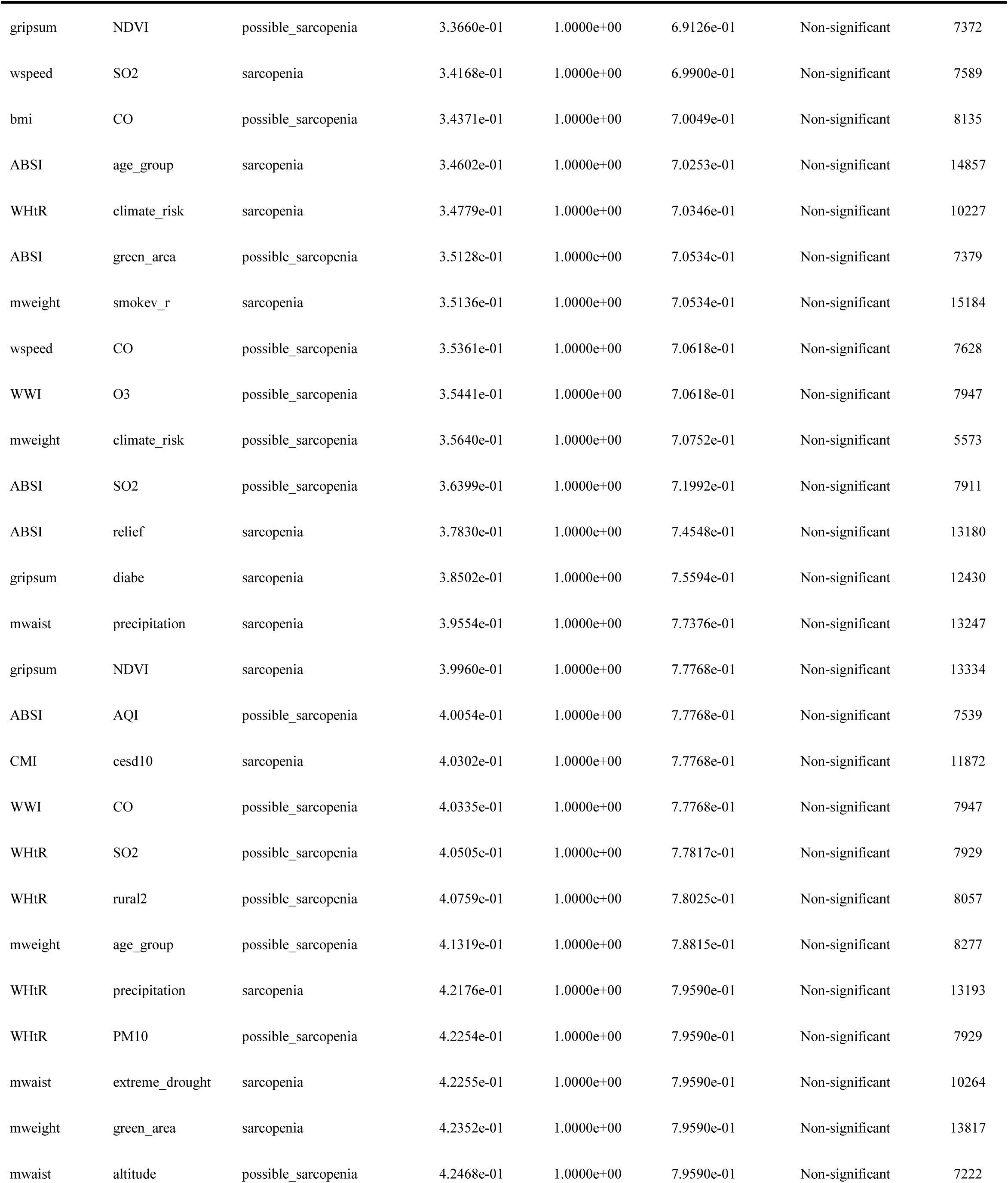
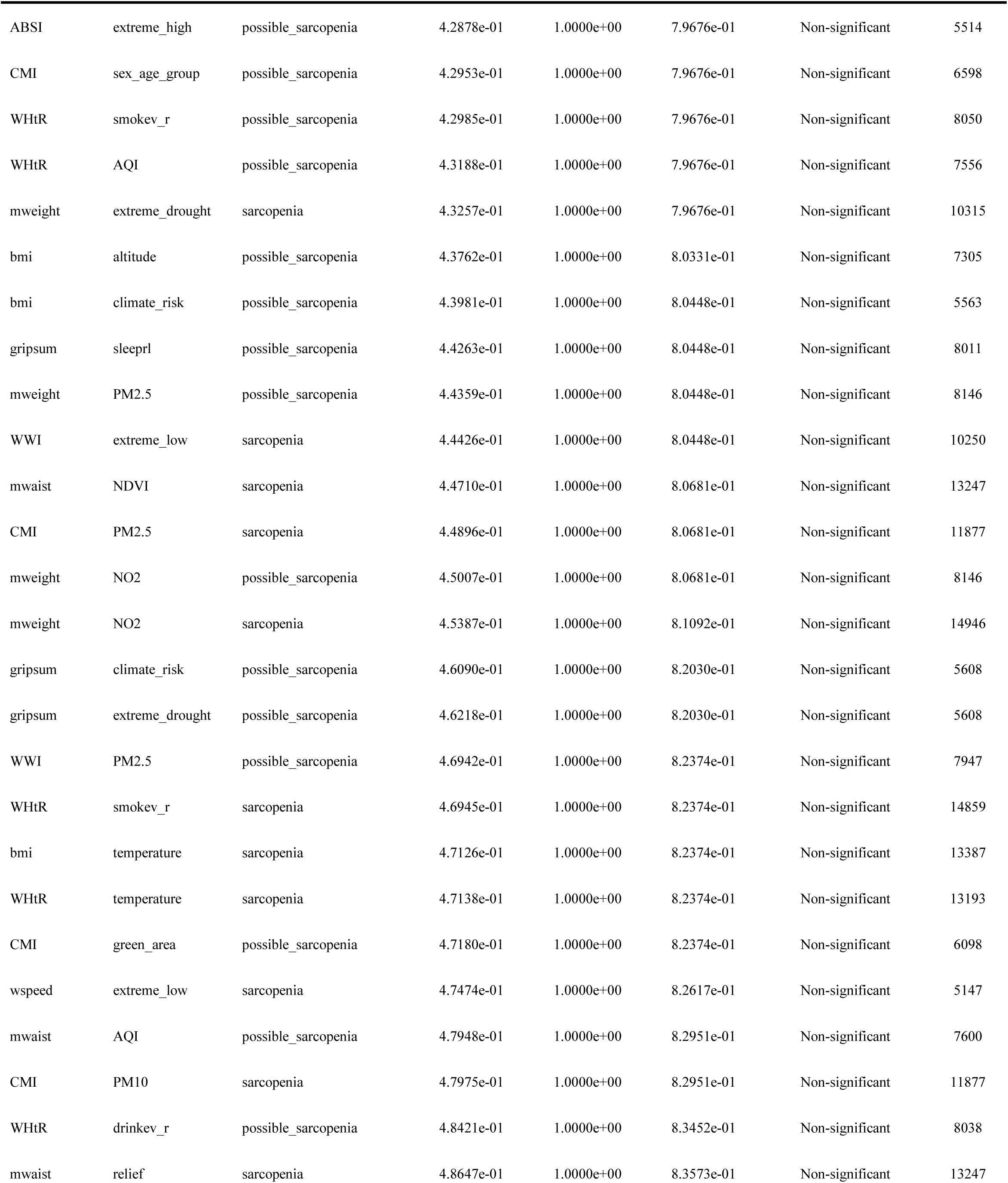
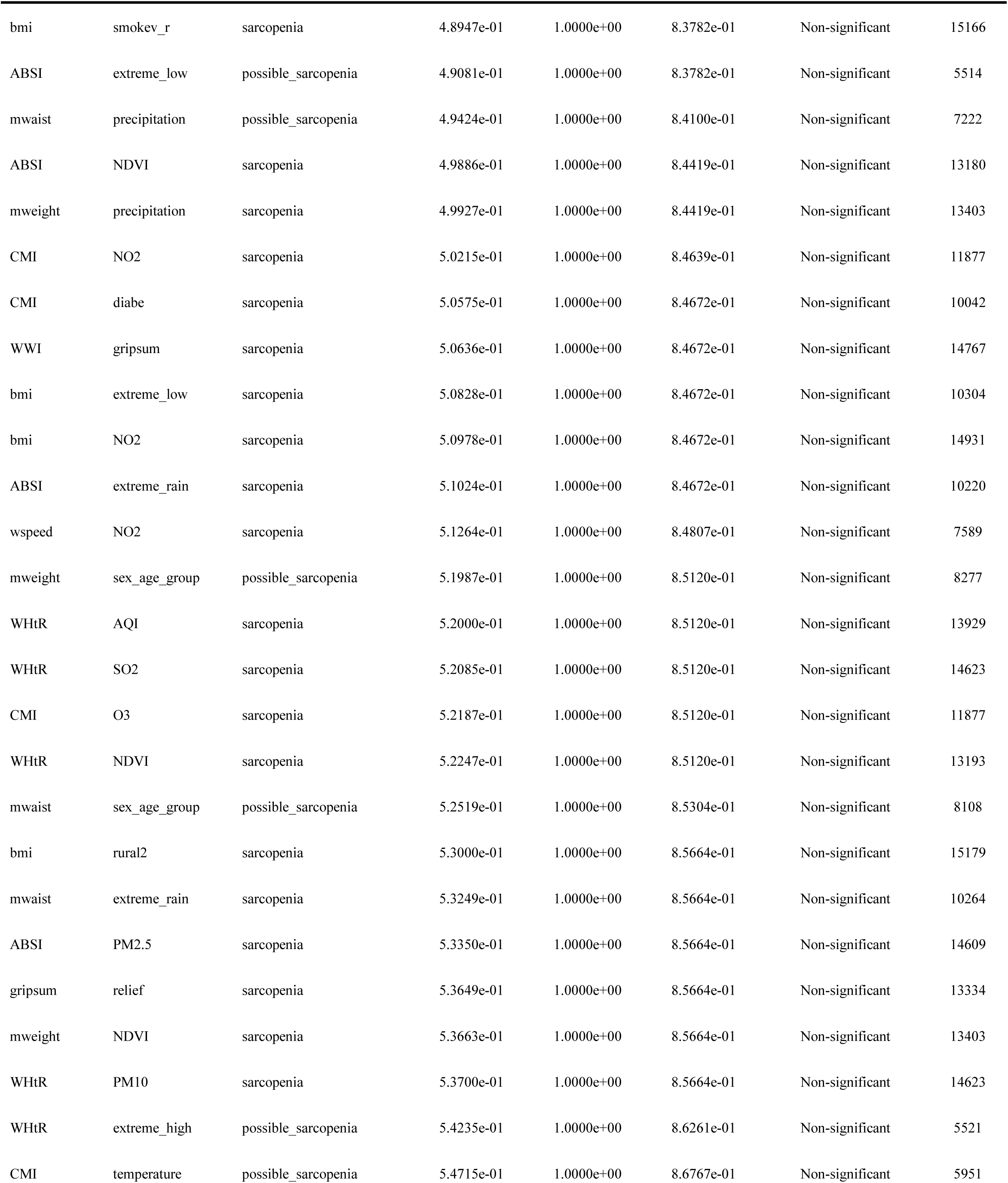
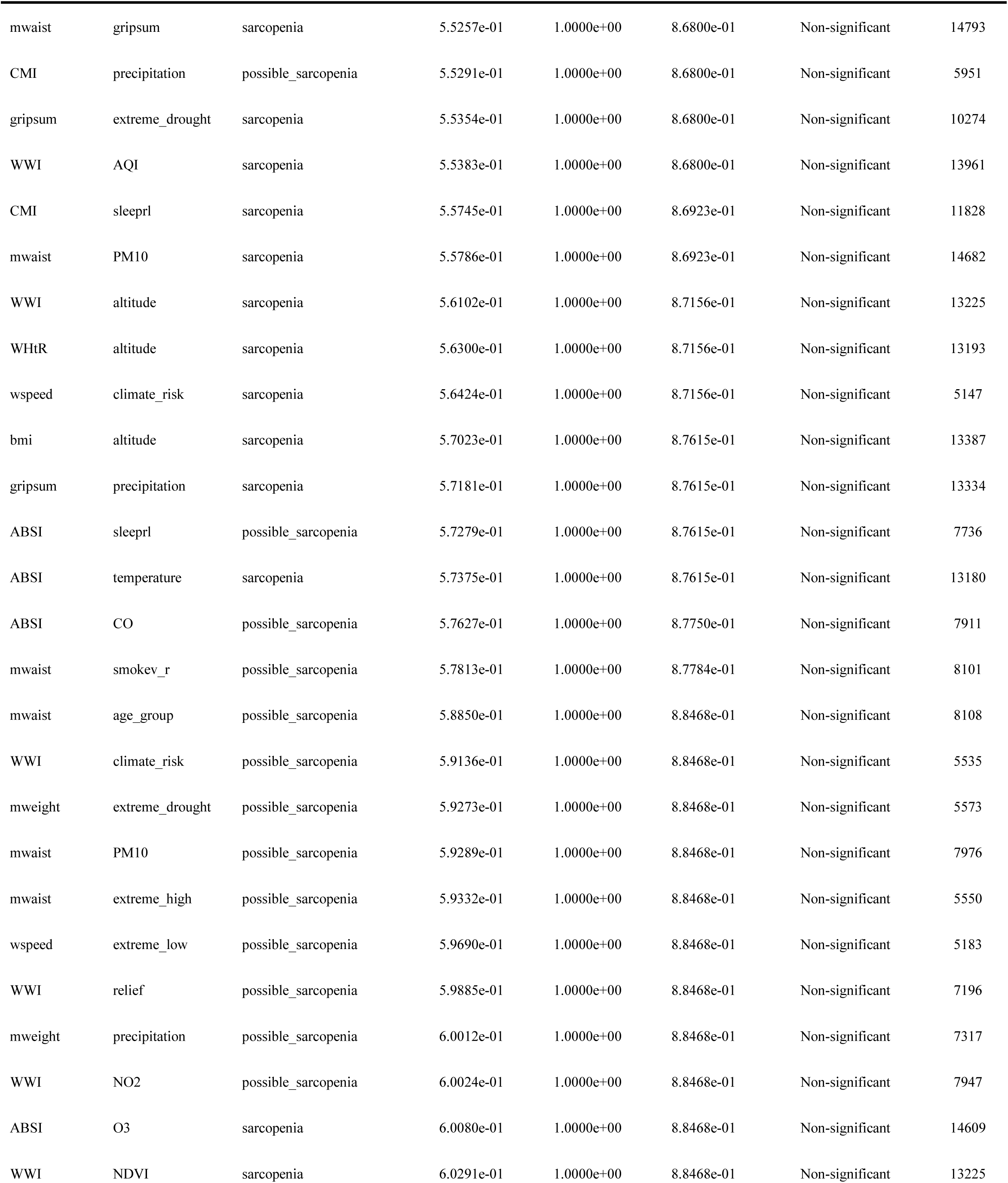
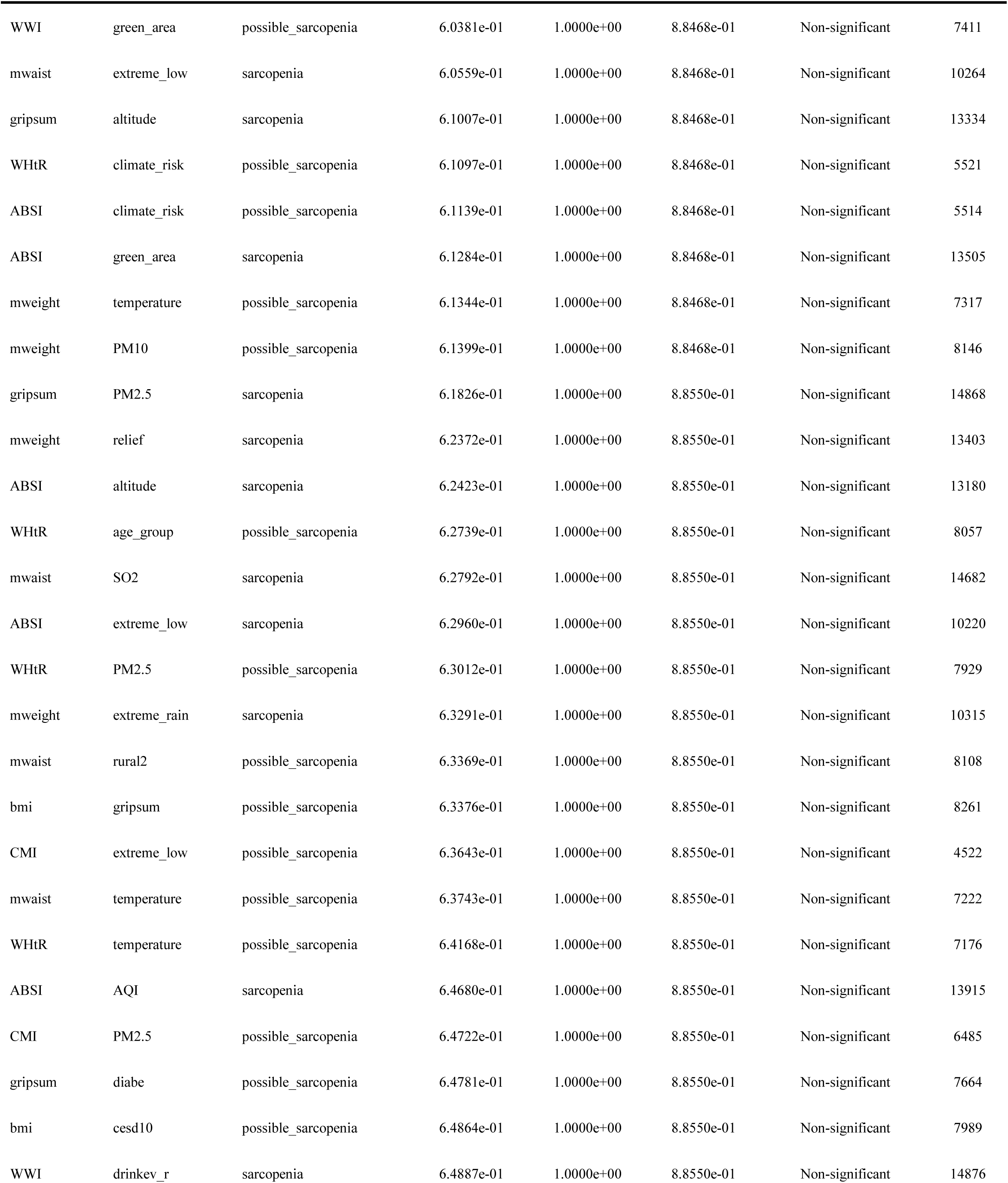
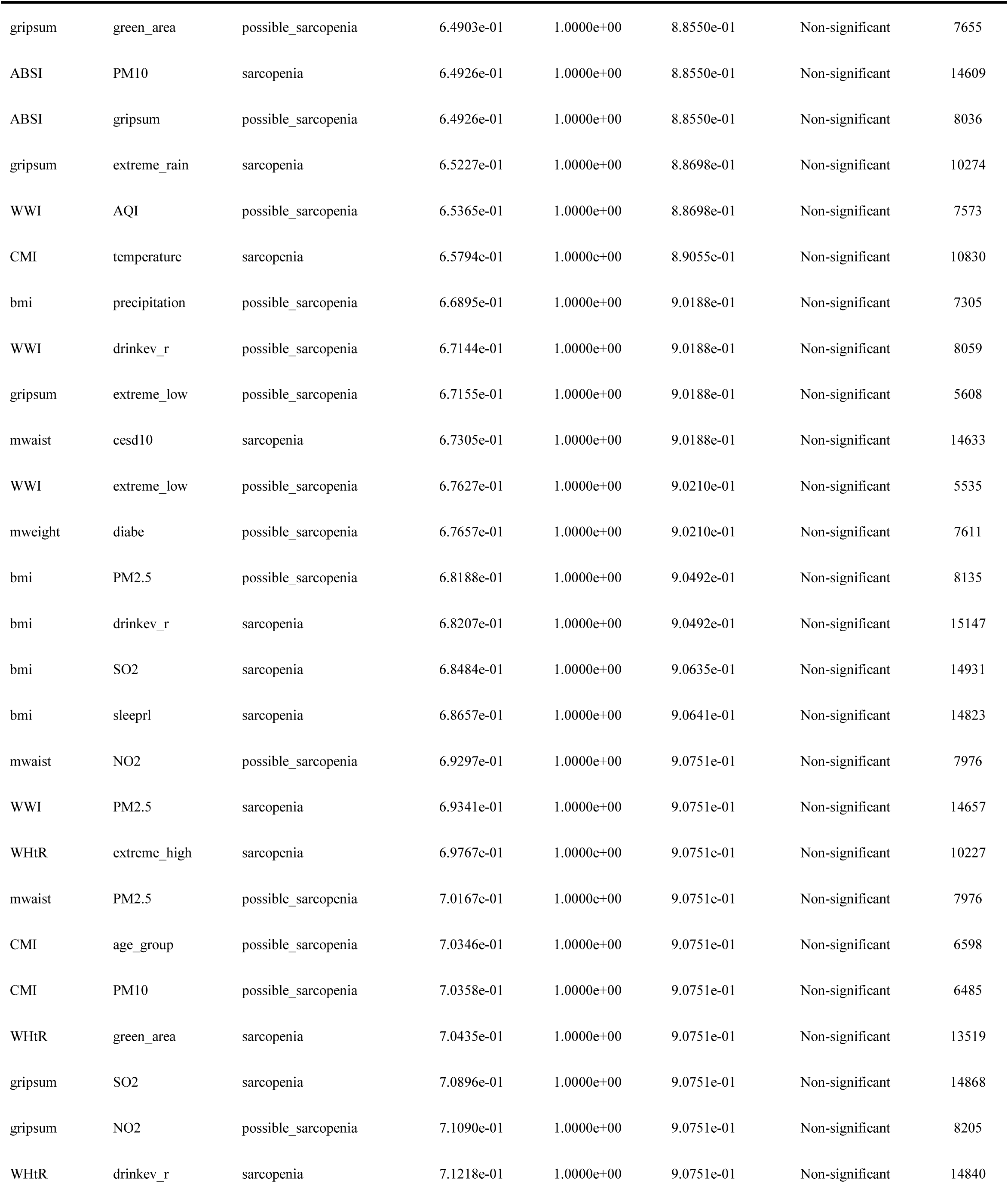
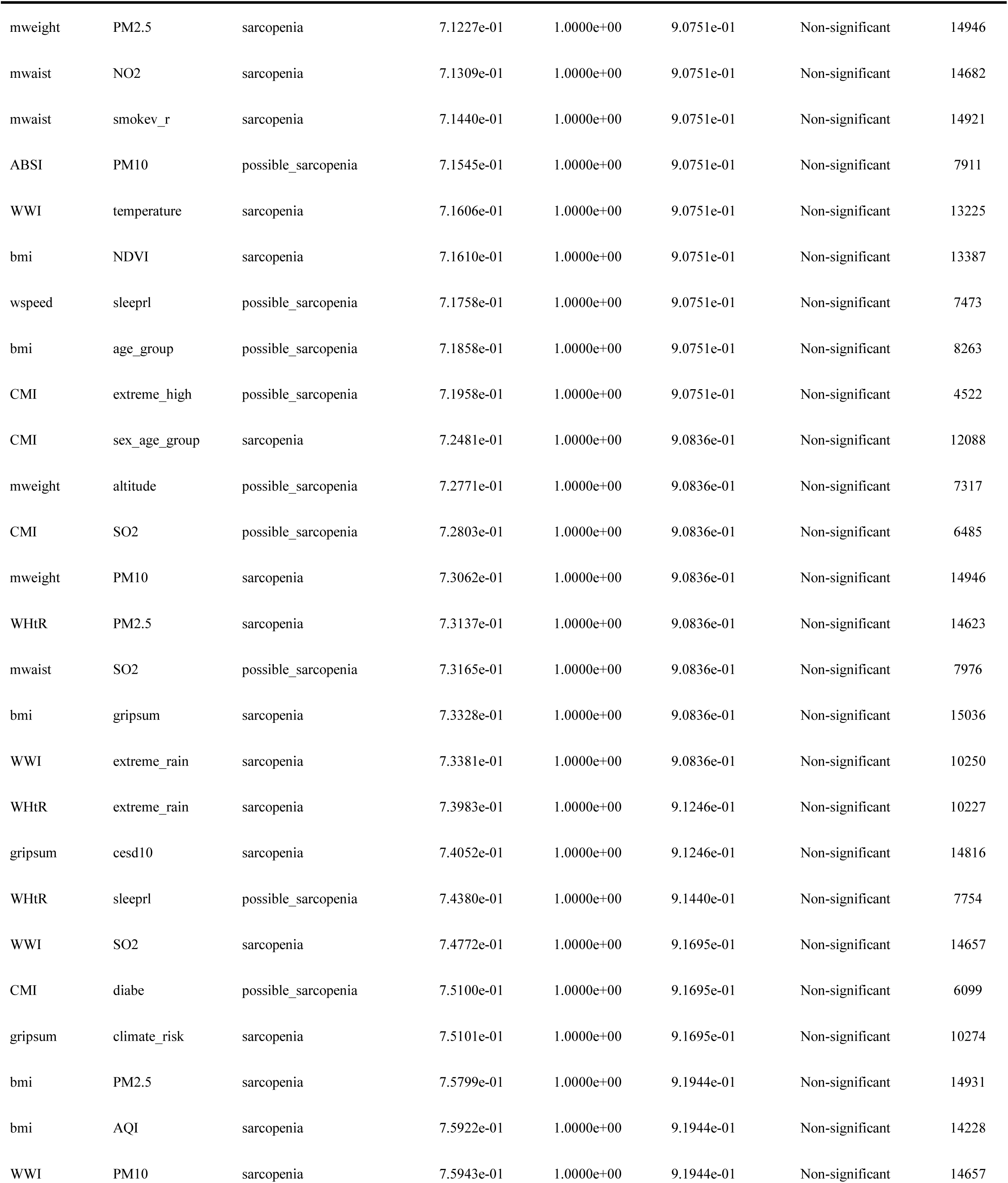
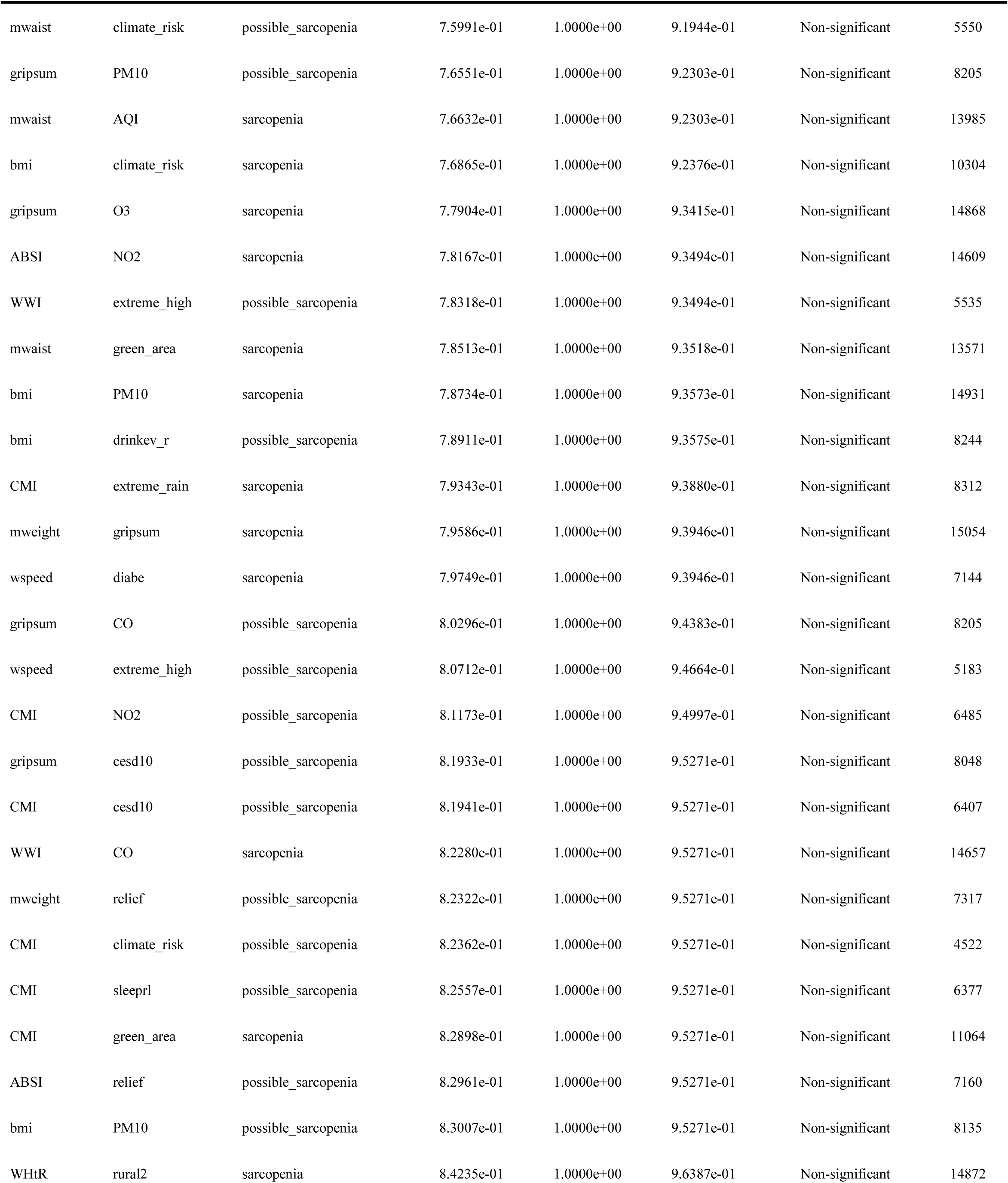
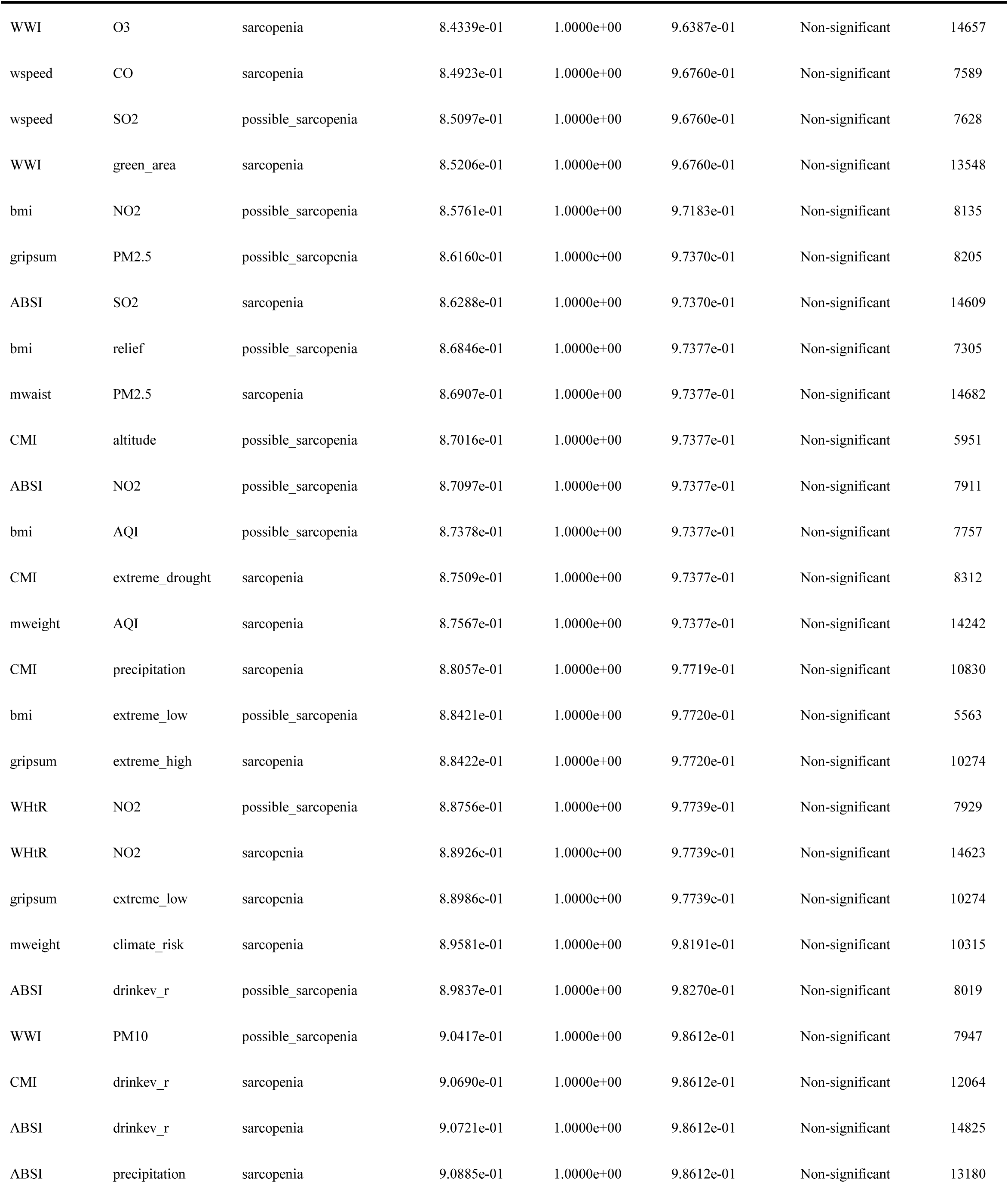
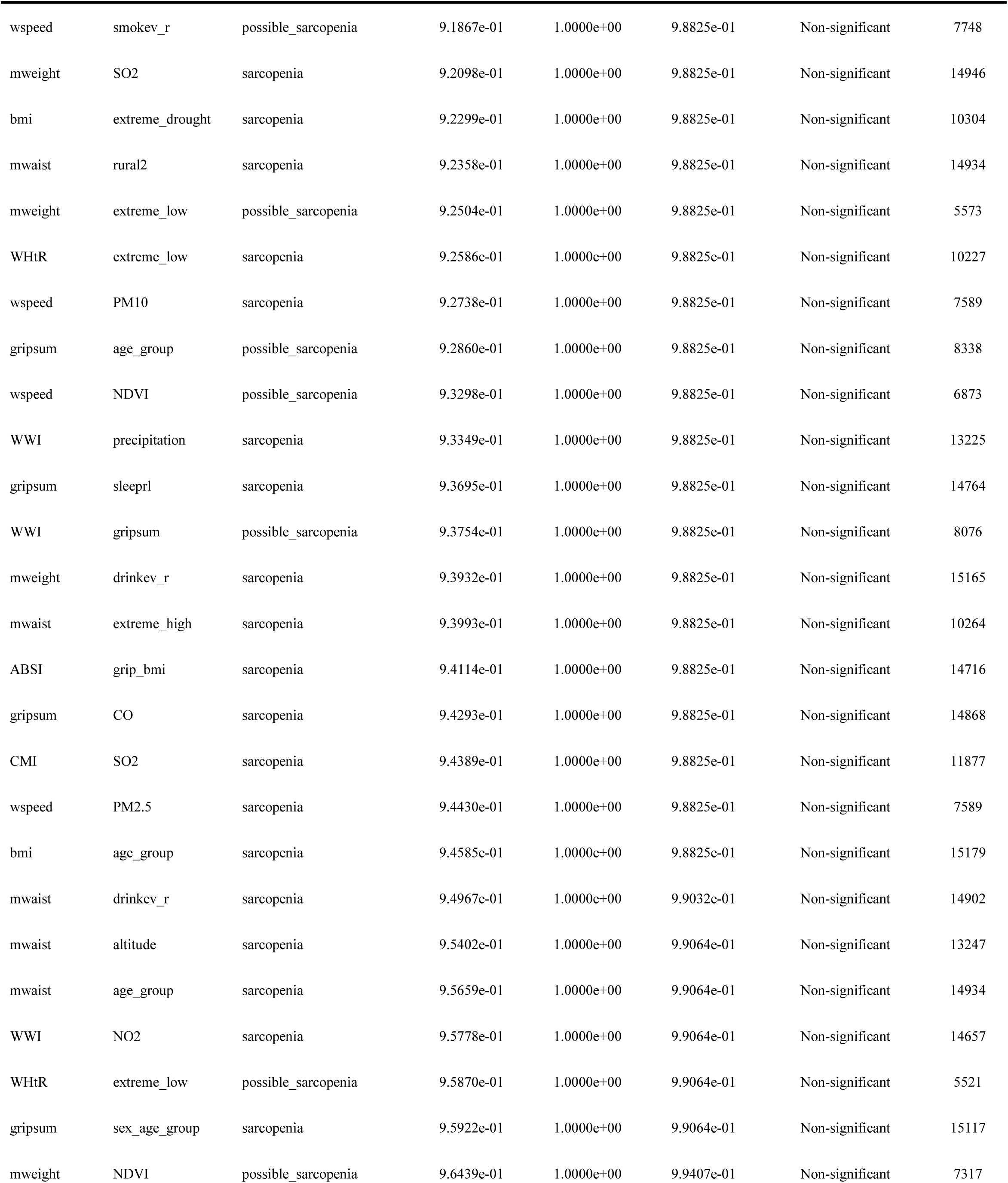
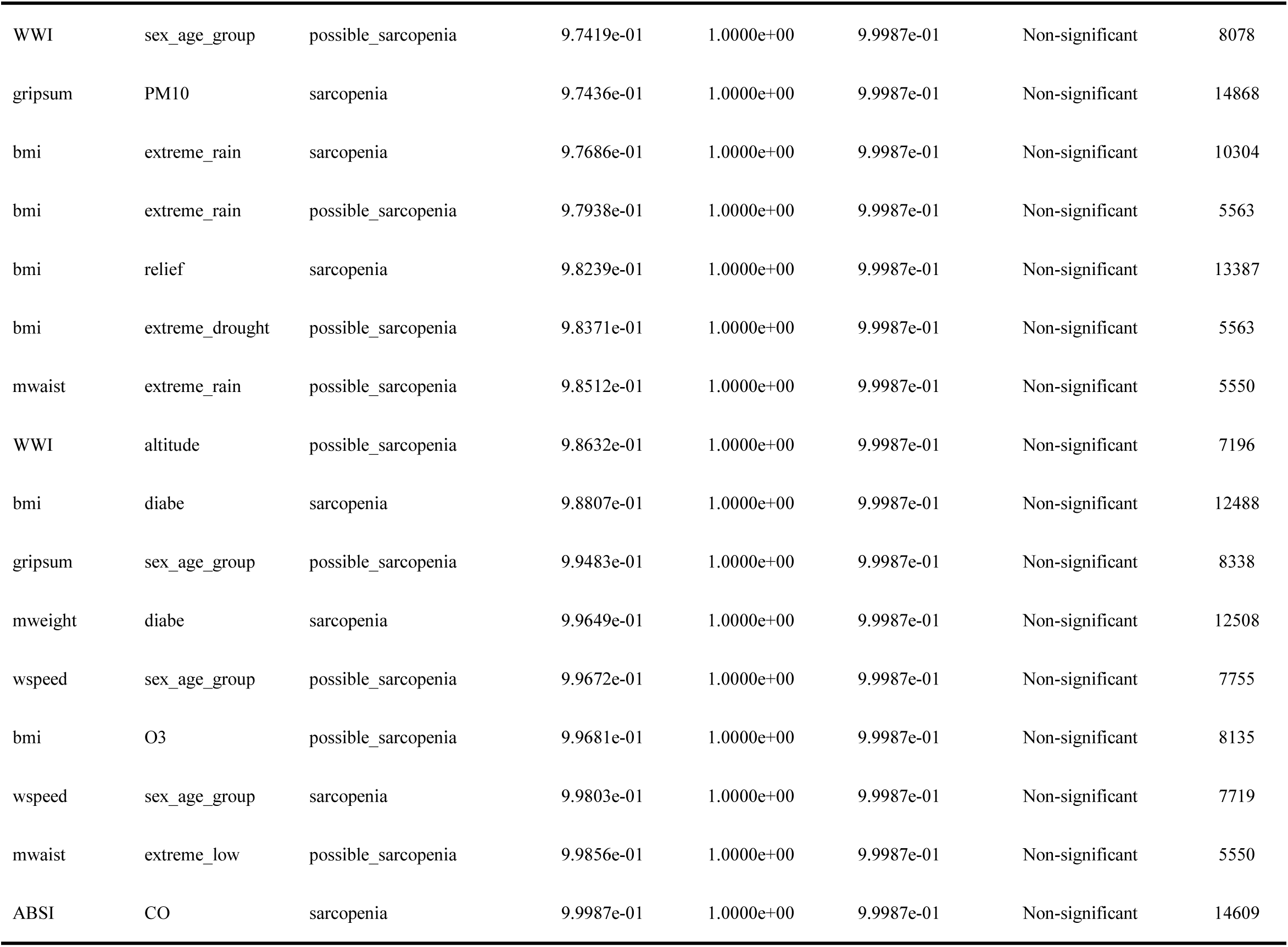
Complete results of the systematic interaction screen (N = 536 interaction terms). All interaction terms are sorted by raw p-value in ascending order. Bonferroni-corrected p-values (threshold = 9.3 × 10⁻⁵, corresponding to 0.05/536) and false discovery rate (FDR) q-values (Benjamini-Hochberg procedure) are reported. Signal classification: Grade 1, Bonferroni significant (p < 0.05 after Bonferroni correction); Grade 2, FDR significant (q < 0.05); Grade 3, nominal p < 0.05; Non-significant, all others. Of the 36 Grade 1 signals, 32 (89%) involved grip-related modifiers, covering all naïve anthropometric measures (waist circumference, BMI, WHtR, body weight) and derived adiposity indices. N denotes the number of observations included in each interaction model. All models were adjusted for age, sex, education, marital status, smoking status, alcohol consumption, and multimorbidity.

| Exposure | Modifier | Outcome | Raw p | Bonferroni p | FDR p | Signal level | N |
| --- | --- | --- | --- | --- | --- | --- | --- |
| mweight | grip_weight | possible_sarcopenia | 1.3924e-113 | 7.4631e-111 | 7.4631e-111 | Grade 1 (Bonferroni) | 8275 |
| bmi | grip_bmi | possible_sarcopenia | 1.1316e-104 | 6.0654e-102 | 3.0327e-102 | Grade 1 (Bonferroni) | 8231 |
| bmi | grip_weight | possible_sarcopenia | 1.9466e-104 | 1.0434e-101 | 3.4779e-102 | Grade 1 (Bonferroni) | 8251 |
| mweight | grip_bmi | possible_sarcopenia | 1.3124e-55 | 7.0344e-53 | 1.7586e-53 | Grade 1 (Bonferroni) | 8231 |
| wspeed | gripsum | possible_sarcopenia | 6.7116e-37 | 3.5974e-34 | 5.9957e-35 | Grade 1 (Bonferroni) | 7753 |
| gripsum | wspeed | possible_sarcopenia | 6.7116e-37 | 3.5974e-34 | 5.9957e-35 | Grade 1 (Bonferroni) | 7753 |
| WHtR | grip_bmi | possible_sarcopenia | 6.8148e-35 | 3.6527e-32 | 5.2182e-33 | Grade 1 (Bonferroni) | 8036 |
| bmi | grip_weight | sarcopenia | 9.5086e-33 | 5.0966e-30 | 6.3708e-31 | Grade 1 (Bonferroni) | 15030 |
| gripsum | grip_bmi | possible_sarcopenia | 1.3143e-30 | 7.0446e-28 | 7.8273e-29 | Grade 1 (Bonferroni) | 8231 |
| bmi | grip_bmi | sarcopenia | 2.2321e-30 | 1.1964e-27 | 1.1964e-28 | Grade 1 (Bonferroni) | 15000 |
| mweight | grip_weight | sarcopenia | 2.7914e-28 | 1.4962e-25 | 1.3602e-26 | Grade 1 (Bonferroni) | 15054 |
| mwaist | grip_weight | possible_sarcopenia | 2.0278e-23 | 1.0869e-20 | 9.0573e-22 | Grade 1 (Bonferroni) | 8076 |
| mwaist | grip_bmi | possible_sarcopenia | 9.4230e-23 | 5.0507e-20 | 3.8852e-21 | Grade 1 (Bonferroni) | 8036 |
| WHtR | grip_weight | possible_sarcopenia | 1.4053e-16 | 7.5327e-14 | 5.3805e-15 | Grade 1 (Bonferroni) | 8036 |
| WWI | grip_bmi | possible_sarcopenia | 2.4211e-15 | 1.2977e-12 | 8.6513e-14 | Grade 1 (Bonferroni) | 8036 |
| gripsum | grip_weight | possible_sarcopenia | 5.9282e-15 | 3.1775e-12 | 1.9859e-13 | Grade 1 (Bonferroni) | 8275 |
| CMI | grip_weight | possible_sarcopenia | 1.6587e-13 | 8.8907e-11 | 5.2298e-12 | Grade 1 (Bonferroni) | 6587 |
| WHtR | grip_bmi | sarcopenia | 1.8225e-13 | 9.7684e-11 | 5.4269e-12 | Grade 1 (Bonferroni) | 14716 |
| CMI | grip_bmi | possible_sarcopenia | 1.0021e-12 | 5.3711e-10 | 2.8269e-11 | Grade 1 (Bonferroni) | 6587 |
| mweight | grip_bmi | sarcopenia | 2.6781e-12 | 1.4355e-09 | 7.1774e-11 | Grade 1 (Bonferroni) | 15000 |
| mwaist | grip_weight | sarcopenia | 3.4326e-11 | 1.8398e-08 | 8.7612e-10 | Grade 1 (Bonferroni) | 14767 |
| mwaist | grip_bmi | sarcopenia | 5.4249e-10 | 2.9077e-07 | 1.3217e-08 | Grade 1 (Bonferroni) | 14716 |
| WHtR | grip_weight | sarcopenia | 2.6255e-08 | 1.4073e-05 | 6.1186e-07 | Grade 1 (Bonferroni) | 14716 |
| mweight | cesd10 | possible_sarcopenia | 3.5323e-08 | 1.8933e-05 | 7.8889e-07 | Grade 1 (Bonferroni) | 8000 |
| gripsum | smokev_r | sarcopenia | 8.3768e-08 | 4.4900e-05 | 1.7960e-06 | Grade 1 (Bonferroni) | 15104 |
| WWI | grip_weight | possible_sarcopenia | 5.1939e-07 | 2.7839e-04 | 1.0707e-05 | Grade 1 (Bonferroni) | 8076 |
| gripsum | age_group | sarcopenia | 5.8938e-07 | 3.1591e-04 | 1.1700e-05 | Grade 1 (Bonferroni) | 15117 |
| wspeed | grip_bmi | possible_sarcopenia | 1.7544e-06 | 9.4034e-04 | 3.3584e-05 | Grade 1 (Bonferroni) | 7690 |
| WWI | grip_bmi | sarcopenia | 5.4986e-06 | 2.9472e-03 | 1.0163e-04 | Grade 1 (Bonferroni) | 14716 |
| wspeed | cesd10 | sarcopenia | 8.9669e-06 | 4.8063e-03 | 1.5507e-04 | Grade 1 (Bonferroni) | 7511 |
| gripsum | grip_weight | sarcopenia | 8.9684e-06 | 4.8070e-03 | 1.5507e-04 | Grade 1 (Bonferroni) | 15054 |
| WWI | cesd10 | sarcopenia | 1.2679e-05 | 6.7961e-03 | 2.1238e-04 | Grade 1 (Bonferroni) | 14609 |
| gripsum | relief | possible_sarcopenia | 1.6489e-05 | 8.8381e-03 | 2.6782e-04 | Grade 1 (Bonferroni) | 7372 |
| gripsum | drinkev_r | sarcopenia | 4.1509e-05 | 2.2249e-02 | 6.5438e-04 | Grade 1 (Bonferroni) | 15085 |
| WHtR | wspeed | sarcopenia | 5.1409e-05 | 2.7555e-02 | 7.8729e-04 | Grade 1 (Bonferroni) | 7492 |
| bmi | wspeed | sarcopenia | 6.1794e-05 | 3.3122e-02 | 9.2005e-04 | Grade 1 (Bonferroni) | 7688 |
| wspeed | grip_weight | possible_sarcopenia | 1.1217e-04 | 6.0122e-02 | 1.5870e-03 | Grade 2 (FDR) | 7729 |
| ABSI | cesd10 | sarcopenia | 1.1251e-04 | 6.0306e-02 | 1.5870e-03 | Grade 2 (FDR) | 14559 |
| wspeed | sleeprl | sarcopenia | 1.5638e-04 | 8.3818e-02 | 2.1041e-03 | Grade 2 (FDR) | 7474 |
| wspeed | extreme_drought | sarcopenia | 1.5702e-04 | 8.4163e-02 | 2.1041e-03 | Grade 2 (FDR) | 5147 |
| mwaist | wspeed | sarcopenia | 1.6138e-04 | 8.6498e-02 | 2.1097e-03 | Grade 2 (FDR) | 7525 |
| bmi | smokev_r | possible_sarcopenia | 2.1635e-04 | 1.1596e-01 | 2.7125e-03 | Grade 2 (FDR) | 8256 |
| ABSI | grip_bmi | possible_sarcopenia | 2.1761e-04 | 1.1664e-01 | 2.7125e-03 | Grade 2 (FDR) | 8036 |
| WWI | sleeprl | sarcopenia | 3.1579e-04 | 1.6926e-01 | 3.8469e-03 | Grade 2 (FDR) | 14556 |
| ABSI | sleeprl | sarcopenia | 4.7133e-04 | 2.5263e-01 | 5.6140e-03 | Grade 2 (FDR) | 14507 |
| mweight | smokev_r | possible_sarcopenia | 4.8946e-04 | 2.6235e-01 | 5.7033e-03 | Grade 2 (FDR) | 8270 |
| mweight | gripsum | possible_sarcopenia | 6.6193e-04 | 3.5479e-01 | 7.5488e-03 | Grade 2 (FDR) | 8275 |
| ABSI | wspeed | sarcopenia | 8.6845e-04 | 4.6549e-01 | 9.6977e-03 | Grade 2 (FDR) | 7482 |
| mweight | cesd10 | sarcopenia | 1.0566e-03 | 5.6632e-01 | 1.1262e-02 | Grade 2 (FDR) | 14893 |
| mweight | O3 | possible_sarcopenia | 1.0593e-03 | 5.6779e-01 | 1.1262e-02 | Grade 2 (FDR) | 8146 |
| WWI | wspeed | sarcopenia | 1.0825e-03 | 5.8020e-01 | 1.1262e-02 | Grade 2 (FDR) | 7515 |
| bmi | green_area | possible_sarcopenia | 1.0926e-03 | 5.8561e-01 | 1.1262e-02 | Grade 2 (FDR) | 7590 |
| wspeed | extreme_rain | possible_sarcopenia | 1.2846e-03 | 6.8855e-01 | 1.2992e-02 | Grade 2 (FDR) | 5183 |
| gripsum | precipitation | possible_sarcopenia | 1.4257e-03 | 7.6416e-01 | 1.4151e-02 | Grade 2 (FDR) | 7372 |
| ABSI | grip_weight | possible_sarcopenia | 1.4853e-03 | 7.9611e-01 | 1.4431e-02 | Grade 2 (FDR) | 8036 |
| wspeed | age_group | possible_sarcopenia | 1.5077e-03 | 8.0813e-01 | 1.4431e-02 | Grade 2 (FDR) | 7755 |
| WWI | smokev_r | possible_sarcopenia | 1.7993e-03 | 9.6443e-01 | 1.6920e-02 | Grade 2 (FDR) | 8071 |
| gripsum | grip_bmi | sarcopenia | 1.9782e-03 | 1.0000e+00 | 1.8281e-02 | Grade 2 (FDR) | 15000 |
| wspeed | drinkev_r | sarcopenia | 2.2457e-03 | 1.0000e+00 | 2.0402e-02 | Grade 2 (FDR) | 7702 |
| wspeed | rural2 | possible_sarcopenia | 2.3612e-03 | 1.0000e+00 | 2.1093e-02 | Grade 2 (FDR) | 7755 |
| bmi | rural2 | possible_sarcopenia | 2.7924e-03 | 1.0000e+00 | 2.4536e-02 | Grade 2 (FDR) | 8263 |
| wspeed | AQI | possible_sarcopenia | 3.0326e-03 | 1.0000e+00 | 2.6218e-02 | Grade 2 (FDR) | 7275 |
| CMI | grip_bmi | sarcopenia | 3.3227e-03 | 1.0000e+00 | 2.8270e-02 | Grade 2 (FDR) | 11964 |
| mwaist | cesd10 | possible_sarcopenia | 3.5923e-03 | 1.0000e+00 | 3.0085e-02 | Grade 2 (FDR) | 7833 |
| wspeed | O3 | possible_sarcopenia | 3.9274e-03 | 1.0000e+00 | 3.2386e-02 | Grade 2 (FDR) | 7628 |
| wspeed | extreme_rain | sarcopenia | 4.0766e-03 | 1.0000e+00 | 3.3107e-02 | Grade 2 (FDR) | 5147 |
| wspeed | relief | sarcopenia | 4.4312e-03 | 1.0000e+00 | 3.5449e-02 | Grade 2 (FDR) | 6847 |
| CMI | wspeed | sarcopenia | 4.7316e-03 | 1.0000e+00 | 3.7296e-02 | Grade 2 (FDR) | 6213 |
| wspeed | rural2 | sarcopenia | 4.8105e-03 | 1.0000e+00 | 3.7368e-02 | Grade 2 (FDR) | 7719 |
| mweight | rural2 | possible_sarcopenia | 5.6012e-03 | 1.0000e+00 | 4.2889e-02 | Grade 2 (FDR) | 8277 |
| mwaist | gripsum | possible_sarcopenia | 6.2835e-03 | 1.0000e+00 | 4.7436e-02 | Grade 2 (FDR) | 8106 |
| mweight | wspeed | sarcopenia | 6.8267e-03 | 1.0000e+00 | 5.0821e-02 | Grade 3 (Nominal) | 7700 |
| CMI | grip_weight | sarcopenia | 7.4867e-03 | 1.0000e+00 | 5.4971e-02 | Grade 3 (Nominal) | 11964 |
| WWI | rural2 | possible_sarcopenia | 9.1131e-03 | 1.0000e+00 | 6.6008e-02 | Grade 3 (Nominal) | 8078 |
| ABSI | smokev_r | possible_sarcopenia | 9.8721e-03 | 1.0000e+00 | 7.0552e-02 | Grade 3 (Nominal) | 8031 |
| wspeed | green_area | sarcopenia | 1.0133e-02 | 1.0000e+00 | 7.1467e-02 | Grade 3 (Nominal) | 7075 |
| CMI | CO | sarcopenia | 1.1690e-02 | 1.0000e+00 | 8.1373e-02 | Grade 3 (Nominal) | 11877 |
| mweight | extreme_rain | possible_sarcopenia | 1.6292e-02 | 1.0000e+00 | 1.1195e-01 | Grade 3 (Nominal) | 5573 |
| wspeed | temperature | sarcopenia | 1.7069e-02 | 1.0000e+00 | 1.1581e-01 | Grade 3 (Nominal) | 6847 |
| gripsum | drinkev_r | possible_sarcopenia | 1.8905e-02 | 1.0000e+00 | 1.2666e-01 | Grade 3 (Nominal) | 8319 |
| WWI | cesd10 | possible_sarcopenia | 1.9742e-02 | 1.0000e+00 | 1.3039e-01 | Grade 3 (Nominal) | 7808 |
| wspeed | precipitation | sarcopenia | 2.0015e-02 | 1.0000e+00 | 1.3039e-01 | Grade 3 (Nominal) | 6847 |
| mwaist | wspeed | possible_sarcopenia | 2.0190e-02 | 1.0000e+00 | 1.3039e-01 | Grade 3 (Nominal) | 7561 |
| bmi | diabe | possible_sarcopenia | 2.0723e-02 | 1.0000e+00 | 1.3194e-01 | Grade 3 (Nominal) | 7594 |
| ABSI | diabe | sarcopenia | 2.0924e-02 | 1.0000e+00 | 1.3194e-01 | Grade 3 (Nominal) | 12218 |
| mweight | drinkev_r | possible_sarcopenia | 2.3257e-02 | 1.0000e+00 | 1.4495e-01 | Grade 3 (Nominal) | 8258 |
| WWI | grip_weight | sarcopenia | 2.4900e-02 | 1.0000e+00 | 1.5171e-01 | Grade 3 (Nominal) | 14767 |
| wspeed | NO2 | possible_sarcopenia | 2.4907e-02 | 1.0000e+00 | 1.5171e-01 | Grade 3 (Nominal) | 7628 |
| CMI | relief | sarcopenia | 2.6862e-02 | 1.0000e+00 | 1.6018e-01 | Grade 3 (Nominal) | 10830 |
| gripsum | rural2 | sarcopenia | 2.6897e-02 | 1.0000e+00 | 1.6018e-01 | Grade 3 (Nominal) | 15117 |
| wspeed | O3 | sarcopenia | 2.8843e-02 | 1.0000e+00 | 1.6837e-01 | Grade 3 (Nominal) | 7589 |
| gripsum | green_area | sarcopenia | 2.9143e-02 | 1.0000e+00 | 1.6837e-01 | Grade 3 (Nominal) | 13750 |
| bmi | O3 | sarcopenia | 2.9214e-02 | 1.0000e+00 | 1.6837e-01 | Grade 3 (Nominal) | 14931 |
| mwaist | green_area | possible_sarcopenia | 3.1042e-02 | 1.0000e+00 | 1.7542e-01 | Grade 3 (Nominal) | 7438 |
| WWI | wspeed | possible_sarcopenia | 3.1092e-02 | 1.0000e+00 | 1.7542e-01 | Grade 3 (Nominal) | 7546 |
| WWI | extreme_rain | possible_sarcopenia | 3.2250e-02 | 1.0000e+00 | 1.8006e-01 | Grade 3 (Nominal) | 5535 |
| WHtR | wspeed | possible_sarcopenia | 3.3366e-02 | 1.0000e+00 | 1.8437e-01 | Grade 3 (Nominal) | 7523 |
| mwaist | drinkev_r | possible_sarcopenia | 3.6831e-02 | 1.0000e+00 | 2.0107e-01 | Grade 3 (Nominal) | 8089 |
| wspeed | green_area | possible_sarcopenia | 3.7459e-02 | 1.0000e+00 | 2.0107e-01 | Grade 3 (Nominal) | 7114 |
| ABSI | NDVI | possible_sarcopenia | 3.7513e-02 | 1.0000e+00 | 2.0107e-01 | Grade 3 (Nominal) | 7160 |
| mweight | green_area | possible_sarcopenia | 3.9812e-02 | 1.0000e+00 | 2.1128e-01 | Grade 3 (Nominal) | 7598 |
| gripsum | O3 | possible_sarcopenia | 4.0726e-02 | 1.0000e+00 | 2.1401e-01 | Grade 3 (Nominal) | 8205 |
| wspeed | relief | possible_sarcopenia | 4.1726e-02 | 1.0000e+00 | 2.1714e-01 | Grade 3 (Nominal) | 6873 |
| WHtR | cesd10 | sarcopenia | 4.3870e-02 | 1.0000e+00 | 2.2305e-01 | Grade 3 (Nominal) | 14573 |
| WWI | sex_age_group | sarcopenia | 4.4079e-02 | 1.0000e+00 | 2.2305e-01 | Grade 3 (Nominal) | 14908 |
| WWI | smokev_r | sarcopenia | 4.4110e-02 | 1.0000e+00 | 2.2305e-01 | Grade 3 (Nominal) | 14895 |
| mweight | sleeprl | possible_sarcopenia | 4.4553e-02 | 1.0000e+00 | 2.2318e-01 | Grade 3 (Nominal) | 7964 |
| ABSI | smokev_r | sarcopenia | 4.7578e-02 | 1.0000e+00 | 2.3613e-01 | Grade 3 (Nominal) | 14844 |
| gripsum | extreme_high | possible_sarcopenia | 4.9021e-02 | 1.0000e+00 | 2.4106e-01 | Grade 3 (Nominal) | 5608 |
| WHtR | gripsum | possible_sarcopenia | 5.0849e-02 | 1.0000e+00 | 2.4501e-01 | Non-significant | 8055 |
| WWI | diabe | sarcopenia | 5.2038e-02 | 1.0000e+00 | 2.4501e-01 | Non-significant | 12265 |
| mwaist | O3 | possible_sarcopenia | 5.2076e-02 | 1.0000e+00 | 2.4501e-01 | Non-significant | 7976 |
| ABSI | sex_age_group | sarcopenia | 5.2118e-02 | 1.0000e+00 | 2.4501e-01 | Non-significant | 14857 |
| CMI | CO | possible_sarcopenia | 5.2379e-02 | 1.0000e+00 | 2.4501e-01 | Non-significant | 6485 |
| ABSI | cesd10 | possible_sarcopenia | 5.2568e-02 | 1.0000e+00 | 2.4501e-01 | Non-significant | 7771 |
| WHtR | sleeprl | sarcopenia | 5.3798e-02 | 1.0000e+00 | 2.4858e-01 | Non-significant | 14521 |
| ABSI | rural2 | possible_sarcopenia | 5.5666e-02 | 1.0000e+00 | 2.5502e-01 | Non-significant | 8038 |
| wspeed | altitude | sarcopenia | 5.6219e-02 | 1.0000e+00 | 2.5537e-01 | Non-significant | 6847 |
| WWI | NDVI | possible_sarcopenia | 5.8232e-02 | 1.0000e+00 | 2.6229e-01 | Non-significant | 7196 |
| ABSI | climate_risk | sarcopenia | 6.1346e-02 | 1.0000e+00 | 2.7401e-01 | Non-significant | 10220 |
| wspeed | altitude | possible_sarcopenia | 6.4792e-02 | 1.0000e+00 | 2.8701e-01 | Non-significant | 6873 |
| wspeed | NDVI | sarcopenia | 6.5732e-02 | 1.0000e+00 | 2.8879e-01 | Non-significant | 6847 |
| wspeed | PM2.5 | possible_sarcopenia | 6.6321e-02 | 1.0000e+00 | 2.8901e-01 | Non-significant | 7628 |
| mweight | O3 | sarcopenia | 7.0823e-02 | 1.0000e+00 | 3.0545e-01 | Non-significant | 14946 |
| WWI | age_group | sarcopenia | 7.1378e-02 | 1.0000e+00 | 3.0545e-01 | Non-significant | 14908 |
| ABSI | extreme_drought | possible_sarcopenia | 7.1804e-02 | 1.0000e+00 | 3.0545e-01 | Non-significant | 5514 |
| bmi | SO2 | possible_sarcopenia | 7.4498e-02 | 1.0000e+00 | 3.1442e-01 | Non-significant | 8135 |
| wspeed | PM10 | possible_sarcopenia | 7.6476e-02 | 1.0000e+00 | 3.1914e-01 | Non-significant | 7628 |
| ABSI | extreme_high | sarcopenia | 7.7351e-02 | 1.0000e+00 | 3.1914e-01 | Non-significant | 10220 |
| ABSI | O3 | possible_sarcopenia | 7.7422e-02 | 1.0000e+00 | 3.1914e-01 | Non-significant | 7911 |
| gripsum | rural2 | possible_sarcopenia | 7.8000e-02 | 1.0000e+00 | 3.1914e-01 | Non-significant | 8338 |
| wspeed | precipitation | possible_sarcopenia | 8.0896e-02 | 1.0000e+00 | 3.2848e-01 | Non-significant | 6873 |
| gripsum | NO2 | sarcopenia | 8.1536e-02 | 1.0000e+00 | 3.2860e-01 | Non-significant | 14868 |
| ABSI | temperature | possible_sarcopenia | 8.2510e-02 | 1.0000e+00 | 3.3004e-01 | Non-significant | 7160 |
| mwaist | sex_age_group | sarcopenia | 8.5147e-02 | 1.0000e+00 | 3.3807e-01 | Non-significant | 14934 |
| mwaist | O3 | sarcopenia | 9.2489e-02 | 1.0000e+00 | 3.6451e-01 | Non-significant | 14682 |
| bmi | temperature | possible_sarcopenia | 9.4080e-02 | 1.0000e+00 | 3.6808e-01 | Non-significant | 7305 |
| WHtR | NDVI | possible_sarcopenia | 9.5722e-02 | 1.0000e+00 | 3.6971e-01 | Non-significant | 7176 |
| WHtR | sex_age_group | sarcopenia | 9.6460e-02 | 1.0000e+00 | 3.6971e-01 | Non-significant | 14872 |
| wspeed | temperature | possible_sarcopenia | 9.6567e-02 | 1.0000e+00 | 3.6971e-01 | Non-significant | 6873 |
| wspeed | gripsum | sarcopenia | 1.0059e-01 | 1.0000e+00 | 3.7968e-01 | Non-significant | 7576 |
| gripsum | wspeed | sarcopenia | 1.0059e-01 | 1.0000e+00 | 3.7968e-01 | Non-significant | 7576 |
| WWI | diabe | possible_sarcopenia | 1.0150e-01 | 1.0000e+00 | 3.8044e-01 | Non-significant | 7427 |
| ABSI | diabe | possible_sarcopenia | 1.0410e-01 | 1.0000e+00 | 3.8749e-01 | Non-significant | 7389 |
| WHtR | O3 | sarcopenia | 1.0497e-01 | 1.0000e+00 | 3.8804e-01 | Non-significant | 14623 |
| mweight | extreme_low | sarcopenia | 1.0886e-01 | 1.0000e+00 | 3.9963e-01 | Non-significant | 10315 |
| WHtR | relief | possible_sarcopenia | 1.1201e-01 | 1.0000e+00 | 4.0843e-01 | Non-significant | 7176 |
| bmi | sex_age_group | possible_sarcopenia | 1.1661e-01 | 1.0000e+00 | 4.2233e-01 | Non-significant | 8263 |
| bmi | CO | sarcopenia | 1.2183e-01 | 1.0000e+00 | 4.3827e-01 | Non-significant | 14931 |
| wspeed | extreme_drought | possible_sarcopenia | 1.2615e-01 | 1.0000e+00 | 4.4879e-01 | Non-significant | 5183 |
| bmi | extreme_high | possible_sarcopenia | 1.2643e-01 | 1.0000e+00 | 4.4879e-01 | Non-significant | 5563 |
| wspeed | extreme_high | sarcopenia | 1.3025e-01 | 1.0000e+00 | 4.5790e-01 | Non-significant | 5147 |
| bmi | NDVI | possible_sarcopenia | 1.3071e-01 | 1.0000e+00 | 4.5790e-01 | Non-significant | 7305 |
| WHtR | gripsum | sarcopenia | 1.3165e-01 | 1.0000e+00 | 4.5821e-01 | Non-significant | 14731 |
| WWI | extreme_high | sarcopenia | 1.3255e-01 | 1.0000e+00 | 4.5838e-01 | Non-significant | 10250 |
| CMI | extreme_high | sarcopenia | 1.3958e-01 | 1.0000e+00 | 4.7959e-01 | Non-significant | 8312 |
| ABSI | wspeed | possible_sarcopenia | 1.4154e-01 | 1.0000e+00 | 4.8323e-01 | Non-significant | 7509 |
| WWI | extreme_drought | sarcopenia | 1.4787e-01 | 1.0000e+00 | 4.9935e-01 | Non-significant | 10250 |
| WWI | extreme_drought | possible_sarcopenia | 1.4838e-01 | 1.0000e+00 | 4.9935e-01 | Non-significant | 5535 |
| gripsum | altitude | possible_sarcopenia | 1.4958e-01 | 1.0000e+00 | 4.9935e-01 | Non-significant | 7372 |
| wspeed | age_group | sarcopenia | 1.5111e-01 | 1.0000e+00 | 4.9935e-01 | Non-significant | 7719 |
| ABSI | extreme_drought | sarcopenia | 1.5168e-01 | 1.0000e+00 | 4.9935e-01 | Non-significant | 10220 |
| mweight | wspeed | possible_sarcopenia | 1.5186e-01 | 1.0000e+00 | 4.9935e-01 | Non-significant | 7731 |
| WHtR | extreme_drought | possible_sarcopenia | 1.5654e-01 | 1.0000e+00 | 5.1043e-01 | Non-significant | 5521 |
| mwaist | NDVI | possible_sarcopenia | 1.5790e-01 | 1.0000e+00 | 5.1043e-01 | Non-significant | 7222 |
| WHtR | O3 | possible_sarcopenia | 1.5808e-01 | 1.0000e+00 | 5.1043e-01 | Non-significant | 7929 |
| WWI | climate_risk | sarcopenia | 1.5955e-01 | 1.0000e+00 | 5.1207e-01 | Non-significant | 10250 |
| mweight | sleeprl | sarcopenia | 1.6194e-01 | 1.0000e+00 | 5.1207e-01 | Non-significant | 14840 |
| WHtR | CO | possible_sarcopenia | 1.6215e-01 | 1.0000e+00 | 5.1207e-01 | Non-significant | 7929 |
| mweight | age_group | sarcopenia | 1.6241e-01 | 1.0000e+00 | 5.1207e-01 | Non-significant | 15197 |
| mweight | temperature | sarcopenia | 1.6590e-01 | 1.0000e+00 | 5.1790e-01 | Non-significant | 13403 |
| mwaist | diabe | sarcopenia | 1.6619e-01 | 1.0000e+00 | 5.1790e-01 | Non-significant | 12285 |
| wspeed | AQI | sarcopenia | 1.6846e-01 | 1.0000e+00 | 5.2046e-01 | Non-significant | 7237 |
| CMI | relief | possible_sarcopenia | 1.7161e-01 | 1.0000e+00 | 5.2046e-01 | Non-significant | 5951 |
| WHtR | altitude | possible_sarcopenia | 1.7193e-01 | 1.0000e+00 | 5.2046e-01 | Non-significant | 7176 |
| CMI | extreme_low | sarcopenia | 1.7234e-01 | 1.0000e+00 | 5.2046e-01 | Non-significant | 8312 |
| CMI | drinkev_r | possible_sarcopenia | 1.7279e-01 | 1.0000e+00 | 5.2046e-01 | Non-significant | 6585 |
| WHtR | diabe | sarcopenia | 1.7393e-01 | 1.0000e+00 | 5.2046e-01 | Non-significant | 12228 |
| CMI | smokev_r | possible_sarcopenia | 1.7477e-01 | 1.0000e+00 | 5.2046e-01 | Non-significant | 6594 |
| WHtR | diabe | possible_sarcopenia | 1.7478e-01 | 1.0000e+00 | 5.2046e-01 | Non-significant | 7404 |
| WHtR | extreme_rain | possible_sarcopenia | 1.7935e-01 | 1.0000e+00 | 5.3110e-01 | Non-significant | 5521 |
| ABSI | extreme_rain | possible_sarcopenia | 1.8351e-01 | 1.0000e+00 | 5.4044e-01 | Non-significant | 5514 |
| CMI | altitude | sarcopenia | 1.8515e-01 | 1.0000e+00 | 5.4162e-01 | Non-significant | 10830 |
| WHtR | green_area | possible_sarcopenia | 1.8593e-01 | 1.0000e+00 | 5.4162e-01 | Non-significant | 7396 |
| mweight | CO | sarcopenia | 1.8854e-01 | 1.0000e+00 | 5.4258e-01 | Non-significant | 14946 |
| CMI | AQI | possible_sarcopenia | 1.8927e-01 | 1.0000e+00 | 5.4258e-01 | Non-significant | 6227 |
| wspeed | cesd10 | possible_sarcopenia | 1.8930e-01 | 1.0000e+00 | 5.4258e-01 | Non-significant | 7509 |
| CMI | wspeed | possible_sarcopenia | 1.9073e-01 | 1.0000e+00 | 5.4378e-01 | Non-significant | 6209 |
| CMI | climate_risk | sarcopenia | 1.9212e-01 | 1.0000e+00 | 5.4485e-01 | Non-significant | 8312 |
| CMI | extreme_drought | possible_sarcopenia | 1.9339e-01 | 1.0000e+00 | 5.4558e-01 | Non-significant | 4522 |
| mwaist | CO | sarcopenia | 2.0377e-01 | 1.0000e+00 | 5.6933e-01 | Non-significant | 14682 |
| ABSI | rural2 | sarcopenia | 2.0394e-01 | 1.0000e+00 | 5.6933e-01 | Non-significant | 14857 |
| WHtR | extreme_drought | sarcopenia | 2.0534e-01 | 1.0000e+00 | 5.7028e-01 | Non-significant | 10227 |
| wspeed | smokev_r | sarcopenia | 2.0764e-01 | 1.0000e+00 | 5.7369e-01 | Non-significant | 7712 |
| WWI | rural2 | sarcopenia | 2.1056e-01 | 1.0000e+00 | 5.7876e-01 | Non-significant | 14908 |
| mwaist | CO | possible_sarcopenia | 2.1356e-01 | 1.0000e+00 | 5.8288e-01 | Non-significant | 7976 |
| bmi | green_area | sarcopenia | 2.1621e-01 | 1.0000e+00 | 5.8288e-01 | Non-significant | 13805 |
| mweight | sex_age_group | sarcopenia | 2.1644e-01 | 1.0000e+00 | 5.8288e-01 | Non-significant | 15197 |
| CMI | NDVI | possible_sarcopenia | 2.1838e-01 | 1.0000e+00 | 5.8288e-01 | Non-significant | 5951 |
| mweight | rural2 | sarcopenia | 2.1856e-01 | 1.0000e+00 | 5.8288e-01 | Non-significant | 15197 |
| mweight | AQI | possible_sarcopenia | 2.1858e-01 | 1.0000e+00 | 5.8288e-01 | Non-significant | 7767 |
| mweight | CO | possible_sarcopenia | 2.2320e-01 | 1.0000e+00 | 5.9225e-01 | Non-significant | 8146 |
| ABSI | age_group | possible_sarcopenia | 2.2761e-01 | 1.0000e+00 | 6.0098e-01 | Non-significant | 8038 |
| CMI | gripsum | possible_sarcopenia | 2.3196e-01 | 1.0000e+00 | 6.0845e-01 | Non-significant | 6596 |
| mweight | extreme_high | sarcopenia | 2.3299e-01 | 1.0000e+00 | 6.0845e-01 | Non-significant | 10315 |
| bmi | sex_age_group | sarcopenia | 2.3385e-01 | 1.0000e+00 | 6.0845e-01 | Non-significant | 15179 |
| mwaist | diabe | possible_sarcopenia | 2.3500e-01 | 1.0000e+00 | 6.0851e-01 | Non-significant | 7452 |
| mwaist | temperature | sarcopenia | 2.3780e-01 | 1.0000e+00 | 6.1279e-01 | Non-significant | 13247 |
| mwaist | extreme_drought | possible_sarcopenia | 2.3999e-01 | 1.0000e+00 | 6.1547e-01 | Non-significant | 5550 |
| gripsum | temperature | sarcopenia | 2.4803e-01 | 1.0000e+00 | 6.3088e-01 | Non-significant | 13334 |
| mwaist | sleeprl | sarcopenia | 2.4835e-01 | 1.0000e+00 | 6.3088e-01 | Non-significant | 14580 |
| CMI | rural2 | possible_sarcopenia | 2.4953e-01 | 1.0000e+00 | 6.3090e-01 | Non-significant | 6598 |
| bmi | wspeed | possible_sarcopenia | 2.5197e-01 | 1.0000e+00 | 6.3406e-01 | Non-significant | 7717 |
| WHtR | relief | sarcopenia | 2.5479e-01 | 1.0000e+00 | 6.3816e-01 | Non-significant | 13193 |
| wspeed | drinkev_r | possible_sarcopenia | 2.5882e-01 | 1.0000e+00 | 6.3902e-01 | Non-significant | 7737 |
| WHtR | CO | sarcopenia | 2.5902e-01 | 1.0000e+00 | 6.3902e-01 | Non-significant | 14623 |
| wspeed | climate_risk | possible_sarcopenia | 2.5952e-01 | 1.0000e+00 | 6.3902e-01 | Non-significant | 5183 |
| gripsum | AQI | sarcopenia | 2.5990e-01 | 1.0000e+00 | 6.3902e-01 | Non-significant | 14173 |
| gripsum | AQI | possible_sarcopenia | 2.6147e-01 | 1.0000e+00 | 6.3995e-01 | Non-significant | 7824 |
| mwaist | sleeprl | possible_sarcopenia | 2.6405e-01 | 1.0000e+00 | 6.4332e-01 | Non-significant | 7796 |
| ABSI | sex_age_group | possible_sarcopenia | 2.6728e-01 | 1.0000e+00 | 6.4823e-01 | Non-significant | 8038 |
| ABSI | precipitation | possible_sarcopenia | 2.6976e-01 | 1.0000e+00 | 6.5061e-01 | Non-significant | 7160 |
| CMI | smokev_r | sarcopenia | 2.7068e-01 | 1.0000e+00 | 6.5061e-01 | Non-significant | 12079 |
| bmi | cesd10 | sarcopenia | 2.7567e-01 | 1.0000e+00 | 6.5912e-01 | Non-significant | 14875 |
| mwaist | relief | possible_sarcopenia | 2.7769e-01 | 1.0000e+00 | 6.5912e-01 | Non-significant | 7222 |
| WWI | sleeprl | possible_sarcopenia | 2.7791e-01 | 1.0000e+00 | 6.5912e-01 | Non-significant | 7772 |
| WWI | precipitation | possible_sarcopenia | 2.8047e-01 | 1.0000e+00 | 6.6224e-01 | Non-significant | 7196 |
| WHtR | precipitation | possible_sarcopenia | 2.8179e-01 | 1.0000e+00 | 6.6246e-01 | Non-significant | 7176 |
| CMI | AQI | sarcopenia | 2.8581e-01 | 1.0000e+00 | 6.6898e-01 | Non-significant | 11410 |
| CMI | O3 | possible_sarcopenia | 2.8975e-01 | 1.0000e+00 | 6.7238e-01 | Non-significant | 6485 |
| WWI | temperature | possible_sarcopenia | 2.8977e-01 | 1.0000e+00 | 6.7238e-01 | Non-significant | 7196 |
| WWI | age_group | possible_sarcopenia | 2.9329e-01 | 1.0000e+00 | 6.7721e-01 | Non-significant | 8078 |
| wspeed | diabe | possible_sarcopenia | 2.9439e-01 | 1.0000e+00 | 6.7721e-01 | Non-significant | 7182 |
| WWI | SO2 | possible_sarcopenia | 2.9598e-01 | 1.0000e+00 | 6.7721e-01 | Non-significant | 7947 |
| ABSI | altitude | possible_sarcopenia | 2.9691e-01 | 1.0000e+00 | 6.7721e-01 | Non-significant | 7160 |
| mweight | altitude | sarcopenia | 3.0029e-01 | 1.0000e+00 | 6.7760e-01 | Non-significant | 13403 |
| CMI | age_group | sarcopenia | 3.0213e-01 | 1.0000e+00 | 6.7760e-01 | Non-significant | 12088 |
| gripsum | smokev_r | possible_sarcopenia | 3.0214e-01 | 1.0000e+00 | 6.7760e-01 | Non-significant | 8331 |
| WHtR | age_group | sarcopenia | 3.0214e-01 | 1.0000e+00 | 6.7760e-01 | Non-significant | 14872 |
| bmi | precipitation | sarcopenia | 3.0680e-01 | 1.0000e+00 | 6.8193e-01 | Non-significant | 13387 |
| CMI | NDVI | sarcopenia | 3.0703e-01 | 1.0000e+00 | 6.8193e-01 | Non-significant | 10830 |
| bmi | extreme_high | sarcopenia | 3.0789e-01 | 1.0000e+00 | 6.8193e-01 | Non-significant | 10304 |
| bmi | sleeprl | possible_sarcopenia | 3.1077e-01 | 1.0000e+00 | 6.8549e-01 | Non-significant | 7954 |
| CMI | rural2 | sarcopenia | 3.1446e-01 | 1.0000e+00 | 6.8927e-01 | Non-significant | 12088 |
| ABSI | grip_weight | sarcopenia | 3.1514e-01 | 1.0000e+00 | 6.8927e-01 | Non-significant | 14716 |
| mweight | extreme_high | possible_sarcopenia | 3.1634e-01 | 1.0000e+00 | 6.8927e-01 | Non-significant | 5573 |
| gripsum | extreme_rain | possible_sarcopenia | 3.2086e-01 | 1.0000e+00 | 6.8972e-01 | Non-significant | 5608 |
| WHtR | cesd10 | possible_sarcopenia | 3.2150e-01 | 1.0000e+00 | 6.8972e-01 | Non-significant | 7789 |
| wspeed | grip_weight | sarcopenia | 3.2277e-01 | 1.0000e+00 | 6.8972e-01 | Non-significant | 7557 |
| ABSI | gripsum | sarcopenia | 3.2380e-01 | 1.0000e+00 | 6.8972e-01 | Non-significant | 14716 |
| mwaist | climate_risk | sarcopenia | 3.2418e-01 | 1.0000e+00 | 6.8972e-01 | Non-significant | 10264 |
| WWI | relief | sarcopenia | 3.2478e-01 | 1.0000e+00 | 6.8972e-01 | Non-significant | 13225 |
| WHtR | sex_age_group | possible_sarcopenia | 3.2556e-01 | 1.0000e+00 | 6.8972e-01 | Non-significant | 8057 |
| CMI | extreme_rain | possible_sarcopenia | 3.3028e-01 | 1.0000e+00 | 6.9126e-01 | Non-significant | 4522 |
| gripsum | temperature | possible_sarcopenia | 3.3112e-01 | 1.0000e+00 | 6.9126e-01 | Non-significant | 7372 |
| mweight | SO2 | possible_sarcopenia | 3.3206e-01 | 1.0000e+00 | 6.9126e-01 | Non-significant | 8146 |
| CMI | gripsum | sarcopenia | 3.3392e-01 | 1.0000e+00 | 6.9126e-01 | Non-significant | 11972 |
| ABSI | PM2.5 | possible_sarcopenia | 3.3435e-01 | 1.0000e+00 | 6.9126e-01 | Non-significant | 7911 |
| wspeed | grip_bmi | sarcopenia | 3.3500e-01 | 1.0000e+00 | 6.9126e-01 | Non-significant | 7522 |
| gripsum | SO2 | possible_sarcopenia | 3.3596e-01 | 1.0000e+00 | 6.9126e-01 | Non-significant | 8205 |
| gripsum | NDVI | possible_sarcopenia | 3.3660e-01 | 1.0000e+00 | 6.9126e-01 | Non-significant | 7372 |
| wspeed | SO2 | sarcopenia | 3.4168e-01 | 1.0000e+00 | 6.9900e-01 | Non-significant | 7589 |
| bmi | CO | possible_sarcopenia | 3.4371e-01 | 1.0000e+00 | 7.0049e-01 | Non-significant | 8135 |
| ABSI | age_group | sarcopenia | 3.4602e-01 | 1.0000e+00 | 7.0253e-01 | Non-significant | 14857 |
| WHtR | climate_risk | sarcopenia | 3.4779e-01 | 1.0000e+00 | 7.0346e-01 | Non-significant | 10227 |
| ABSI | green_area | possible_sarcopenia | 3.5128e-01 | 1.0000e+00 | 7.0534e-01 | Non-significant | 7379 |
| mweight | smokev_r | sarcopenia | 3.5136e-01 | 1.0000e+00 | 7.0534e-01 | Non-significant | 15184 |
| wspeed | CO | possible_sarcopenia | 3.5361e-01 | 1.0000e+00 | 7.0618e-01 | Non-significant | 7628 |
| WWI | O3 | possible_sarcopenia | 3.5441e-01 | 1.0000e+00 | 7.0618e-01 | Non-significant | 7947 |
| mweight | climate_risk | possible_sarcopenia | 3.5640e-01 | 1.0000e+00 | 7.0752e-01 | Non-significant | 5573 |
| ABSI | SO2 | possible_sarcopenia | 3.6399e-01 | 1.0000e+00 | 7.1992e-01 | Non-significant | 7911 |
| ABSI | relief | sarcopenia | 3.7830e-01 | 1.0000e+00 | 7.4548e-01 | Non-significant | 13180 |
| gripsum | diabe | sarcopenia | 3.8502e-01 | 1.0000e+00 | 7.5594e-01 | Non-significant | 12430 |
| mwaist | precipitation | sarcopenia | 3.9554e-01 | 1.0000e+00 | 7.7376e-01 | Non-significant | 13247 |
| gripsum | NDVI | sarcopenia | 3.9960e-01 | 1.0000e+00 | 7.7768e-01 | Non-significant | 13334 |
| ABSI | AQI | possible_sarcopenia | 4.0054e-01 | 1.0000e+00 | 7.7768e-01 | Non-significant | 7539 |
| CMI | cesd10 | sarcopenia | 4.0302e-01 | 1.0000e+00 | 7.7768e-01 | Non-significant | 11872 |
| WWI | CO | possible_sarcopenia | 4.0335e-01 | 1.0000e+00 | 7.7768e-01 | Non-significant | 7947 |
| WHtR | SO2 | possible_sarcopenia | 4.0505e-01 | 1.0000e+00 | 7.7817e-01 | Non-significant | 7929 |
| WHtR | rural2 | possible_sarcopenia | 4.0759e-01 | 1.0000e+00 | 7.8025e-01 | Non-significant | 8057 |
| mweight | age_group | possible_sarcopenia | 4.1319e-01 | 1.0000e+00 | 7.8815e-01 | Non-significant | 8277 |
| WHtR | precipitation | sarcopenia | 4.2176e-01 | 1.0000e+00 | 7.9590e-01 | Non-significant | 13193 |
| WHtR | PM10 | possible_sarcopenia | 4.2254e-01 | 1.0000e+00 | 7.9590e-01 | Non-significant | 7929 |
| mwaist | extreme_drought | sarcopenia | 4.2255e-01 | 1.0000e+00 | 7.9590e-01 | Non-significant | 10264 |
| mweight | green_area | sarcopenia | 4.2352e-01 | 1.0000e+00 | 7.9590e-01 | Non-significant | 13817 |
| mwaist | altitude | possible_sarcopenia | 4.2468e-01 | 1.0000e+00 | 7.9590e-01 | Non-significant | 7222 |
| ABSI | extreme_high | possible_sarcopenia | 4.2878e-01 | 1.0000e+00 | 7.9676e-01 | Non-significant | 5514 |
| CMI | sex_age_group | possible_sarcopenia | 4.2953e-01 | 1.0000e+00 | 7.9676e-01 | Non-significant | 6598 |
| WHtR | smokev_r | possible_sarcopenia | 4.2985e-01 | 1.0000e+00 | 7.9676e-01 | Non-significant | 8050 |
| WHtR | AQI | possible_sarcopenia | 4.3188e-01 | 1.0000e+00 | 7.9676e-01 | Non-significant | 7556 |
| mweight | extreme_drought | sarcopenia | 4.3257e-01 | 1.0000e+00 | 7.9676e-01 | Non-significant | 10315 |
| bmi | altitude | possible_sarcopenia | 4.3762e-01 | 1.0000e+00 | 8.0331e-01 | Non-significant | 7305 |
| bmi | climate_risk | possible_sarcopenia | 4.3981e-01 | 1.0000e+00 | 8.0448e-01 | Non-significant | 5563 |
| gripsum | sleeprl | possible_sarcopenia | 4.4263e-01 | 1.0000e+00 | 8.0448e-01 | Non-significant | 8011 |
| mweight | PM2.5 | possible_sarcopenia | 4.4359e-01 | 1.0000e+00 | 8.0448e-01 | Non-significant | 8146 |
| WWI | extreme_low | sarcopenia | 4.4426e-01 | 1.0000e+00 | 8.0448e-01 | Non-significant | 10250 |
| mwaist | NDVI | sarcopenia | 4.4710e-01 | 1.0000e+00 | 8.0681e-01 | Non-significant | 13247 |
| CMI | PM2.5 | sarcopenia | 4.4896e-01 | 1.0000e+00 | 8.0681e-01 | Non-significant | 11877 |
| mweight | NO2 | possible_sarcopenia | 4.5007e-01 | 1.0000e+00 | 8.0681e-01 | Non-significant | 8146 |
| mweight | NO2 | sarcopenia | 4.5387e-01 | 1.0000e+00 | 8.1092e-01 | Non-significant | 14946 |
| gripsum | climate_risk | possible_sarcopenia | 4.6090e-01 | 1.0000e+00 | 8.2030e-01 | Non-significant | 5608 |
| gripsum | extreme_drought | possible_sarcopenia | 4.6218e-01 | 1.0000e+00 | 8.2030e-01 | Non-significant | 5608 |
| WWI | PM2.5 | possible_sarcopenia | 4.6942e-01 | 1.0000e+00 | 8.2374e-01 | Non-significant | 7947 |
| WHtR | smokev_r | sarcopenia | 4.6945e-01 | 1.0000e+00 | 8.2374e-01 | Non-significant | 14859 |
| bmi | temperature | sarcopenia | 4.7126e-01 | 1.0000e+00 | 8.2374e-01 | Non-significant | 13387 |
| WHtR | temperature | sarcopenia | 4.7138e-01 | 1.0000e+00 | 8.2374e-01 | Non-significant | 13193 |
| CMI | green_area | possible_sarcopenia | 4.7180e-01 | 1.0000e+00 | 8.2374e-01 | Non-significant | 6098 |
| wspeed | extreme_low | sarcopenia | 4.7474e-01 | 1.0000e+00 | 8.2617e-01 | Non-significant | 5147 |
| mwaist | AQI | possible_sarcopenia | 4.7948e-01 | 1.0000e+00 | 8.2951e-01 | Non-significant | 7600 |
| CMI | PM10 | sarcopenia | 4.7975e-01 | 1.0000e+00 | 8.2951e-01 | Non-significant | 11877 |
| WHtR | drinkev_r | possible_sarcopenia | 4.8421e-01 | 1.0000e+00 | 8.3452e-01 | Non-significant | 8038 |
| mwaist | relief | sarcopenia | 4.8647e-01 | 1.0000e+00 | 8.3573e-01 | Non-significant | 13247 |
| bmi | smokev_r | sarcopenia | 4.8947e-01 | 1.0000e+00 | 8.3782e-01 | Non-significant | 15166 |
| ABSI | extreme_low | possible_sarcopenia | 4.9081e-01 | 1.0000e+00 | 8.3782e-01 | Non-significant | 5514 |
| mwaist | precipitation | possible_sarcopenia | 4.9424e-01 | 1.0000e+00 | 8.4100e-01 | Non-significant | 7222 |
| ABSI | NDVI | sarcopenia | 4.9886e-01 | 1.0000e+00 | 8.4419e-01 | Non-significant | 13180 |
| mweight | precipitation | sarcopenia | 4.9927e-01 | 1.0000e+00 | 8.4419e-01 | Non-significant | 13403 |
| CMI | NO2 | sarcopenia | 5.0215e-01 | 1.0000e+00 | 8.4639e-01 | Non-significant | 11877 |
| CMI | diabe | sarcopenia | 5.0575e-01 | 1.0000e+00 | 8.4672e-01 | Non-significant | 10042 |
| WWI | gripsum | sarcopenia | 5.0636e-01 | 1.0000e+00 | 8.4672e-01 | Non-significant | 14767 |
| bmi | extreme_low | sarcopenia | 5.0828e-01 | 1.0000e+00 | 8.4672e-01 | Non-significant | 10304 |
| bmi | NO2 | sarcopenia | 5.0978e-01 | 1.0000e+00 | 8.4672e-01 | Non-significant | 14931 |
| ABSI | extreme_rain | sarcopenia | 5.1024e-01 | 1.0000e+00 | 8.4672e-01 | Non-significant | 10220 |
| wspeed | NO2 | sarcopenia | 5.1264e-01 | 1.0000e+00 | 8.4807e-01 | Non-significant | 7589 |
| mweight | sex_age_group | possible_sarcopenia | 5.1987e-01 | 1.0000e+00 | 8.5120e-01 | Non-significant | 8277 |
| WHtR | AQI | sarcopenia | 5.2000e-01 | 1.0000e+00 | 8.5120e-01 | Non-significant | 13929 |
| WHtR | SO2 | sarcopenia | 5.2085e-01 | 1.0000e+00 | 8.5120e-01 | Non-significant | 14623 |
| CMI | O3 | sarcopenia | 5.2187e-01 | 1.0000e+00 | 8.5120e-01 | Non-significant | 11877 |
| WHtR | NDVI | sarcopenia | 5.2247e-01 | 1.0000e+00 | 8.5120e-01 | Non-significant | 13193 |
| mwaist | sex_age_group | possible_sarcopenia | 5.2519e-01 | 1.0000e+00 | 8.5304e-01 | Non-significant | 8108 |
| bmi | rural2 | sarcopenia | 5.3000e-01 | 1.0000e+00 | 8.5664e-01 | Non-significant | 15179 |
| mwaist | extreme_rain | sarcopenia | 5.3249e-01 | 1.0000e+00 | 8.5664e-01 | Non-significant | 10264 |
| ABSI | PM2.5 | sarcopenia | 5.3350e-01 | 1.0000e+00 | 8.5664e-01 | Non-significant | 14609 |
| gripsum | relief | sarcopenia | 5.3649e-01 | 1.0000e+00 | 8.5664e-01 | Non-significant | 13334 |
| mweight | NDVI | sarcopenia | 5.3663e-01 | 1.0000e+00 | 8.5664e-01 | Non-significant | 13403 |
| WHtR | PM10 | sarcopenia | 5.3700e-01 | 1.0000e+00 | 8.5664e-01 | Non-significant | 14623 |
| WHtR | extreme_high | possible_sarcopenia | 5.4235e-01 | 1.0000e+00 | 8.6261e-01 | Non-significant | 5521 |
| CMI | temperature | possible_sarcopenia | 5.4715e-01 | 1.0000e+00 | 8.6767e-01 | Non-significant | 5951 |
| mwaist | gripsum | sarcopenia | 5.5257e-01 | 1.0000e+00 | 8.6800e-01 | Non-significant | 14793 |
| CMI | precipitation | possible_sarcopenia | 5.5291e-01 | 1.0000e+00 | 8.6800e-01 | Non-significant | 5951 |
| gripsum | extreme_drought | sarcopenia | 5.5354e-01 | 1.0000e+00 | 8.6800e-01 | Non-significant | 10274 |
| WWI | AQI | sarcopenia | 5.5383e-01 | 1.0000e+00 | 8.6800e-01 | Non-significant | 13961 |
| CMI | sleeprl | sarcopenia | 5.5745e-01 | 1.0000e+00 | 8.6923e-01 | Non-significant | 11828 |
| mwaist | PM10 | sarcopenia | 5.5786e-01 | 1.0000e+00 | 8.6923e-01 | Non-significant | 14682 |
| WWI | altitude | sarcopenia | 5.6102e-01 | 1.0000e+00 | 8.7156e-01 | Non-significant | 13225 |
| WHtR | altitude | sarcopenia | 5.6300e-01 | 1.0000e+00 | 8.7156e-01 | Non-significant | 13193 |
| wspeed | climate_risk | sarcopenia | 5.6424e-01 | 1.0000e+00 | 8.7156e-01 | Non-significant | 5147 |
| bmi | altitude | sarcopenia | 5.7023e-01 | 1.0000e+00 | 8.7615e-01 | Non-significant | 13387 |
| gripsum | precipitation | sarcopenia | 5.7181e-01 | 1.0000e+00 | 8.7615e-01 | Non-significant | 13334 |
| ABSI | sleeprl | possible_sarcopenia | 5.7279e-01 | 1.0000e+00 | 8.7615e-01 | Non-significant | 7736 |
| ABSI | temperature | sarcopenia | 5.7375e-01 | 1.0000e+00 | 8.7615e-01 | Non-significant | 13180 |
| ABSI | CO | possible_sarcopenia | 5.7627e-01 | 1.0000e+00 | 8.7750e-01 | Non-significant | 7911 |
| mwaist | smokev_r | possible_sarcopenia | 5.7813e-01 | 1.0000e+00 | 8.7784e-01 | Non-significant | 8101 |
| mwaist | age_group | possible_sarcopenia | 5.8850e-01 | 1.0000e+00 | 8.8468e-01 | Non-significant | 8108 |
| WWI | climate_risk | possible_sarcopenia | 5.9136e-01 | 1.0000e+00 | 8.8468e-01 | Non-significant | 5535 |
| mweight | extreme_drought | possible_sarcopenia | 5.9273e-01 | 1.0000e+00 | 8.8468e-01 | Non-significant | 5573 |
| mwaist | PM10 | possible_sarcopenia | 5.9289e-01 | 1.0000e+00 | 8.8468e-01 | Non-significant | 7976 |
| mwaist | extreme_high | possible_sarcopenia | 5.9332e-01 | 1.0000e+00 | 8.8468e-01 | Non-significant | 5550 |
| wspeed | extreme_low | possible_sarcopenia | 5.9690e-01 | 1.0000e+00 | 8.8468e-01 | Non-significant | 5183 |
| WWI | relief | possible_sarcopenia | 5.9885e-01 | 1.0000e+00 | 8.8468e-01 | Non-significant | 7196 |
| mweight | precipitation | possible_sarcopenia | 6.0012e-01 | 1.0000e+00 | 8.8468e-01 | Non-significant | 7317 |
| WWI | NO2 | possible_sarcopenia | 6.0024e-01 | 1.0000e+00 | 8.8468e-01 | Non-significant | 7947 |
| ABSI | O3 | sarcopenia | 6.0080e-01 | 1.0000e+00 | 8.8468e-01 | Non-significant | 14609 |
| WWI | NDVI | sarcopenia | 6.0291e-01 | 1.0000e+00 | 8.8468e-01 | Non-significant | 13225 |
| WWI | green_area | possible_sarcopenia | 6.0381e-01 | 1.0000e+00 | 8.8468e-01 | Non-significant | 7411 |
| mwaist | extreme_low | sarcopenia | 6.0559e-01 | 1.0000e+00 | 8.8468e-01 | Non-significant | 10264 |
| gripsum | altitude | sarcopenia | 6.1007e-01 | 1.0000e+00 | 8.8468e-01 | Non-significant | 13334 |
| WHtR | climate_risk | possible_sarcopenia | 6.1097e-01 | 1.0000e+00 | 8.8468e-01 | Non-significant | 5521 |
| ABSI | climate_risk | possible_sarcopenia | 6.1139e-01 | 1.0000e+00 | 8.8468e-01 | Non-significant | 5514 |
| ABSI | green_area | sarcopenia | 6.1284e-01 | 1.0000e+00 | 8.8468e-01 | Non-significant | 13505 |
| mweight | temperature | possible_sarcopenia | 6.1344e-01 | 1.0000e+00 | 8.8468e-01 | Non-significant | 7317 |
| mweight | PM10 | possible_sarcopenia | 6.1399e-01 | 1.0000e+00 | 8.8468e-01 | Non-significant | 8146 |
| gripsum | PM2.5 | sarcopenia | 6.1826e-01 | 1.0000e+00 | 8.8550e-01 | Non-significant | 14868 |
| mweight | relief | sarcopenia | 6.2372e-01 | 1.0000e+00 | 8.8550e-01 | Non-significant | 13403 |
| ABSI | altitude | sarcopenia | 6.2423e-01 | 1.0000e+00 | 8.8550e-01 | Non-significant | 13180 |
| WHtR | age_group | possible_sarcopenia | 6.2739e-01 | 1.0000e+00 | 8.8550e-01 | Non-significant | 8057 |
| mwaist | SO2 | sarcopenia | 6.2792e-01 | 1.0000e+00 | 8.8550e-01 | Non-significant | 14682 |
| ABSI | extreme_low | sarcopenia | 6.2960e-01 | 1.0000e+00 | 8.8550e-01 | Non-significant | 10220 |
| WHtR | PM2.5 | possible_sarcopenia | 6.3012e-01 | 1.0000e+00 | 8.8550e-01 | Non-significant | 7929 |
| mweight | extreme_rain | sarcopenia | 6.3291e-01 | 1.0000e+00 | 8.8550e-01 | Non-significant | 10315 |
| mwaist | rural2 | possible_sarcopenia | 6.3369e-01 | 1.0000e+00 | 8.8550e-01 | Non-significant | 8108 |
| bmi | gripsum | possible_sarcopenia | 6.3376e-01 | 1.0000e+00 | 8.8550e-01 | Non-significant | 8261 |
| CMI | extreme_low | possible_sarcopenia | 6.3643e-01 | 1.0000e+00 | 8.8550e-01 | Non-significant | 4522 |
| mwaist | temperature | possible_sarcopenia | 6.3743e-01 | 1.0000e+00 | 8.8550e-01 | Non-significant | 7222 |
| WHtR | temperature | possible_sarcopenia | 6.4168e-01 | 1.0000e+00 | 8.8550e-01 | Non-significant | 7176 |
| ABSI | AQI | sarcopenia | 6.4680e-01 | 1.0000e+00 | 8.8550e-01 | Non-significant | 13915 |
| CMI | PM2.5 | possible_sarcopenia | 6.4722e-01 | 1.0000e+00 | 8.8550e-01 | Non-significant | 6485 |
| gripsum | diabe | possible_sarcopenia | 6.4781e-01 | 1.0000e+00 | 8.8550e-01 | Non-significant | 7664 |
| bmi | cesd10 | possible_sarcopenia | 6.4864e-01 | 1.0000e+00 | 8.8550e-01 | Non-significant | 7989 |
| WWI | drinkev_r | sarcopenia | 6.4887e-01 | 1.0000e+00 | 8.8550e-01 | Non-significant | 14876 |
| gripsum | green_area | possible_sarcopenia | 6.4903e-01 | 1.0000e+00 | 8.8550e-01 | Non-significant | 7655 |
| ABSI | PM10 | sarcopenia | 6.4926e-01 | 1.0000e+00 | 8.8550e-01 | Non-significant | 14609 |
| ABSI | gripsum | possible_sarcopenia | 6.4926e-01 | 1.0000e+00 | 8.8550e-01 | Non-significant | 8036 |
| gripsum | extreme_rain | sarcopenia | 6.5227e-01 | 1.0000e+00 | 8.8698e-01 | Non-significant | 10274 |
| WWI | AQI | possible_sarcopenia | 6.5365e-01 | 1.0000e+00 | 8.8698e-01 | Non-significant | 7573 |
| CMI | temperature | sarcopenia | 6.5794e-01 | 1.0000e+00 | 8.9055e-01 | Non-significant | 10830 |
| bmi | precipitation | possible_sarcopenia | 6.6895e-01 | 1.0000e+00 | 9.0188e-01 | Non-significant | 7305 |
| WWI | drinkev_r | possible_sarcopenia | 6.7144e-01 | 1.0000e+00 | 9.0188e-01 | Non-significant | 8059 |
| gripsum | extreme_low | possible_sarcopenia | 6.7155e-01 | 1.0000e+00 | 9.0188e-01 | Non-significant | 5608 |
| mwaist | cesd10 | sarcopenia | 6.7305e-01 | 1.0000e+00 | 9.0188e-01 | Non-significant | 14633 |
| WWI | extreme_low | possible_sarcopenia | 6.7627e-01 | 1.0000e+00 | 9.0210e-01 | Non-significant | 5535 |
| mweight | diabe | possible_sarcopenia | 6.7657e-01 | 1.0000e+00 | 9.0210e-01 | Non-significant | 7611 |
| bmi | PM2.5 | possible_sarcopenia | 6.8188e-01 | 1.0000e+00 | 9.0492e-01 | Non-significant | 8135 |
| bmi | drinkev_r | sarcopenia | 6.8207e-01 | 1.0000e+00 | 9.0492e-01 | Non-significant | 15147 |
| bmi | SO2 | sarcopenia | 6.8484e-01 | 1.0000e+00 | 9.0635e-01 | Non-significant | 14931 |
| bmi | sleeprl | sarcopenia | 6.8657e-01 | 1.0000e+00 | 9.0641e-01 | Non-significant | 14823 |
| mwaist | NO2 | possible_sarcopenia | 6.9297e-01 | 1.0000e+00 | 9.0751e-01 | Non-significant | 7976 |
| WWI | PM2.5 | sarcopenia | 6.9341e-01 | 1.0000e+00 | 9.0751e-01 | Non-significant | 14657 |
| WHtR | extreme_high | sarcopenia | 6.9767e-01 | 1.0000e+00 | 9.0751e-01 | Non-significant | 10227 |
| mwaist | PM2.5 | possible_sarcopenia | 7.0167e-01 | 1.0000e+00 | 9.0751e-01 | Non-significant | 7976 |
| CMI | age_group | possible_sarcopenia | 7.0346e-01 | 1.0000e+00 | 9.0751e-01 | Non-significant | 6598 |
| CMI | PM10 | possible_sarcopenia | 7.0358e-01 | 1.0000e+00 | 9.0751e-01 | Non-significant | 6485 |
| WHtR | green_area | sarcopenia | 7.0435e-01 | 1.0000e+00 | 9.0751e-01 | Non-significant | 13519 |
| gripsum | SO2 | sarcopenia | 7.0896e-01 | 1.0000e+00 | 9.0751e-01 | Non-significant | 14868 |
| gripsum | NO2 | possible_sarcopenia | 7.1090e-01 | 1.0000e+00 | 9.0751e-01 | Non-significant | 8205 |
| WHtR | drinkev_r | sarcopenia | 7.1218e-01 | 1.0000e+00 | 9.0751e-01 | Non-significant | 14840 |
| mweight | PM2.5 | sarcopenia | 7.1227e-01 | 1.0000e+00 | 9.0751e-01 | Non-significant | 14946 |
| mwaist | NO2 | sarcopenia | 7.1309e-01 | 1.0000e+00 | 9.0751e-01 | Non-significant | 14682 |
| mwaist | smokev_r | sarcopenia | 7.1440e-01 | 1.0000e+00 | 9.0751e-01 | Non-significant | 14921 |
| ABSI | PM10 | possible_sarcopenia | 7.1545e-01 | 1.0000e+00 | 9.0751e-01 | Non-significant | 7911 |
| WWI | temperature | sarcopenia | 7.1606e-01 | 1.0000e+00 | 9.0751e-01 | Non-significant | 13225 |
| bmi | NDVI | sarcopenia | 7.1610e-01 | 1.0000e+00 | 9.0751e-01 | Non-significant | 13387 |
| wspeed | sleeprl | possible_sarcopenia | 7.1758e-01 | 1.0000e+00 | 9.0751e-01 | Non-significant | 7473 |
| bmi | age_group | possible_sarcopenia | 7.1858e-01 | 1.0000e+00 | 9.0751e-01 | Non-significant | 8263 |
| CMI | extreme_high | possible_sarcopenia | 7.1958e-01 | 1.0000e+00 | 9.0751e-01 | Non-significant | 4522 |
| CMI | sex_age_group | sarcopenia | 7.2481e-01 | 1.0000e+00 | 9.0836e-01 | Non-significant | 12088 |
| mweight | altitude | possible_sarcopenia | 7.2771e-01 | 1.0000e+00 | 9.0836e-01 | Non-significant | 7317 |
| CMI | SO2 | possible_sarcopenia | 7.2803e-01 | 1.0000e+00 | 9.0836e-01 | Non-significant | 6485 |
| mweight | PM10 | sarcopenia | 7.3062e-01 | 1.0000e+00 | 9.0836e-01 | Non-significant | 14946 |
| WHtR | PM2.5 | sarcopenia | 7.3137e-01 | 1.0000e+00 | 9.0836e-01 | Non-significant | 14623 |
| mwaist | SO2 | possible_sarcopenia | 7.3165e-01 | 1.0000e+00 | 9.0836e-01 | Non-significant | 7976 |
| bmi | gripsum | sarcopenia | 7.3328e-01 | 1.0000e+00 | 9.0836e-01 | Non-significant | 15036 |
| WWI | extreme_rain | sarcopenia | 7.3381e-01 | 1.0000e+00 | 9.0836e-01 | Non-significant | 10250 |
| WHtR | extreme_rain | sarcopenia | 7.3983e-01 | 1.0000e+00 | 9.1246e-01 | Non-significant | 10227 |
| gripsum | cesd10 | sarcopenia | 7.4052e-01 | 1.0000e+00 | 9.1246e-01 | Non-significant | 14816 |
| WHtR | sleeprl | possible_sarcopenia | 7.4380e-01 | 1.0000e+00 | 9.1440e-01 | Non-significant | 7754 |
| WWI | SO2 | sarcopenia | 7.4772e-01 | 1.0000e+00 | 9.1695e-01 | Non-significant | 14657 |
| CMI | diabe | possible_sarcopenia | 7.5100e-01 | 1.0000e+00 | 9.1695e-01 | Non-significant | 6099 |
| gripsum | climate_risk | sarcopenia | 7.5101e-01 | 1.0000e+00 | 9.1695e-01 | Non-significant | 10274 |
| bmi | PM2.5 | sarcopenia | 7.5799e-01 | 1.0000e+00 | 9.1944e-01 | Non-significant | 14931 |
| bmi | AQI | sarcopenia | 7.5922e-01 | 1.0000e+00 | 9.1944e-01 | Non-significant | 14228 |
| WWI | PM10 | sarcopenia | 7.5943e-01 | 1.0000e+00 | 9.1944e-01 | Non-significant | 14657 |
| mwaist | climate_risk | possible_sarcopenia | 7.5991e-01 | 1.0000e+00 | 9.1944e-01 | Non-significant | 5550 |
| gripsum | PM10 | possible_sarcopenia | 7.6551e-01 | 1.0000e+00 | 9.2303e-01 | Non-significant | 8205 |
| mwaist | AQI | sarcopenia | 7.6632e-01 | 1.0000e+00 | 9.2303e-01 | Non-significant | 13985 |
| bmi | climate_risk | sarcopenia | 7.6865e-01 | 1.0000e+00 | 9.2376e-01 | Non-significant | 10304 |
| gripsum | O3 | sarcopenia | 7.7904e-01 | 1.0000e+00 | 9.3415e-01 | Non-significant | 14868 |
| ABSI | NO2 | sarcopenia | 7.8167e-01 | 1.0000e+00 | 9.3494e-01 | Non-significant | 14609 |
| WWI | extreme_high | possible_sarcopenia | 7.8318e-01 | 1.0000e+00 | 9.3494e-01 | Non-significant | 5535 |
| mwaist | green_area | sarcopenia | 7.8513e-01 | 1.0000e+00 | 9.3518e-01 | Non-significant | 13571 |
| bmi | PM10 | sarcopenia | 7.8734e-01 | 1.0000e+00 | 9.3573e-01 | Non-significant | 14931 |
| bmi | drinkev_r | possible_sarcopenia | 7.8911e-01 | 1.0000e+00 | 9.3575e-01 | Non-significant | 8244 |
| CMI | extreme_rain | sarcopenia | 7.9343e-01 | 1.0000e+00 | 9.3880e-01 | Non-significant | 8312 |
| mweight | gripsum | sarcopenia | 7.9586e-01 | 1.0000e+00 | 9.3946e-01 | Non-significant | 15054 |
| wspeed | diabe | sarcopenia | 7.9749e-01 | 1.0000e+00 | 9.3946e-01 | Non-significant | 7144 |
| gripsum | CO | possible_sarcopenia | 8.0296e-01 | 1.0000e+00 | 9.4383e-01 | Non-significant | 8205 |
| wspeed | extreme_high | possible_sarcopenia | 8.0712e-01 | 1.0000e+00 | 9.4664e-01 | Non-significant | 5183 |
| CMI | NO2 | possible_sarcopenia | 8.1173e-01 | 1.0000e+00 | 9.4997e-01 | Non-significant | 6485 |
| gripsum | cesd10 | possible_sarcopenia | 8.1933e-01 | 1.0000e+00 | 9.5271e-01 | Non-significant | 8048 |
| CMI | cesd10 | possible_sarcopenia | 8.1941e-01 | 1.0000e+00 | 9.5271e-01 | Non-significant | 6407 |
| WWI | CO | sarcopenia | 8.2280e-01 | 1.0000e+00 | 9.5271e-01 | Non-significant | 14657 |
| mweight | relief | possible_sarcopenia | 8.2322e-01 | 1.0000e+00 | 9.5271e-01 | Non-significant | 7317 |
| CMI | climate_risk | possible_sarcopenia | 8.2362e-01 | 1.0000e+00 | 9.5271e-01 | Non-significant | 4522 |
| CMI | sleeprl | possible_sarcopenia | 8.2557e-01 | 1.0000e+00 | 9.5271e-01 | Non-significant | 6377 |
| CMI | green_area | sarcopenia | 8.2898e-01 | 1.0000e+00 | 9.5271e-01 | Non-significant | 11064 |
| ABSI | relief | possible_sarcopenia | 8.2961e-01 | 1.0000e+00 | 9.5271e-01 | Non-significant | 7160 |
| bmi | PM10 | possible_sarcopenia | 8.3007e-01 | 1.0000e+00 | 9.5271e-01 | Non-significant | 8135 |
| WHtR | rural2 | sarcopenia | 8.4235e-01 | 1.0000e+00 | 9.6387e-01 | Non-significant | 14872 |
| WWI | O3 | sarcopenia | 8.4339e-01 | 1.0000e+00 | 9.6387e-01 | Non-significant | 14657 |
| wspeed | CO | sarcopenia | 8.4923e-01 | 1.0000e+00 | 9.6760e-01 | Non-significant | 7589 |
| wspeed | SO2 | possible_sarcopenia | 8.5097e-01 | 1.0000e+00 | 9.6760e-01 | Non-significant | 7628 |
| WWI | green_area | sarcopenia | 8.5206e-01 | 1.0000e+00 | 9.6760e-01 | Non-significant | 13548 |
| bmi | NO2 | possible_sarcopenia | 8.5761e-01 | 1.0000e+00 | 9.7183e-01 | Non-significant | 8135 |
| gripsum | PM2.5 | possible_sarcopenia | 8.6160e-01 | 1.0000e+00 | 9.7370e-01 | Non-significant | 8205 |
| ABSI | SO2 | sarcopenia | 8.6288e-01 | 1.0000e+00 | 9.7370e-01 | Non-significant | 14609 |
| bmi | relief | possible_sarcopenia | 8.6846e-01 | 1.0000e+00 | 9.7377e-01 | Non-significant | 7305 |
| mwaist | PM2.5 | sarcopenia | 8.6907e-01 | 1.0000e+00 | 9.7377e-01 | Non-significant | 14682 |
| CMI | altitude | possible_sarcopenia | 8.7016e-01 | 1.0000e+00 | 9.7377e-01 | Non-significant | 5951 |
| ABSI | NO2 | possible_sarcopenia | 8.7097e-01 | 1.0000e+00 | 9.7377e-01 | Non-significant | 7911 |
| bmi | AQI | possible_sarcopenia | 8.7378e-01 | 1.0000e+00 | 9.7377e-01 | Non-significant | 7757 |
| CMI | extreme_drought | sarcopenia | 8.7509e-01 | 1.0000e+00 | 9.7377e-01 | Non-significant | 8312 |
| mweight | AQI | sarcopenia | 8.7567e-01 | 1.0000e+00 | 9.7377e-01 | Non-significant | 14242 |
| CMI | precipitation | sarcopenia | 8.8057e-01 | 1.0000e+00 | 9.7719e-01 | Non-significant | 10830 |
| bmi | extreme_low | possible_sarcopenia | 8.8421e-01 | 1.0000e+00 | 9.7720e-01 | Non-significant | 5563 |
| gripsum | extreme_high | sarcopenia | 8.8422e-01 | 1.0000e+00 | 9.7720e-01 | Non-significant | 10274 |
| WHtR | NO2 | possible_sarcopenia | 8.8756e-01 | 1.0000e+00 | 9.7739e-01 | Non-significant | 7929 |
| WHtR | NO2 | sarcopenia | 8.8926e-01 | 1.0000e+00 | 9.7739e-01 | Non-significant | 14623 |
| gripsum | extreme_low | sarcopenia | 8.8986e-01 | 1.0000e+00 | 9.7739e-01 | Non-significant | 10274 |
| mweight | climate_risk | sarcopenia | 8.9581e-01 | 1.0000e+00 | 9.8191e-01 | Non-significant | 10315 |
| ABSI | drinkev_r | possible_sarcopenia | 8.9837e-01 | 1.0000e+00 | 9.8270e-01 | Non-significant | 8019 |
| WWI | PM10 | possible_sarcopenia | 9.0417e-01 | 1.0000e+00 | 9.8612e-01 | Non-significant | 7947 |
| CMI | drinkev_r | sarcopenia | 9.0690e-01 | 1.0000e+00 | 9.8612e-01 | Non-significant | 12064 |
| ABSI | drinkev_r | sarcopenia | 9.0721e-01 | 1.0000e+00 | 9.8612e-01 | Non-significant | 14825 |
| ABSI | precipitation | sarcopenia | 9.0885e-01 | 1.0000e+00 | 9.8612e-01 | Non-significant | 13180 |
| wspeed | smokev_r | possible_sarcopenia | 9.1867e-01 | 1.0000e+00 | 9.8825e-01 | Non-significant | 7748 |
| mweight | SO2 | sarcopenia | 9.2098e-01 | 1.0000e+00 | 9.8825e-01 | Non-significant | 14946 |
| bmi | extreme_drought | sarcopenia | 9.2299e-01 | 1.0000e+00 | 9.8825e-01 | Non-significant | 10304 |
| mwaist | rural2 | sarcopenia | 9.2358e-01 | 1.0000e+00 | 9.8825e-01 | Non-significant | 14934 |
| mweight | extreme_low | possible_sarcopenia | 9.2504e-01 | 1.0000e+00 | 9.8825e-01 | Non-significant | 5573 |
| WHtR | extreme_low | sarcopenia | 9.2586e-01 | 1.0000e+00 | 9.8825e-01 | Non-significant | 10227 |
| wspeed | PM10 | sarcopenia | 9.2738e-01 | 1.0000e+00 | 9.8825e-01 | Non-significant | 7589 |
| gripsum | age_group | possible_sarcopenia | 9.2860e-01 | 1.0000e+00 | 9.8825e-01 | Non-significant | 8338 |
| wspeed | NDVI | possible_sarcopenia | 9.3298e-01 | 1.0000e+00 | 9.8825e-01 | Non-significant | 6873 |
| WWI | precipitation | sarcopenia | 9.3349e-01 | 1.0000e+00 | 9.8825e-01 | Non-significant | 13225 |
| gripsum | sleeprl | sarcopenia | 9.3695e-01 | 1.0000e+00 | 9.8825e-01 | Non-significant | 14764 |
| WWI | gripsum | possible_sarcopenia | 9.3754e-01 | 1.0000e+00 | 9.8825e-01 | Non-significant | 8076 |
| mweight | drinkev_r | sarcopenia | 9.3932e-01 | 1.0000e+00 | 9.8825e-01 | Non-significant | 15165 |
| mwaist | extreme_high | sarcopenia | 9.3993e-01 | 1.0000e+00 | 9.8825e-01 | Non-significant | 10264 |
| ABSI | grip_bmi | sarcopenia | 9.4114e-01 | 1.0000e+00 | 9.8825e-01 | Non-significant | 14716 |
| gripsum | CO | sarcopenia | 9.4293e-01 | 1.0000e+00 | 9.8825e-01 | Non-significant | 14868 |
| CMI | SO2 | sarcopenia | 9.4389e-01 | 1.0000e+00 | 9.8825e-01 | Non-significant | 11877 |
| wspeed | PM2.5 | sarcopenia | 9.4430e-01 | 1.0000e+00 | 9.8825e-01 | Non-significant | 7589 |
| bmi | age_group | sarcopenia | 9.4585e-01 | 1.0000e+00 | 9.8825e-01 | Non-significant | 15179 |
| mwaist | drinkev_r | sarcopenia | 9.4967e-01 | 1.0000e+00 | 9.9032e-01 | Non-significant | 14902 |
| mwaist | altitude | sarcopenia | 9.5402e-01 | 1.0000e+00 | 9.9064e-01 | Non-significant | 13247 |
| mwaist | age_group | sarcopenia | 9.5659e-01 | 1.0000e+00 | 9.9064e-01 | Non-significant | 14934 |
| WWI | NO2 | sarcopenia | 9.5778e-01 | 1.0000e+00 | 9.9064e-01 | Non-significant | 14657 |
| WHtR | extreme_low | possible_sarcopenia | 9.5870e-01 | 1.0000e+00 | 9.9064e-01 | Non-significant | 5521 |
| gripsum | sex_age_group | sarcopenia | 9.5922e-01 | 1.0000e+00 | 9.9064e-01 | Non-significant | 15117 |
| mweight | NDVI | possible_sarcopenia | 9.6439e-01 | 1.0000e+00 | 9.9407e-01 | Non-significant | 7317 |
| WWI | sex_age_group | possible_sarcopenia | 9.7419e-01 | 1.0000e+00 | 9.9987e-01 | Non-significant | 8078 |
| gripsum | PM10 | sarcopenia | 9.7436e-01 | 1.0000e+00 | 9.9987e-01 | Non-significant | 14868 |
| bmi | extreme_rain | sarcopenia | 9.7686e-01 | 1.0000e+00 | 9.9987e-01 | Non-significant | 10304 |
| bmi | extreme_rain | possible_sarcopenia | 9.7938e-01 | 1.0000e+00 | 9.9987e-01 | Non-significant | 5563 |
| bmi | relief | sarcopenia | 9.8239e-01 | 1.0000e+00 | 9.9987e-01 | Non-significant | 13387 |
| bmi | extreme_drought | possible_sarcopenia | 9.8371e-01 | 1.0000e+00 | 9.9987e-01 | Non-significant | 5563 |
| mwaist | extreme_rain | possible_sarcopenia | 9.8512e-01 | 1.0000e+00 | 9.9987e-01 | Non-significant | 5550 |
| WWI | altitude | possible_sarcopenia | 9.8632e-01 | 1.0000e+00 | 9.9987e-01 | Non-significant | 7196 |
| bmi | diabe | sarcopenia | 9.8807e-01 | 1.0000e+00 | 9.9987e-01 | Non-significant | 12488 |
| gripsum | sex_age_group | possible_sarcopenia | 9.9483e-01 | 1.0000e+00 | 9.9987e-01 | Non-significant | 8338 |
| mweight | diabe | sarcopenia | 9.9649e-01 | 1.0000e+00 | 9.9987e-01 | Non-significant | 12508 |
| wspeed | sex_age_group | possible_sarcopenia | 9.9672e-01 | 1.0000e+00 | 9.9987e-01 | Non-significant | 7755 |
| bmi | O3 | possible_sarcopenia | 9.9681e-01 | 1.0000e+00 | 9.9987e-01 | Non-significant | 8135 |
| wspeed | sex_age_group | sarcopenia | 9.9803e-01 | 1.0000e+00 | 9.9987e-01 | Non-significant | 7719 |
| mwaist | extreme_low | possible_sarcopenia | 9.9856e-01 | 1.0000e+00 | 9.9987e-01 | Non-significant | 5550 |
| ABSI | CO | sarcopenia | 9.9987e-01 | 1.0000e+00 | 9.9987e-01 | Non-significant | 14609 |

**Table S2.** Sensitivity analyses of the five core interaction signals. Each core signal was tested under six conditions: (1) the full adjustment model (Model 2: age, sex, education, marital status, smoking, alcohol consumption, multimorbidity; with additional BMI adjustment for waist circumference models); (2) exclusion of triglyceride-truncated observations; (3) exclusion of extreme BMI values (top and bottom 1%); (4) substitution of absolute grip strength for the grip/weight ratio as the modifier; (5) exclusion of participants with grip strength below AWGS 2019 diagnostic thresholds; and (6) substitution of confirmed sarcopenia for possible sarcopenia as the outcome. Interaction p-values are reported for each condition. All five core signals remained Bonferroni-significant across conditions (1)–(3) and (6). The effect modification disappeared under conditions (4) and (5), supporting that the interaction is driven by muscle function rather than mathematical coupling or diagnostic circularity.

| Core signal | Sensitivity analysis | Interaction p |
| --- | --- | --- |
| Waist circumference × grip/weight → possible sarcopenia | Baseline (full adjustment) | 1.0629e-23 |
| BMI × grip/weight → possible sarcopenia | Baseline (full adjustment) | 5.8325e-104 |
| WHtR × grip/weight → possible sarcopenia | Baseline (full adjustment) | 2.8284e-17 |
| Body weight × grip/weight → possible sarcopenia | Baseline (full adjustment) | 4.4144e-113 |
| Waist circumference × grip/weight → sarcopenia | Baseline (full adjustment) | 8.8869e-15 |
| Waist circumference × grip/weight → possible sarcopenia | Excluding TG truncation | 2.2109e-23 |
| BMI × grip/weight → possible sarcopenia | Excluding TG truncation | 1.6008e-102 |
| WHtR × grip/weight → possible sarcopenia | Excluding TG truncation | 6.3893e-17 |
| Body weight × grip/weight → possible sarcopenia | Excluding TG truncation | 5.4670e-112 |
| Waist circumference × grip/weight → sarcopenia | Excluding TG truncation | 3.6745e-14 |
| Waist circumference × grip/weight → possible sarcopenia | Excluding BMI extremes | 6.7223e-36 |
| BMI × grip/weight → possible sarcopenia | Excluding BMI extremes | 4.0356e-43 |
| WHtR × grip/weight → possible sarcopenia | Excluding BMI extremes | 2.9162e-12 |
| Body weight × grip/weight → possible sarcopenia | Excluding BMI extremes | 3.1194e-56 |
| Waist circumference × grip/weight → sarcopenia | Excluding BMI extremes | 2.4888e-13 |
| BMI × absolute grip strength → possible sarcopenia | Absolute grip strength substitution | 6.3400e-01 |
| BMI × grip/weight → possible sarcopenia | Excluding low grip (< AWGS) | 6.4900e-01 |

**Table S3.** Mediation analysis of metabolic pathways linking waist circumference to possible sarcopenia. The total effect represents the overall association between waist circumference and sarcopenia risk. The average direct effect (ADE) represents the effect not transmitted through the mediators. The average causal mediation effect (ACME) represents the effect transmitted through each mediator. The proportion mediated was calculated as ACME divided by the total effect × 100%. Bootstrapped 95% confidence intervals are based on 1,000 simulations. All models were adjusted for age, sex, education, marital status, smoking status, alcohol consumption, multimorbidity, and BMI. Abbreviations: CRP, C-reactive protein; TG, triglycerides.

| Mediator | ACME | ACME p | ADE | ADE p | Total effect | Proportion mediated |
| --- | --- | --- | --- | --- | --- | --- |
| CRP (Inflammation) | 0.000091 | 1.8000e-02 | -0.004130 | 1.4000e-02 | -0.004039 | -2.2% |
| Glucose (Insulin resistance) | 0.000029 | 8.1000e-01 | -0.004093 | 8.0000e-03 | -0.004064 | -0.7% |
| TG (Lipid metabolism) | -0.000426 | 2.2000e-02 | -0.003631 | 4.0000e-02 | -0.004058 | 10.5% |

